## Supplementary Material for "A mathematical model of H5N1 influenza transmission in US dairy cattle"

List of Figures 38

List of Tables 39

### 1 US Cattle Populations

Data on the number of dairy cattle in each of the 48 continental US states is taken from the United States 2022 Census of Agriculture [1]. Table S1 below details the information extracted from the census on the total number of dairy cattle and dairy herds in each US state. It also lists the mean herd size (number of cattle divided by number of herds), however the census does provide more detailed information in the distribution of herd sizes which is used within the model.

Figure S1 plots the mean herd size by US state, ordered left-to-right from the state with the largest total dairy cow population (California, 1,688,202 cows) to the smallest (Rhode Island, 722 cows). Those states that have declared outbreaks as of December 2nd 2024 are shaded in red. The plot demonstrates how the total number of cows per state does not align with mean herd size. For example, Wisconsin has the second largest total number of dairy cows in the country, but has a far smaller mean herd size than California, as its cattle population is distributed across a large number of smaller holdings.

**Table S1:** The total number of dairy cattle and the number of dairy herds in each of the 48 continental US states (Hawaii and Alaska not included).

| State | Total Cattle Population | Total Herds | Mean Herd Size |
|---|---|---|---|
| Alabama | 2,225 | 92 | 24.18 |
| Arizona | 199,279 | 122 | 1,633.43 |
| Arkansas | 3,820 | 75 | 50.93 |
| California | 1,688,202 | 1,117 | 1,511.37 |
| Colorado | 201,750 | 414 | 487.32 |
| Connecticut | 18,889 | 145 | 130.27 |
| Delaware | 2,528 | 41 | 61.66 |
| Florida | 94,069 | 369 | 254.93 |
| Georgia | 72,830 | 391 | 186.27 |
| Idaho | 664,479 | 549 | 1,210.34 |
| Illinois | 74,918 | 607 | 123.42 |
| Indiana | 183,176 | 1,521 | 120.43 |
| Iowa | 238,087 | 1,016 | 234.34 |
| Kansas | 175,670 | 501 | 350.64 |
| Kentucky | 45,904 | 1,062 | 43.22 |
| Louisiana | 7,796 | 74 | 105.35 |
| Maine | 24,836 | 292 | 85.05 |
| Maryland | 38,290 | 342 | 111.96 |
| Massachusetts | 9,998 | 151 | 66.21 |
| Michigan | 436,254 | 1,481 | 294.57 |
| Minnesota | 459,536 | 2,185 | 210.31 |
| Mississippi | 6,904 | 45 | 153.42 |
| Missouri | 63,882 | 1,704 | 37.49 |
| Montana | 10,841 | 273 | 39.71 |
| Nebraska | 57,036 | 300 | 190.12 |
| Nevada | 32,021 | 48 | 667.10 |
| New Hampshire | 11,451 | 129 | 88.77 |
| New Jersey | 4,563 | 95 | 48.03 |
| New Mexico | 279,664 | 233 | 1,200.27 |
| New York | 631,199 | 2,783 | 226.81 |
| North Carolina | 36,127 | 402 | 89.87 |
| North Dakota | 14,191 | 107 | 132.63 |
| Ohio | 247,743 | 2,195 | 112.87 |
| Oklahoma | 40,457 | 298 | 135.76 |
| Oregon | 117,533 | 516 | 227.78 |
| Pennsylvania | 455,651 | 4,027 | 113.15 |
| Rhode Island | 722 | 13 | 55.54 |
| South Carolina | 10,203 | 153 | 66.69 |
| South Dakota | 188,764 | 255 | 740.25 |
| Tennessee | 24,525 | 680 | 36.07 |
| Texas | 639,506 | 479 | 1,335.09 |
| Utah | 91,505 | 334 | 273.97 |

| | | | |
|---|---|---|---|
| Vermont | 106,632 | 528 | 201.95 |
| Virginia | 65,218 | 703 | 92.77 |
| Washington | 255,872 | 419 | 610.67 |
| West Virginia | 4,268 | 335 | 12.74 |
| Wisconsin | 1,264,272 | 6,216 | 203.39 |
| Wyoming | 5,421 | 157 | 34.53 |

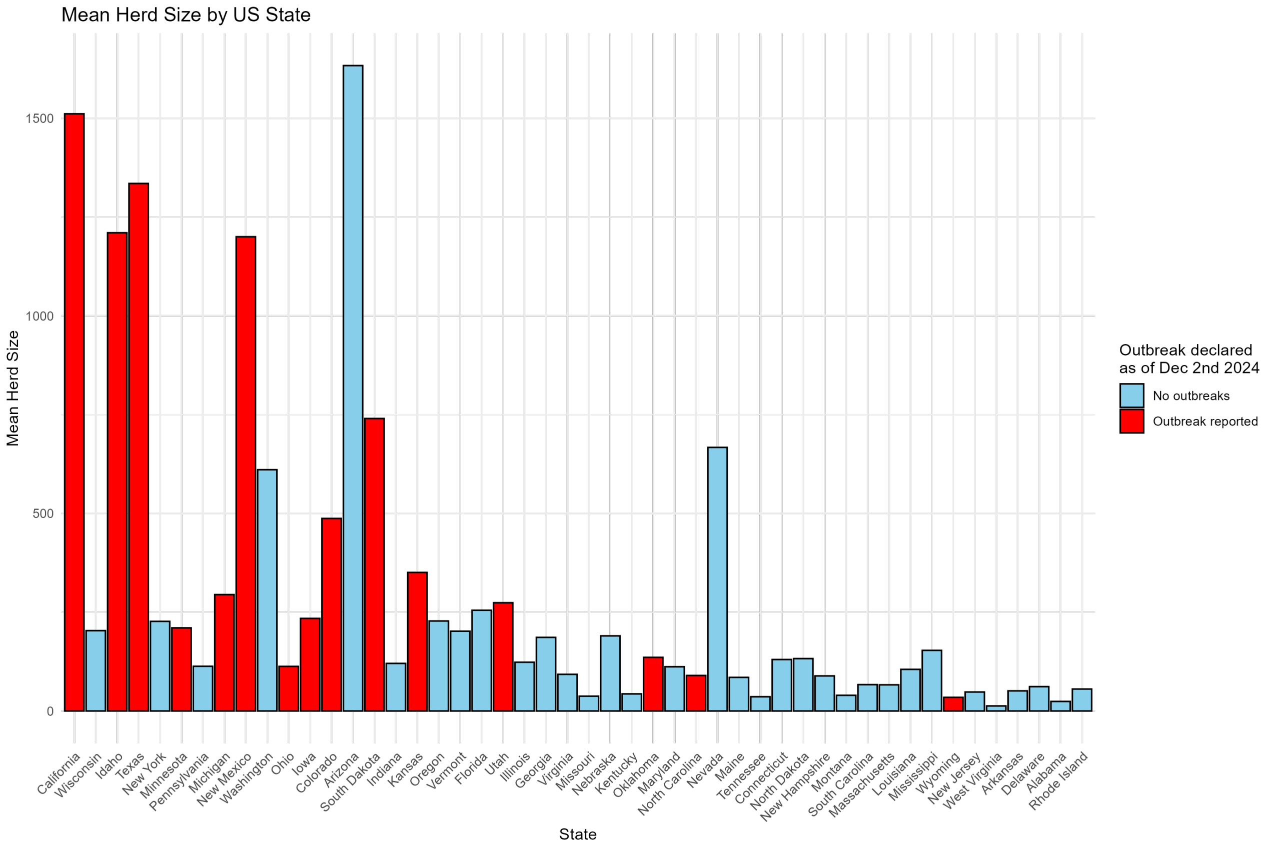

**Figure S1:** The mean herd size (number of dairy cows divided by number of herds) for each of the 48 continental US states. States are presented, left-to-right, in order of largest total dairy cow population to smallest. States highlighted in red are those that have reported an outbreak of H5N1 in dairy cattle as of December 2nd 2024.

### 2 Methods

#### 2.1 Model Equations

In this section we provide a more extensive and in-depth explanation of the underlying model mechanics. A metapopulation model of 35,974 herds, modelled as 35,974 separate SEIR frameworks is built, with a total of 9,308,707 dairy cows distributed across these 35,974 herds. Index $i$ refers to the herd, and index $j$ to the US state. Each cow in herd $\{i, j\}$ is compartmentalised within either the susceptible, $S_{i,j}$, exposed, $E_{i,j}$, infected (and infectious), $I_{i,j}$, or recovered, $R_{i,j}$, compartments. Thus, $S_{i,j}$ refers to the susceptible cows in herd $i$ in state $j$. The basic epidemiological dynamics of the spread of infection within each individual herd can be described by the following series of ODEs:

$$\frac{dS_{i,j}}{dt} = -\beta \frac{S_{i,j}I_{i,j}}{N_{i,j}}, \tag{1}$$

$$\frac{dE_{i,j}}{dt} = \beta \frac{S_{i,j}I_{i,j}}{N_{i,j}} - \sigma E_{i,j}, \tag{2}$$

$$\frac{dI_{i,j}}{dt} = \sigma E_{i,j} - \gamma I_{i,j}, \tag{3}$$

$$\frac{dR_{i,j}}{dt} = \gamma I_{i,j}. \tag{4}$$

All model parameters are presented in Table S2. $N_{i,j}$ is the total number of cows in herd $i$ of state $j$: $N_{i,j} = S_{i,j} + E_{i,j} + I_{i,j} + R_{i,j}$.

The above equations currently do not allow for any infections to occur across or between herds. Recognising that many neighbouring dairy herds will share workers, grazing areas, equipment, or be exposed to environmental run-off from one-another, we adapt equations (1) and (2) above to allow for herds within the same US state to cause infections between one-another. This transmission occurs at a reduced rate controlled by the parameter $\alpha$:

$$\frac{dS_{i,j}}{dt} = -\beta \frac{S_{i,j}I_{i,j}}{N_{i,j}} - \beta S_{i,j}\alpha \left( \sum_{k=1,k\neq i}^{N^j_{\text{herds}}} \frac{I_{k,j}}{N_j - N_{k,j}} \right), \tag{5}$$

$$\frac{dE_{i,j}}{dt} = \beta \frac{S_{i,j}I_{i,j}}{N_{i,j}} + \beta S_{i,j}\alpha \left( \sum_{k=1,k\neq i}^{N^j_{\text{herds}}} \frac{I_{k,j}}{N_j - N_{k,j}} \right) - \sigma E_{i,j}, \tag{6}$$

$$\frac{dI_{i,j}}{dt} = \sigma E_{i,j} - \gamma I_{i,j}, \tag{7}$$

$$\frac{dR_{i,j}}{dt} = \gamma I_{i,j}. \tag{8}$$

Here, $N^j_{\text{herds}}$ is the total number of herds in state $j$, and $N_j$ is the total number of cows in state $j$.

| Parameter | Definition |
|---|---|
| $\beta$ | Transmission rate. |
| $\alpha$ | Proportion decrease of transmission rate between cows in different herds, but within the same state. |
| $\sigma$ | Incubation rate. |
| $\gamma$ | Recovery rate. |
| $A^{\text{asc}}$ | Global ascertainment rate scaling. A model parameter controlling how many infections are reported. |

**Table S2:** Definitions of model parameters.

Rather than use the above deterministic series of ODEs, we model the transition between epidemiological states via the stochastic interpretation of equations (5) - (8). We calculate the number of cattle progressing between epidemiological compartments via binomial distributions, for each timestep $dt$ as:

$$n_{SE} \sim \text{Binomial}\left( S_{i,j}, 1 - \exp\left( - \left( \beta \frac{S_{i,j}I_{i,j}}{N_{i,j}} + \beta S_{i,j}\alpha \sum_{k=1,k\neq i}^{N^j_{\text{herds}}} \frac{I_{k,j}}{N_j - N_{k,j}} \right) dt \right) \right), \tag{9}$$

$$n_{EI} \sim \text{Binomial}\left(E_{i,j}, 1-\exp(-\sigma\, dt)\right), \tag{10}$$
$$n_{IR} \sim \text{Binomial}\left(I_{i,j}, 1-\exp(-\gamma dt)\right). \tag{11}$$

Here $n_{XY}$ is the number of cattle moved from compartment $X$ to $Y$ (for general $X$ and $Y$) in a time step of size $dt$. $n_{SE}$, $n_{EI}$, and $n_{IR}$ is calculated for each herd.

#### 2.2 Model Logic

The previous section outlined the underlying epidemiological dynamics dictating the movement of cattle between epidemiological compartments. However, within each timestep of model evaluation, we also calculate multiple other movements between compartments, and construct other model output variables. We probabilistically calculate the movement of cattle between herds and states, as is frequently done within the dairy industry. We also probabilistically calculate whether or not such exports proceed subject to border testing at certain periods of time. We also calculate whether or not each herd will declare an outbreak present at each time point. Algorithm 1 below outlines the mechanical steps taken at each time point in pseudocode. We move cattle between epidemiological compartments following the methods outlined above in equations (9) - (11). We then calculate whether or not each herd reports an outbreak. We then simulate the shipment of cattle between herds for each herd.

**Algorithm 1** Model calculations at each timestep.

```
for each time t do
    for each herd i do
        Calculate and move all cows between epidemiological compartments.
    end for

    for each herd i do
        Calculate whether or not herd i reports an outbreak.
    end for

    for each herd i do
        Stochastically decide whether or not any cows will be exported from herd i.
        if cows will be exported then
            Stochastically decide which herd the exported cattle will be exported to.
            if Destination herd is in a different state then
                Simulate the testing of up to 30 random cows in the cohort for H5N1.
                if H5N1 detected then
                    Cancel export.
                else
                    Move cattle from herd i to their same epidemiological compartments in the destination herd.
                end if
            end if
        end if
    end for

end for
```

The entire model code can be seen in the `inst/dust/cows.cpp` file within our associated `cowflu` package. We explain each segment in greater detail in the following subsections.

#### 2.3 Reporting an outbreak

After moving cattle between epidemiological compartments, we then stochastically calculate, for each herd, whether or not that herd will report an outbreak in that time step. If a herd has previously reported an outbreak, we skip this herd - a herd cannot report an outbreak twice. For each herd, we calculate the baseline probability of declaring an outbreak, $\phi_i$, as being dependent on the number of infected cattle in the herd, $I_i$, and on the proportion of the herd that is infected $\frac{I_i}{N_i}$:

$$\phi_i = \left(\frac{I_i}{(0.7N_i)^{0.95}} + \frac{I_i}{150}\right) A^{\text{asc}}\, dt, \tag{12}$$

where $A^{\text{asc}}$ is a model parameter that we fit, and $dt$ is the size of the timestep. Figure 3A in the main manuscript plots the form of this function for differing values of $I_i$ and $N_i$. In general, the probability of declaring an outbreak increases as the total number of infected cattle increases, and as the proportion of the herd that is infected increases.

To ensure this probability is always bounded between 0 and 1 for large time steps, we scale $\phi_i$ via: $P_i^{\text{outbreak}} = 1 - e^{-\phi_i}$. For each time step, for each herd, we draw a random number, $X$, from a standard Uniform(0,1) distribution, and record a reported outbreak for that herd at that timepoint if $X < P_i^{\text{outbreak}}$.

#### 2.4 Moving cattle between herds.

After calculating which herds report new outbreaks, we simulate the trade of cattle between premises across the country. For each herd, we first ascertain, stochastically, whether or not each herd will, or will not, export cattle this time step. The US Animal Movement Model (USAMM) provides model simulations of the annual movement of dairy cattle between premises [2]. The USAMM is trained on real world Interstate Certificates of Veterinary Inspection (ICVI) data. Whenever cattle are moved across state lines, an ICVI must be issued as record of the movement. These are mostly paper records which were accessed on request by the developers of the USAMM. 1000 stochastic realisations are provided from the USAMM model at `https://mountainscholar.org/items/75fd3841-11c3-4cec-91b5-45d36b8c6b5a`. Each realisation is a simulated annual ledger of all animal movements between premises (including the number of cattle shipped) as based on previous real world exports. It is from this dataset that we infer our probabilities of whether or not a herd will export cattle at any given time step.

We sum the total number of exports from each state, and divide by the total number of herds in that state, to get the mean number of exports from each herd, annually. We divide this value by $\frac{7}{365}$ for the mean weekly number of exports from each herd for each state. This value is then further scaled by the size of the timestep $dt$. We present these state specific "probabilities of exporting cattle each week" for a herd in Table S3.

**Table S3:** The probability, for each state, that a dairy herd within that state will export dairy cattle in a 7-day period.

| State | Weekly probability of a herd exporting cattle |
|---|---|
| Alabama | 0.007901 |
| Arizona | 0.087296 |
| Arkansas | 0.057525 |
| California | 0.298799 |
| Colorado | 0.025591 |
| Connecticut | 0.020105 |
| Delaware | 0.010928 |
| Florida | 0.069742 |
| Georgia | 0.020383 |
| Idaho | 0.045670 |
| Illinois | 0.012280 |
| Indiana | 0.007776 |
| Iowa | 0.033667 |
| Kansas | 0.016292 |
| Kentucky | 0.012766 |
| Louisiana | 0.006318 |
| Maine | 0.004024 |
| Maryland | 0.017974 |
| Massachusetts | 0.008827 |
| Michigan | 0.014362 |
| Minnesota | 0.019918 |
| Mississippi | 0.026062 |
| Missouri | 0.003321 |
| Montana | 0.008265 |
| Nebraska | 0.004062 |
| Nevada | 0.017928 |
| New Hampshire | 0.005835 |
| New Jersey | 0.017042 |
| New Mexico | 0.056869 |
| New York | 0.046736 |
| North Carolina | 0.010130 |
| North Dakota | 0.044735 |

| | |
|---|---|
| Ohio | 0.005976 |
| Oklahoma | 0.024752 |
| Oregon | 0.011311 |
| Pennsylvania | 0.004942 |
| Rhode Island | 0.009117 |
| South Carolina | 0.035950 |
| South Dakota | 0.014929 |
| Tennessee | 0.017748 |
| Texas | 0.076588 |
| Utah | 0.019551 |
| Vermont | 0.015776 |
| Virginia | 0.009487 |
| Washington | 0.039005 |
| West Virginia | 0.009223 |
| Wisconsin | 0.011569 |
| Wyoming | 0.003867 |

At each time step of the model, for each herd, we draw a random number from the standard uniform distribution, $X \sim$ Uniform(0,1). If $X$ is less than the above values, that herd is denoted as exporting cattle in that timestep. We then calculate how many cattle from the herd will be exported. The USAMM simulated data also includes simulated numbers of cattle exported, and the size of the herd of origin. From this we can average, across all simulations and all exports of cattle, the mean proportion of a herd that is exported for each US state. These values will differ as some states will prioritise breeding and selling of milk cattle, while others will import more calves for milk stock. We notate this proportion as $P_j^{\text{export size}}$, the average proportion of a herd exported from state $j$. We then randomly sample cows to be exported from the 4 epidemiological compartments of the origin herd, where $S_{i,j}^{\text{exported}}$ is the number of cows that will be removed (exported) from the susceptible compartment of herd $i$ in state $j$, and likewise for the exposed, infected, and recovered compartments:

$$S_{i,j}^{\text{exported}} \sim \text{Binomial}\left(S_{i,j}, P_j^{\text{export size}}\right), \tag{13}$$

$$E_{i,j}^{\text{exported}} \sim \text{Binomial}\left(E_{i,j}, P_j^{\text{export size}})\right), \tag{14}$$

$$I_{i,j}^{\text{exported}} \sim \text{Binomial}\left(I_{i,j}, P_j^{\text{export size}})\right). \tag{15}$$

$$R_{i,j}^{\text{exported}} \sim \text{Binomial}\left(R_{i,j}, P_j^{\text{export size}})\right). \tag{16}$$

If $N_{i,j}^{\text{exported}} = 0$, i.e. no cattle are drawn to be exported, then we repeat the draws of cattle exported, to ensure that there is always at least one cow exported from a herd that has already been probabilistically defined as exporting cattle that time step. We list the values of $P_j^{\text{export size}}$ in Table S4 below:

**Table S4:** The mean proportion of a herd's population that is exported by state.

| State | Mean proportion of origin herd's total population that is exported. |
|---|---|
| Alabama | 0.043641 |
| Arizona | 0.043284 |
| Arkansas | 0.049256 |
| California | 0.042684 |
| Colorado | 0.043804 |
| Connecticut | 0.044499 |
| Delaware | 0.047330 |
| Florida | 0.041638 |
| Georgia | 0.044375 |
| Idaho | 0.043360 |
| Illinois | 0.041901 |
| Indiana | 0.049943 |
| Iowa | 0.046084 |
| Kansas | 0.046716 |

| | |
|---|---|
| Kentucky | 0.049534 |
| Louisiana | 0.036121 |
| Maine | 0.039004 |
| Maryland | 0.043558 |
| Massachusetts | 0.046212 |
| Michigan | 0.043190 |
| Minnesota | 0.045885 |
| Mississippi | 0.041946 |
| Missouri | 0.052611 |
| Montana | 0.048579 |
| Nebraska | 0.036096 |
| Nevada | 0.034981 |
| New Hampshire | 0.046465 |
| New Jersey | 0.044953 |
| New Mexico | 0.040021 |
| New York | 0.045065 |
| North Carolina | 0.048999 |
| North Dakota | 0.047075 |
| Ohio | 0.046338 |
| Oklahoma | 0.052091 |
| Oregon | 0.043457 |
| Pennsylvania | 0.046188 |
| Rhode Island | 0.038215 |
| South Carolina | 0.045295 |
| South Dakota | 0.049772 |
| Tennessee | 0.049928 |
| Texas | 0.042814 |
| Utah | 0.046098 |
| Vermont | 0.053861 |
| Virginia | 0.048895 |
| Washington | 0.043427 |
| West Virginia | 0.063624 |
| Wisconsin | 0.045235 |
| Wyoming | 0.060808 |

We then stochastically decide which herd, and in which state, the exported cattle will be delivered to. The USAMM simulated annual exports data allows us to identify the most common inter-state trading partners for each state. We construct a 48 × 48 movement matrix $M$, where element $M_{i,j}$ is the probability that cattle exported from state $i$, will be exported to state $j$. The rows of matrix $M$ thus all sum to 1. To calculate each value $M_{i,j}$, we tally the total number of exports from state $i$ to state $j$ across all realisations and dates from the USAMM model, and divide by the total number of exports from state $i$. We present these values in Table S5, however they are also available within our `cowflu` package via `cowflu::movement$movement_matrix`. As expected, these values mostly align with how geographically close states are, and the size of the destination state's respective dairy industry. We also see that the highest probability of destination is usually (but not always) for the export to go to a different herd within the same state.

**Table S5:** The probability that a given export from an origin state will be sent to a particular other state.

| Origin | Destination | Probability | Origin | Destination | Probability | Origin | Destination | Probability |
|---|---|---|---|---|---|---|---|---|
| Alabama | Alabama | 0.07868 | Alabama | Arizona | 0.00592 | Alabama | Arkansas | 0.00628 |
| Alabama | California | 0.01089 | Alabama | Colorado | 0.00366 | Alabama | Connecticut | 0.00060 |
| Alabama | Delaware | 0.00042 | Alabama | Florida | 0.23364 | Alabama | Georgia | 0.14907 |
| Alabama | Idaho | 0.00331 | Alabama | Illinois | 0.01599 | Alabama | Indiana | 0.03187 |
| Alabama | Iowa | 0.00876 | Alabama | Kansas | 0.00563 | Alabama | Kentucky | 0.05738 |
| Alabama | Louisiana | 0.00502 | Alabama | Maine | 0.00100 | Alabama | Maryland | 0.00344 |
| Alabama | Massachusetts | 0.00108 | Alabama | Michigan | 0.02209 | Alabama | Minnesota | 0.01701 |
| Alabama | Mississippi | 0.02219 | Alabama | Missouri | 0.01875 | Alabama | Montana | 0.00034 |
| Alabama | Nebraska | 0.00055 | Alabama | Nevada | 0.00024 | Alabama | New Hampshire | 0.00045 |
| Alabama | New Jersey | 0.00079 | Alabama | New Mexico | 0.00557 | Alabama | New York | 0.01438 |
| Alabama | North Carolina | 0.01189 | Alabama | North Dakota | 0.00074 | Alabama | Ohio | 0.02585 |
| Alabama | Oklahoma | 0.00655 | Alabama | Oregon | 0.00055 | Alabama | Pennsylvania | 0.01412 |
| Alabama | Rhode Island | 0.00000 | Alabama | South Carolina | 0.00797 | Alabama | South Dakota | 0.00176 |
| Alabama | Tennessee | 0.11249 | Alabama | Texas | 0.03894 | Alabama | Utah | 0.00142 |
| Alabama | Vermont | 0.00452 | Alabama | Virginia | 0.00734 | Alabama | Washington | 0.00221 |
| Alabama | West Virginia | 0.00137 | Alabama | Wisconsin | 0.03705 | Alabama | Wyoming | 0.00024 |
| Arizona | Alabama | 0.00039 | Arizona | Arizona | 0.68777 | Arizona | Arkansas | 0.00113 |
| Arizona | California | 0.12023 | Arizona | Colorado | 0.01213 | Arizona | Connecticut | 0.00028 |

| | | | | | | | | |
|---|---|---|---|---|---|---|---|---|
| Arizona | Delaware | 0.00005 | Arizona | Florida | 0.00632 | Arizona | Georgia | 0.00286 |
| Arizona | Idaho | 0.01405 | Arizona | Illinois | 0.00230 | Arizona | Indiana | 0.00442 |
| Arizona | Iowa | 0.00395 | Arizona | Kansas | 0.00509 | Arizona | Kentucky | 0.00376 |
| Arizona | Louisiana | 0.00057 | Arizona | Maine | 0.00037 | Arizona | Maryland | 0.00056 |
| Arizona | Massachusetts | 0.00029 | Arizona | Michigan | 0.00516 | Arizona | Minnesota | 0.00812 |
| Arizona | Mississippi | 0.00060 | Arizona | Missouri | 0.00377 | Arizona | Montana | 0.00124 |
| Arizona | Nebraska | 0.00066 | Arizona | Nevada | 0.00135 | Arizona | New Hampshire | 0.00012 |
| Arizona | New Jersey | 0.00015 | Arizona | New Mexico | 0.02667 | Arizona | New York | 0.00291 |
| Arizona | North Carolina | 0.00095 | Arizona | North Dakota | 0.00067 | Arizona | Ohio | 0.00366 |
| Arizona | Oklahoma | 0.00408 | Arizona | Oregon | 0.00211 | Arizona | Pennsylvania | 0.00255 |
| Arizona | Rhode Island | 0.00002 | Arizona | South Carolina | 0.00040 | Arizona | South Dakota | 0.00230 |
| Arizona | Tennessee | 0.00218 | Arizona | Texas | 0.03098 | Arizona | Utah | 0.01156 |
| Arizona | Vermont | 0.00136 | Arizona | Virginia | 0.00076 | Arizona | Washington | 0.00644 |
| Arizona | West Virginia | 0.00023 | Arizona | Wisconsin | 0.01146 | Arizona | Wyoming | 0.00104 |
| Arkansas | Alabama | 0.00358 | Arkansas | Arizona | 0.01291 | Arkansas | Arkansas | 0.09951 |
| Arkansas | California | 0.02579 | Arkansas | Colorado | 0.01151 | Arkansas | Connecticut | 0.00101 |
| Arkansas | Delaware | 0.00022 | Arkansas | Florida | 0.03551 | Arkansas | Georgia | 0.01901 |
| Arkansas | Idaho | 0.00784 | Arkansas | Illinois | 0.02795 | Arkansas | Indiana | 0.03710 |
| Arkansas | Iowa | 0.02351 | Arkansas | Kansas | 0.02848 | Arkansas | Kentucky | 0.04166 |
| Arkansas | Louisiana | 0.00720 | Arkansas | Maine | 0.00119 | Arkansas | Maryland | 0.00313 |
| Arkansas | Massachusetts | 0.00103 | Arkansas | Michigan | 0.02997 | Arkansas | Minnesota | 0.03508 |
| Arkansas | Mississippi | 0.01022 | Arkansas | Missouri | 0.13578 | Arkansas | Montana | 0.00098 |
| Arkansas | Nebraska | 0.00210 | Arkansas | Nevada | 0.00036 | Arkansas | New Hampshire | 0.00045 |
| Arkansas | New Jersey | 0.00062 | Arkansas | New Mexico | 0.01511 | Arkansas | New York | 0.01400 |
| Arkansas | North Carolina | 0.00521 | Arkansas | North Dakota | 0.00147 | Arkansas | Ohio | 0.02511 |
| Arkansas | Oklahoma | 0.05609 | Arkansas | Oregon | 0.00128 | Arkansas | Pennsylvania | 0.01360 |
| Arkansas | Rhode Island | 0.00006 | Arkansas | South Carolina | 0.00257 | Arkansas | South Dakota | 0.00613 |
| Arkansas | Tennessee | 0.02818 | Arkansas | Texas | 0.14674 | Arkansas | Utah | 0.00348 |
| Arkansas | Vermont | 0.00576 | Arkansas | Virginia | 0.00473 | Arkansas | Washington | 0.00451 |
| Arkansas | West Virginia | 0.00096 | Arkansas | Wisconsin | 0.06033 | Arkansas | Wyoming | 0.00100 |
| California | Alabama | 0.00004 | California | Arizona | 0.01048 | California | Arkansas | 0.00009 |
| California | California | 0.95808 | California | Colorado | 0.00125 | California | Connecticut | 0.00002 |
| California | Delaware | 0.00001 | California | Florida | 0.00071 | California | Georgia | 0.00026 |
| California | Idaho | 0.00597 | California | Illinois | 0.00027 | California | Indiana | 0.00052 |
| California | Iowa | 0.00044 | California | Kansas | 0.00053 | California | Kentucky | 0.00034 |
| California | Louisiana | 0.00005 | California | Maine | 0.00003 | California | Maryland | 0.00008 |
| California | Massachusetts | 0.00004 | California | Michigan | 0.00062 | California | Minnesota | 0.00130 |
| California | Mississippi | 0.00006 | California | Missouri | 0.00047 | California | Montana | 0.00026 |
| California | Nebraska | 0.00007 | California | Nevada | 0.00243 | California | New Hampshire | 0.00001 |
| California | New Jersey | 0.00002 | California | New Mexico | 0.00164 | California | New York | 0.00039 |
| California | North Carolina | 0.00009 | California | North Dakota | 0.00015 | California | Ohio | 0.00044 |
| California | Oklahoma | 0.00037 | California | Oregon | 0.00142 | California | Pennsylvania | 0.00033 |
| California | Rhode Island | 0.00000 | California | South Carolina | 0.00005 | California | South Dakota | 0.00028 |
| California | Tennessee | 0.00022 | California | Texas | 0.00265 | California | Utah | 0.00248 |
| California | Vermont | 0.00018 | California | Virginia | 0.00008 | California | Washington | 0.00305 |
| California | West Virginia | 0.00002 | California | Wisconsin | 0.00143 | California | Wyoming | 0.00029 |
| Colorado | Alabama | 0.00075 | Colorado | Arizona | 0.02880 | Colorado | Arkansas | 0.00292 |
| Colorado | California | 0.05609 | Colorado | Colorado | 0.49104 | Colorado | Connecticut | 0.00053 |
| Colorado | Delaware | 0.00009 | Colorado | Florida | 0.01070 | Colorado | Georgia | 0.00519 |
| Colorado | Idaho | 0.02585 | Colorado | Illinois | 0.00636 | Colorado | Indiana | 0.01124 |
| Colorado | Iowa | 0.01546 | Colorado | Kansas | 0.03504 | Colorado | Kentucky | 0.00874 |
| Colorado | Louisiana | 0.00093 | Colorado | Maine | 0.00076 | Colorado | Maryland | 0.00125 |
| Colorado | Massachusetts | 0.00050 | Colorado | Michigan | 0.01381 | Colorado | Minnesota | 0.03263 |
| Colorado | Mississippi | 0.00112 | Colorado | Missouri | 0.01156 | Colorado | Montana | 0.00400 |
| Colorado | Nebraska | 0.00697 | Colorado | Nevada | 0.00116 | Colorado | New Hampshire | 0.00025 |
| Colorado | New Jersey | 0.00030 | Colorado | New Mexico | 0.02811 | Colorado | New York | 0.00694 |
| Colorado | North Carolina | 0.00166 | Colorado | North Dakota | 0.00348 | Colorado | Ohio | 0.00900 |
| Colorado | Oklahoma | 0.01224 | Colorado | Oregon | 0.00252 | Colorado | Pennsylvania | 0.00597 |
| Colorado | Rhode Island | 0.00004 | Colorado | South Carolina | 0.00079 | Colorado | South Dakota | 0.01415 |
| Colorado | Tennessee | 0.00456 | Colorado | Texas | 0.05872 | Colorado | Utah | 0.01751 |
| Colorado | Vermont | 0.00295 | Colorado | Virginia | 0.00159 | Colorado | Washington | 0.00953 |
| Colorado | West Virginia | 0.00035 | Colorado | Wisconsin | 0.03459 | Colorado | Wyoming | 0.01128 |
| Connecticut | Alabama | 0.00071 | Connecticut | Arizona | 0.00306 | Connecticut | Arkansas | 0.00096 |
| Connecticut | California | 0.00745 | Connecticut | Colorado | 0.00221 | Connecticut | Connecticut | 0.14814 |
| Connecticut | Delaware | 0.00190 | Connecticut | Florida | 0.01868 | Connecticut | Georgia | 0.00907 |
| Connecticut | Idaho | 0.00259 | Connecticut | Illinois | 0.00613 | Connecticut | Indiana | 0.01820 |
| Connecticut | Iowa | 0.00552 | Connecticut | Kansas | 0.00247 | Connecticut | Kentucky | 0.01105 |
| Connecticut | Louisiana | 0.00050 | Connecticut | Maine | 0.01403 | Connecticut | Maryland | 0.01796 |
| Connecticut | Massachusetts | 0.12186 | Connecticut | Michigan | 0.02780 | Connecticut | Minnesota | 0.01323 |
| Connecticut | Mississippi | 0.00097 | Connecticut | Missouri | 0.00567 | Connecticut | Montana | 0.00034 |
| Connecticut | Nebraska | 0.00035 | Connecticut | Nevada | 0.00024 | Connecticut | New Hampshire | 0.01614 |
| Connecticut | New Jersey | 0.01352 | Connecticut | New Mexico | 0.00244 | Connecticut | New York | 0.19330 |
| Connecticut | North Carolina | 0.00680 | Connecticut | North Dakota | 0.00063 | Connecticut | Ohio | 0.03094 |
| Connecticut | Oklahoma | 0.00165 | Connecticut | Oregon | 0.00055 | Connecticut | Pennsylvania | 0.10154 |
| Connecticut | Rhode Island | 0.00941 | Connecticut | South Carolina | 0.00218 | Connecticut | South Dakota | 0.00134 |
| Connecticut | Tennessee | 0.00732 | Connecticut | Texas | 0.01010 | Connecticut | Utah | 0.00131 |
| Connecticut | Vermont | 0.11241 | Connecticut | Virginia | 0.00913 | Connecticut | Washington | 0.00214 |
| Connecticut | West Virginia | 0.00250 | Connecticut | Wisconsin | 0.03324 | Connecticut | Wyoming | 0.00029 |
| Delaware | Alabama | 0.00158 | Delaware | Arizona | 0.00383 | Delaware | Arkansas | 0.00170 |
| Delaware | California | 0.00988 | Delaware | Colorado | 0.00349 | Delaware | Connecticut | 0.01057 |
| Delaware | Delaware | 0.03920 | Delaware | Florida | 0.02450 | Delaware | Georgia | 0.01704 |
| Delaware | Idaho | 0.00366 | Delaware | Illinois | 0.00605 | Delaware | Indiana | 0.02411 |
| Delaware | Iowa | 0.00626 | Delaware | Kansas | 0.00366 | Delaware | Kentucky | 0.02497 |
| Delaware | Louisiana | 0.00089 | Delaware | Maine | 0.00571 | Delaware | Maryland | 0.13842 |
| Delaware | Massachusetts | 0.00741 | Delaware | Michigan | 0.03485 | Delaware | Minnesota | 0.01316 |
| Delaware | Mississippi | 0.00098 | Delaware | Missouri | 0.00516 | Delaware | Montana | 0.00026 |
| Delaware | Nebraska | 0.00089 | Delaware | Nevada | 0.00004 | Delaware | New Hampshire | 0.00328 |
| Delaware | New Jersey | 0.05006 | Delaware | New Mexico | 0.00230 | Delaware | New York | 0.08836 |
| Delaware | North Carolina | 0.01645 | Delaware | North Dakota | 0.00034 | Delaware | Ohio | 0.04482 |

| | | | | | | | | |
|---|---|---|---|---|---|---|---|---|
| Delaware | Oklahoma | 0.00298 | Delaware | Oregon | 0.00030 | Delaware | Pennsylvania | 0.26236 |
| Delaware | Rhode Island | 0.00064 | Delaware | South Carolina | 0.00341 | Delaware | South Dakota | 0.00209 |
| Delaware | Tennessee | 0.01308 | Delaware | Texas | 0.01257 | Delaware | Utah | 0.00158 |
| Delaware | Vermont | 0.02978 | Delaware | Virginia | 0.03553 | Delaware | Washington | 0.00209 |
| Delaware | West Virginia | 0.00665 | Delaware | Wisconsin | 0.03281 | Delaware | Wyoming | 0.00026 |
| Florida | Alabama | 0.00455 | Florida | Arizona | 0.00347 | Florida | Arkansas | 0.00149 |
| Florida | California | 0.00646 | Florida | Colorado | 0.00187 | Florida | Connecticut | 0.00064 |
| Florida | Delaware | 0.00019 | Florida | Florida | 0.75673 | Florida | Georgia | 0.06313 |
| Florida | Idaho | 0.00204 | Florida | Illinois | 0.00407 | Florida | Indiana | 0.00997 |
| Florida | Iowa | 0.00337 | Florida | Kansas | 0.00224 | Florida | Kentucky | 0.01195 |
| Florida | Louisiana | 0.00159 | Florida | Maine | 0.00061 | Florida | Maryland | 0.00233 |
| Florida | Massachusetts | 0.00078 | Florida | Michigan | 0.00925 | Florida | Minnesota | 0.00737 |
| Florida | Mississippi | 0.00269 | Florida | Missouri | 0.00518 | Florida | Montana | 0.00024 |
| Florida | Nebraska | 0.00026 | Florida | Nevada | 0.00013 | Florida | New Hampshire | 0.00025 |
| Florida | New Jersey | 0.00050 | Florida | New Mexico | 0.00279 | Florida | New York | 0.00797 |
| Florida | North Carolina | 0.00597 | Florida | North Dakota | 0.00036 | Florida | Ohio | 0.01039 |
| Florida | Oklahoma | 0.00241 | Florida | Oregon | 0.00037 | Florida | Pennsylvania | 0.00892 |
| Florida | Rhode Island | 0.00004 | Florida | South Carolina | 0.00496 | Florida | South Dakota | 0.00101 |
| Florida | Tennessee | 0.01084 | Florida | Texas | 0.01637 | Florida | Utah | 0.00084 |
| Florida | Vermont | 0.00324 | Florida | Virginia | 0.00369 | Florida | Washington | 0.00145 |
| Florida | West Virginia | 0.00074 | Florida | Wisconsin | 0.01405 | Florida | Wyoming | 0.00025 |
| Georgia | Alabama | 0.01104 | Georgia | Arizona | 0.00508 | Georgia | Arkansas | 0.00216 |
| Georgia | California | 0.00828 | Georgia | Colorado | 0.00255 | Georgia | Connecticut | 0.00142 |
| Georgia | Delaware | 0.00044 | Georgia | Florida | 0.24152 | Georgia | Georgia | 0.33489 |
| Georgia | Idaho | 0.00345 | Georgia | Illinois | 0.00919 | Georgia | Indiana | 0.02697 |
| Georgia | Iowa | 0.00623 | Georgia | Kansas | 0.00369 | Georgia | Kentucky | 0.03537 |
| Georgia | Louisiana | 0.00247 | Georgia | Maine | 0.00064 | Georgia | Maryland | 0.00638 |
| Georgia | Massachusetts | 0.00166 | Georgia | Michigan | 0.02162 | Georgia | Minnesota | 0.01723 |
| Georgia | Mississippi | 0.00416 | Georgia | Missouri | 0.01150 | Georgia | Montana | 0.00033 |
| Georgia | Nebraska | 0.00035 | Georgia | Nevada | 0.00022 | Georgia | New Hampshire | 0.00033 |
| Georgia | New Jersey | 0.00098 | Georgia | New Mexico | 0.00465 | Georgia | New York | 0.01702 |
| Georgia | North Carolina | 0.01676 | Georgia | North Dakota | 0.00079 | Georgia | Ohio | 0.02899 |
| Georgia | Oklahoma | 0.00351 | Georgia | Oregon | 0.00053 | Georgia | Pennsylvania | 0.02226 |
| Georgia | Rhode Island | 0.00011 | Georgia | South Carolina | 0.02581 | Georgia | South Dakota | 0.00156 |
| Georgia | Tennessee | 0.04471 | Georgia | Texas | 0.02500 | Georgia | Utah | 0.00128 |
| Georgia | Vermont | 0.00612 | Georgia | Virginia | 0.01045 | Georgia | Washington | 0.00256 |
| Georgia | West Virginia | 0.00151 | Georgia | Wisconsin | 0.02573 | Georgia | Wyoming | 0.00049 |
| Idaho | Alabama | 0.00028 | Idaho | Arizona | 0.01921 | Idaho | Arkansas | 0.00070 |
| Idaho | California | 0.08287 | Idaho | Colorado | 0.01207 | Idaho | Connecticut | 0.00026 |
| Idaho | Delaware | 0.00005 | Idaho | Florida | 0.00488 | Idaho | Georgia | 0.00200 |
| Idaho | Idaho | 0.65847 | Idaho | Illinois | 0.00202 | Idaho | Indiana | 0.00424 |
| Idaho | Iowa | 0.00405 | Idaho | Kansas | 0.00427 | Idaho | Kentucky | 0.00292 |
| Idaho | Louisiana | 0.00033 | Idaho | Maine | 0.00025 | Idaho | Maryland | 0.00065 |
| Idaho | Massachusetts | 0.00029 | Idaho | Michigan | 0.00561 | Idaho | Minnesota | 0.01334 |
| Idaho | Mississippi | 0.00035 | Idaho | Missouri | 0.00358 | Idaho | Montana | 0.00659 |
| Idaho | Nebraska | 0.00076 | Idaho | Nevada | 0.00597 | Idaho | New Hampshire | 0.00009 |
| Idaho | New Jersey | 0.00015 | Idaho | New Mexico | 0.00797 | Idaho | New York | 0.00335 |
| Idaho | North Carolina | 0.00066 | Idaho | North Dakota | 0.00196 | Idaho | Ohio | 0.00369 |
| Idaho | Oklahoma | 0.00276 | Idaho | Oregon | 0.01159 | Idaho | Pennsylvania | 0.00281 |
| Idaho | Rhode Island | 0.00002 | Idaho | South Carolina | 0.00034 | Idaho | South Dakota | 0.00345 |
| Idaho | Tennessee | 0.00158 | Idaho | Texas | 0.01624 | Idaho | Utah | 0.05016 |
| Idaho | Vermont | 0.00159 | Idaho | Virginia | 0.00070 | Idaho | Washington | 0.03527 |
| Idaho | West Virginia | 0.00015 | Idaho | Wisconsin | 0.01165 | Idaho | Wyoming | 0.00781 |
| Illinois | Alabama | 0.00147 | Illinois | Arizona | 0.00514 | Illinois | Arkansas | 0.00433 |
| Illinois | California | 0.01073 | Illinois | Colorado | 0.00475 | Illinois | Connecticut | 0.00063 |
| Illinois | Delaware | 0.00018 | Illinois | Florida | 0.01611 | Illinois | Georgia | 0.00956 |
| Illinois | Idaho | 0.00379 | Illinois | Illinois | 0.17702 | Illinois | Indiana | 0.09159 |
| Illinois | Iowa | 0.05673 | Illinois | Kansas | 0.00859 | Illinois | Kentucky | 0.03916 |
| Illinois | Louisiana | 0.00095 | Illinois | Maine | 0.00092 | Illinois | Maryland | 0.00253 |
| Illinois | Massachusetts | 0.00092 | Illinois | Michigan | 0.05620 | Illinois | Minnesota | 0.05427 |
| Illinois | Mississippi | 0.00187 | Illinois | Missouri | 0.04074 | Illinois | Montana | 0.00064 |
| Illinois | Nebraska | 0.00108 | Illinois | Nevada | 0.00024 | Illinois | New Hampshire | 0.00039 |
| Illinois | New Jersey | 0.00059 | Illinois | New Mexico | 0.00424 | Illinois | New York | 0.01291 |
| Illinois | North Carolina | 0.00378 | Illinois | North Dakota | 0.00107 | Illinois | Ohio | 0.03492 |
| Illinois | Oklahoma | 0.00569 | Illinois | Oregon | 0.00067 | Illinois | Pennsylvania | 0.01242 |
| Illinois | Rhode Island | 0.00006 | Illinois | South Carolina | 0.00166 | Illinois | South Dakota | 0.00446 |
| Illinois | Tennessee | 0.01790 | Illinois | Texas | 0.02094 | Illinois | Utah | 0.00175 |
| Illinois | Vermont | 0.00494 | Illinois | Virginia | 0.00361 | Illinois | Washington | 0.00259 |
| Illinois | West Virginia | 0.00110 | Illinois | Wisconsin | 0.27363 | Illinois | Wyoming | 0.00056 |
| Indiana | Alabama | 0.00096 | Indiana | Arizona | 0.00251 | Indiana | Arkansas | 0.00166 |
| Indiana | California | 0.00583 | Indiana | Colorado | 0.00218 | Indiana | Connecticut | 0.00092 |
| Indiana | Delaware | 0.00023 | Indiana | Florida | 0.01109 | Indiana | Georgia | 0.00786 |
| Indiana | Idaho | 0.00219 | Indiana | Illinois | 0.03072 | Indiana | Indiana | 0.42392 |
| Indiana | Iowa | 0.01332 | Indiana | Kansas | 0.00347 | Indiana | Kentucky | 0.04745 |
| Indiana | Louisiana | 0.00056 | Indiana | Maine | 0.00072 | Indiana | Maryland | 0.00341 |
| Indiana | Massachusetts | 0.00092 | Indiana | Michigan | 0.12300 | Indiana | Minnesota | 0.02224 |
| Indiana | Mississippi | 0.00118 | Indiana | Missouri | 0.01159 | Indiana | Montana | 0.00034 |
| Indiana | Nebraska | 0.00044 | Indiana | Nevada | 0.00011 | Indiana | New Hampshire | 0.00030 |
| Indiana | New Jersey | 0.00059 | Indiana | New Mexico | 0.00227 | Indiana | New York | 0.01585 |
| Indiana | North Carolina | 0.00349 | Indiana | North Dakota | 0.00069 | Indiana | Ohio | 0.12465 |
| Indiana | Oklahoma | 0.00243 | Indiana | Oregon | 0.00032 | Indiana | Pennsylvania | 0.01788 |
| Indiana | Rhode Island | 0.00004 | Indiana | South Carolina | 0.00144 | Indiana | South Dakota | 0.00221 |
| Indiana | Tennessee | 0.01167 | Indiana | Texas | 0.01104 | Indiana | Utah | 0.00083 |
| Indiana | Vermont | 0.00439 | Indiana | Virginia | 0.00436 | Indiana | Washington | 0.00139 |
| Indiana | West Virginia | 0.00131 | Indiana | Wisconsin | 0.07369 | Indiana | Wyoming | 0.00033 |
| Iowa | Alabama | 0.00011 | Iowa | Arizona | 0.00071 | Iowa | Arkansas | 0.00065 |
| Iowa | California | 0.00146 | Iowa | Colorado | 0.00193 | Iowa | Connecticut | 0.00010 |
| Iowa | Delaware | 0.00002 | Iowa | Florida | 0.00117 | Iowa | Georgia | 0.00087 |
| Iowa | Idaho | 0.00075 | Iowa | Illinois | 0.03753 | Iowa | Indiana | 0.00982 |

| | | | | | | | | |
|---|---|---|---|---|---|---|---|---|
| Iowa | Iowa | 0.37189 | Iowa | Kansas | 0.00745 | Iowa | Kentucky | 0.00340 |
| Iowa | Louisiana | 0.00013 | Iowa | Maine | 0.00008 | Iowa | Maryland | 0.00028 |
| Iowa | Massachusetts | 0.00007 | Iowa | Michigan | 0.01105 | Iowa | Minnesota | 0.23169 |
| Iowa | Mississippi | 0.00019 | Iowa | Missouri | 0.03080 | Iowa | Montana | 0.00017 |
| Iowa | Nebraska | 0.00510 | Iowa | Nevada | 0.00003 | Iowa | New Hampshire | 0.00003 |
| Iowa | New Jersey | 0.00006 | Iowa | New Mexico | 0.00090 | Iowa | New York | 0.00155 |
| Iowa | North Carolina | 0.00028 | Iowa | North Dakota | 0.00094 | Iowa | Ohio | 0.00400 |
| Iowa | Oklahoma | 0.00152 | Iowa | Oregon | 0.00009 | Iowa | Pennsylvania | 0.00126 |
| Iowa | Rhode Island | 0.00001 | Iowa | South Carolina | 0.00014 | Iowa | South Dakota | 0.02679 |
| Iowa | Tennessee | 0.00115 | Iowa | Texas | 0.00389 | Iowa | Utah | 0.00031 |
| Iowa | Vermont | 0.00049 | Iowa | Virginia | 0.00036 | Iowa | Washington | 0.00033 |
| Iowa | West Virginia | 0.00007 | Iowa | Wisconsin | 0.23810 | Iowa | Wyoming | 0.00027 |
| Kansas | Alabama | 0.00121 | Kansas | Arizona | 0.02199 | Kansas | Arkansas | 0.00717 |
| Kansas | California | 0.03250 | Kansas | Colorado | 0.05122 | Kansas | Connecticut | 0.00067 |
| Kansas | Delaware | 0.00018 | Kansas | Florida | 0.01844 | Kansas | Georgia | 0.00787 |
| Kansas | Idaho | 0.01576 | Kansas | Illinois | 0.01498 | Kansas | Indiana | 0.02333 |
| Kansas | Iowa | 0.03381 | Kansas | Kansas | 0.22898 | Kansas | Kentucky | 0.01474 |
| Kansas | Louisiana | 0.00183 | Kansas | Maine | 0.00066 | Kansas | Maryland | 0.00243 |
| Kansas | Massachusetts | 0.00091 | Kansas | Michigan | 0.02467 | Kansas | Minnesota | 0.06435 |
| Kansas | Mississippi | 0.00212 | Kansas | Missouri | 0.05775 | Kansas | Montana | 0.00170 |
| Kansas | Nebraska | 0.00953 | Kansas | Nevada | 0.00081 | Kansas | New Hampshire | 0.00032 |
| Kansas | New Jersey | 0.00050 | Kansas | New Mexico | 0.03415 | Kansas | New York | 0.01104 |
| Kansas | North Carolina | 0.00244 | Kansas | North Dakota | 0.00358 | Kansas | Ohio | 0.01644 |
| Kansas | Oklahoma | 0.04931 | Kansas | Oregon | 0.00184 | Kansas | Pennsylvania | 0.00999 |
| Kansas | Rhode Island | 0.00004 | Kansas | South Carolina | 0.00135 | Kansas | South Dakota | 0.01332 |
| Kansas | Tennessee | 0.00825 | Kansas | Texas | 0.11935 | Kansas | Utah | 0.00674 |
| Kansas | Vermont | 0.00448 | Kansas | Virginia | 0.00258 | Kansas | Washington | 0.00735 |
| Kansas | West Virginia | 0.00059 | Kansas | Wisconsin | 0.06311 | Kansas | Wyoming | 0.00365 |
| Kentucky | Alabama | 0.00336 | Kentucky | Arizona | 0.00425 | Kentucky | Arkansas | 0.00421 |
| Kentucky | California | 0.00947 | Kentucky | Colorado | 0.00350 | Kentucky | Connecticut | 0.00102 |
| Kentucky | Delaware | 0.00030 | Kentucky | Florida | 0.02649 | Kentucky | Georgia | 0.02635 |
| Kentucky | Idaho | 0.00281 | Kentucky | Illinois | 0.02509 | Kentucky | Indiana | 0.08919 |
| Kentucky | Iowa | 0.01107 | Kentucky | Kansas | 0.00493 | Kentucky | Kentucky | 0.40236 |
| Kentucky | Louisiana | 0.00113 | Kentucky | Maine | 0.00129 | Kentucky | Maryland | 0.00456 |
| Kentucky | Massachusetts | 0.00101 | Kentucky | Michigan | 0.03811 | Kentucky | Minnesota | 0.01621 |
| Kentucky | Mississippi | 0.00326 | Kentucky | Missouri | 0.01621 | Kentucky | Montana | 0.00041 |
| Kentucky | Nebraska | 0.00064 | Kentucky | Nevada | 0.00016 | Kentucky | New Hampshire | 0.00055 |
| Kentucky | New Jersey | 0.00081 | Kentucky | New Mexico | 0.00347 | Kentucky | New York | 0.01721 |
| Kentucky | North Carolina | 0.01222 | Kentucky | North Dakota | 0.00056 | Kentucky | Ohio | 0.06572 |
| Kentucky | Oklahoma | 0.00424 | Kentucky | Oregon | 0.00048 | Kentucky | Pennsylvania | 0.02013 |
| Kentucky | Rhode Island | 0.00007 | Kentucky | South Carolina | 0.00457 | Kentucky | South Dakota | 0.00257 |
| Kentucky | Tennessee | 0.08185 | Kentucky | Texas | 0.01882 | Kentucky | Utah | 0.00125 |
| Kentucky | Vermont | 0.00564 | Kentucky | Virginia | 0.01158 | Kentucky | Washington | 0.00176 |
| Kentucky | West Virginia | 0.00309 | Kentucky | Wisconsin | 0.04571 | Kentucky | Wyoming | 0.00034 |
| Louisiana | Alabama | 0.00687 | Louisiana | Arizona | 0.01252 | Louisiana | Arkansas | 0.02086 |
| Louisiana | California | 0.01996 | Louisiana | Colorado | 0.00663 | Louisiana | Connecticut | 0.00086 |
| Louisiana | Delaware | 0.00029 | Louisiana | Florida | 0.07810 | Louisiana | Georgia | 0.02888 |
| Louisiana | Idaho | 0.00589 | Louisiana | Illinois | 0.01309 | Louisiana | Indiana | 0.02299 |
| Louisiana | Iowa | 0.01203 | Louisiana | Kansas | 0.01039 | Louisiana | Kentucky | 0.02622 |
| Louisiana | Louisiana | 0.17944 | Louisiana | Maine | 0.00119 | Louisiana | Maryland | 0.00270 |
| Louisiana | Massachusetts | 0.00065 | Louisiana | Michigan | 0.02017 | Louisiana | Minnesota | 0.02262 |
| Louisiana | Mississippi | 0.05241 | Louisiana | Missouri | 0.02598 | Louisiana | Montana | 0.00057 |
| Louisiana | Nebraska | 0.00098 | Louisiana | Nevada | 0.00037 | Louisiana | New Hampshire | 0.00025 |
| Louisiana | New Jersey | 0.00049 | Louisiana | New Mexico | 0.01391 | Louisiana | New York | 0.01092 |
| Louisiana | North Carolina | 0.00565 | Louisiana | North Dakota | 0.00139 | Louisiana | Ohio | 0.01849 |
| Louisiana | Oklahoma | 0.01804 | Louisiana | Oregon | 0.00102 | Louisiana | Pennsylvania | 0.01170 |
| Louisiana | Rhode Island | 0.00000 | Louisiana | South Carolina | 0.00360 | Louisiana | South Dakota | 0.00290 |
| Louisiana | Tennessee | 0.02295 | Louisiana | Texas | 0.26232 | Louisiana | Utah | 0.00241 |
| Louisiana | Vermont | 0.00507 | Louisiana | Virginia | 0.00413 | Louisiana | Washington | 0.00344 |
| Louisiana | West Virginia | 0.00078 | Louisiana | Wisconsin | 0.03723 | Louisiana | Wyoming | 0.00061 |
| Maine | Alabama | 0.00067 | Maine | Arizona | 0.00406 | Maine | Arkansas | 0.00121 |
| Maine | California | 0.00961 | Maine | Colorado | 0.00267 | Maine | Connecticut | 0.01204 |
| Maine | Delaware | 0.00072 | Maine | Florida | 0.01931 | Maine | Georgia | 0.00779 |
| Maine | Idaho | 0.00326 | Maine | Illinois | 0.00645 | Maine | Indiana | 0.01659 |
| Maine | Iowa | 0.00598 | Maine | Kansas | 0.00292 | Maine | Kentucky | 0.00965 |
| Maine | Louisiana | 0.00067 | Maine | Maine | 0.28449 | Maine | Maryland | 0.00833 |
| Maine | Massachusetts | 0.03190 | Maine | Michigan | 0.02820 | Maine | Minnesota | 0.01530 |
| Maine | Mississippi | 0.00091 | Maine | Missouri | 0.00528 | Maine | Montana | 0.00037 |
| Maine | Nebraska | 0.00037 | Maine | Nevada | 0.00026 | Maine | New Hampshire | 0.02566 |
| Maine | New Jersey | 0.00437 | Maine | New Mexico | 0.00314 | Maine | New York | 0.10734 |
| Maine | North Carolina | 0.00629 | Maine | North Dakota | 0.00054 | Maine | Ohio | 0.02319 |
| Maine | Oklahoma | 0.00256 | Maine | Oregon | 0.00083 | Maine | Pennsylvania | 0.04359 |
| Maine | Rhode Island | 0.00186 | Maine | South Carolina | 0.00189 | Maine | South Dakota | 0.00181 |
| Maine | Tennessee | 0.00787 | Maine | Texas | 0.01185 | Maine | Utah | 0.00168 |
| Maine | Vermont | 0.22213 | Maine | Virginia | 0.00627 | Maine | Washington | 0.00277 |
| Maine | West Virginia | 0.00199 | Maine | Wisconsin | 0.04308 | Maine | Wyoming | 0.00031 |
| Maryland | Alabama | 0.00103 | Maryland | Arizona | 0.00294 | Maryland | Arkansas | 0.00089 |
| Maryland | California | 0.00561 | Maryland | Colorado | 0.00215 | Maryland | Connecticut | 0.00456 |
| Maryland | Delaware | 0.00745 | Maryland | Florida | 0.02144 | Maryland | Georgia | 0.01085 |
| Maryland | Idaho | 0.00247 | Maryland | Illinois | 0.00592 | Maryland | Indiana | 0.02383 |
| Maryland | Iowa | 0.00466 | Maryland | Kansas | 0.00262 | Maryland | Kentucky | 0.01712 |
| Maryland | Louisiana | 0.00060 | Maryland | Maine | 0.00203 | Maryland | Maryland | 0.19199 |
| Maryland | Massachusetts | 0.00603 | Maryland | Michigan | 0.03353 | Maryland | Minnesota | 0.01454 |
| Maryland | Mississippi | 0.00091 | Maryland | Missouri | 0.00557 | Maryland | Montana | 0.00023 |
| Maryland | Nebraska | 0.00028 | Maryland | Nevada | 0.00018 | Maryland | New Hampshire | 0.00134 |
| Maryland | New Jersey | 0.01471 | Maryland | New Mexico | 0.00239 | Maryland | New York | 0.08053 |
| Maryland | North Carolina | 0.01087 | Maryland | North Dakota | 0.00065 | Maryland | Ohio | 0.06012 |
| Maryland | Oklahoma | 0.00208 | Maryland | Oregon | 0.00040 | Maryland | Pennsylvania | 0.32551 |
| Maryland | Rhode Island | 0.00028 | Maryland | South Carolina | 0.00335 | Maryland | South Dakota | 0.00149 |

| | | | | | | | | |
|---|---|---|---|---|---|---|---|---|
| Maryland | Tennessee | 0.00951 | Maryland | Texas | 0.01082 | Maryland | Utah | 0.00089 |
| Maryland | Vermont | 0.01992 | Maryland | Virginia | 0.04174 | Maryland | Washington | 0.00185 |
| Maryland | West Virginia | 0.01275 | Maryland | Wisconsin | 0.02894 | Maryland | Wyoming | 0.00043 |
| Massachusetts | Alabama | 0.00063 | Massachusetts | Arizona | 0.00295 | Massachusetts | Arkansas | 0.00080 |
| Massachusetts | California | 0.00644 | Massachusetts | Colorado | 0.00195 | Massachusetts | Connecticut | 0.06606 |
| Massachusetts | Delaware | 0.00155 | Massachusetts | Florida | 0.01587 | Massachusetts | Georgia | 0.00622 |
| Massachusetts | Idaho | 0.00196 | Massachusetts | Illinois | 0.00473 | Massachusetts | Indiana | 0.01453 |
| Massachusetts | Iowa | 0.00417 | Massachusetts | Kansas | 0.00208 | Massachusetts | Kentucky | 0.00793 |
| Massachusetts | Louisiana | 0.00036 | Massachusetts | Maine | 0.02027 | Massachusetts | Maryland | 0.01253 |
| Massachusetts | Massachusetts | 0.19167 | Massachusetts | Michigan | 0.02490 | Massachusetts | Minnesota | 0.01198 |
| Massachusetts | Mississippi | 0.00077 | Massachusetts | Missouri | 0.00380 | Massachusetts | Montana | 0.00022 |
| Massachusetts | Nebraska | 0.00019 | Massachusetts | Nevada | 0.00027 | Massachusetts | New Hampshire | 0.03063 |
| Massachusetts | New Jersey | 0.01019 | Massachusetts | New Mexico | 0.00224 | Massachusetts | New York | 0.20340 |
| Massachusetts | North Carolina | 0.00507 | Massachusetts | North Dakota | 0.00046 | Massachusetts | Ohio | 0.02408 |
| Massachusetts | Oklahoma | 0.00175 | Massachusetts | Oregon | 0.00063 | Massachusetts | Pennsylvania | 0.06787 |
| Massachusetts | Rhode Island | 0.01087 | Massachusetts | South Carolina | 0.00145 | Massachusetts | South Dakota | 0.00116 |
| Massachusetts | Tennessee | 0.00519 | Massachusetts | Texas | 0.00894 | Massachusetts | Utah | 0.00092 |
| Massachusetts | Vermont | 0.17847 | Massachusetts | Virginia | 0.00712 | Massachusetts | Washington | 0.00221 |
| Massachusetts | West Virginia | 0.00188 | Massachusetts | Wisconsin | 0.03040 | Massachusetts | Wyoming | 0.00024 |
| Michigan | Alabama | 0.00036 | Michigan | Arizona | 0.00125 | Michigan | Arkansas | 0.00077 |
| Michigan | California | 0.00298 | Michigan | Colorado | 0.00139 | Michigan | Connecticut | 0.00059 |
| Michigan | Delaware | 0.00012 | Michigan | Florida | 0.00470 | Michigan | Georgia | 0.00330 |
| Michigan | Idaho | 0.00101 | Michigan | Illinois | 0.00923 | Michigan | Indiana | 0.08854 |
| Michigan | Iowa | 0.00849 | Michigan | Kansas | 0.00184 | Michigan | Kentucky | 0.01138 |
| Michigan | Louisiana | 0.00021 | Michigan | Maine | 0.00073 | Michigan | Maryland | 0.00205 |
| Michigan | Massachusetts | 0.00061 | Michigan | Michigan | 0.63507 | Michigan | Minnesota | 0.01499 |
| Michigan | Mississippi | 0.00043 | Michigan | Missouri | 0.00396 | Michigan | Montana | 0.00024 |
| Michigan | Nebraska | 0.00033 | Michigan | Nevada | 0.00006 | Michigan | New Hampshire | 0.00030 |
| Michigan | New Jersey | 0.00044 | Michigan | New Mexico | 0.00105 | Michigan | New York | 0.01564 |
| Michigan | North Carolina | 0.00193 | Michigan | North Dakota | 0.00040 | Michigan | Ohio | 0.05489 |
| Michigan | Oklahoma | 0.00119 | Michigan | Oregon | 0.00020 | Michigan | Pennsylvania | 0.01260 |
| Michigan | Rhode Island | 0.00003 | Michigan | South Carolina | 0.00061 | Michigan | South Dakota | 0.00151 |
| Michigan | Tennessee | 0.00418 | Michigan | Texas | 0.00475 | Michigan | Utah | 0.00050 |
| Michigan | Vermont | 0.00395 | Michigan | Virginia | 0.00228 | Michigan | Washington | 0.00070 |
| Michigan | West Virginia | 0.00084 | Michigan | Wisconsin | 0.09721 | Michigan | Wyoming | 0.00015 |
| Minnesota | Alabama | 0.00017 | Minnesota | Arizona | 0.00138 | Minnesota | Arkansas | 0.00057 |
| Minnesota | California | 0.00314 | Minnesota | Colorado | 0.00224 | Minnesota | Connecticut | 0.00015 |
| Minnesota | Delaware | 0.00003 | Minnesota | Florida | 0.00201 | Minnesota | Georgia | 0.00118 |
| Minnesota | Idaho | 0.00144 | Minnesota | Illinois | 0.00630 | Minnesota | Indiana | 0.00719 |
| Minnesota | Iowa | 0.05123 | Minnesota | Kansas | 0.00305 | Minnesota | Kentucky | 0.00292 |
| Minnesota | Louisiana | 0.00015 | Minnesota | Maine | 0.00020 | Minnesota | Maryland | 0.00042 |
| Minnesota | Massachusetts | 0.00017 | Minnesota | Michigan | 0.01086 | Minnesota | Minnesota | 0.68629 |
| Minnesota | Mississippi | 0.00022 | Minnesota | Missouri | 0.00504 | Minnesota | Montana | 0.00040 |
| Minnesota | Nebraska | 0.00107 | Minnesota | Nevada | 0.00007 | Minnesota | New Hampshire | 0.00007 |
| Minnesota | New Jersey | 0.00009 | Minnesota | New Mexico | 0.00125 | Minnesota | New York | 0.00248 |
| Minnesota | North Carolina | 0.00048 | Minnesota | North Dakota | 0.00253 | Minnesota | Ohio | 0.00434 |
| Minnesota | Oklahoma | 0.00126 | Minnesota | Oregon | 0.00021 | Minnesota | Pennsylvania | 0.00214 |
| Minnesota | Rhode Island | 0.00001 | Minnesota | South Carolina | 0.00021 | Minnesota | South Dakota | 0.01625 |
| Minnesota | Tennessee | 0.00136 | Minnesota | Texas | 0.00453 | Minnesota | Utah | 0.00066 |
| Minnesota | Vermont | 0.00097 | Minnesota | Virginia | 0.00049 | Minnesota | Washington | 0.00090 |
| Minnesota | West Virginia | 0.00014 | Minnesota | Wisconsin | 0.17143 | Minnesota | Wyoming | 0.00030 |
| Mississippi | Alabama | 0.02160 | Mississippi | Arizona | 0.01096 | Mississippi | Arkansas | 0.02002 |
| Mississippi | California | 0.01905 | Mississippi | Colorado | 0.00745 | Mississippi | Connecticut | 0.00086 |
| Mississippi | Delaware | 0.00019 | Mississippi | Florida | 0.10605 | Mississippi | Georgia | 0.05320 |
| Mississippi | Idaho | 0.00598 | Mississippi | Illinois | 0.02264 | Mississippi | Indiana | 0.03596 |
| Mississippi | Iowa | 0.01471 | Mississippi | Kansas | 0.01137 | Mississippi | Kentucky | 0.05691 |
| Mississippi | Louisiana | 0.03454 | Mississippi | Maine | 0.00160 | Mississippi | Maryland | 0.00388 |
| Mississippi | Massachusetts | 0.00123 | Mississippi | Michigan | 0.02663 | Mississippi | Minnesota | 0.02353 |
| Mississippi | Mississippi | 0.13221 | Mississippi | Missouri | 0.03590 | Mississippi | Montana | 0.00057 |
| Mississippi | Nebraska | 0.00097 | Mississippi | Nevada | 0.00019 | Mississippi | New Hampshire | 0.00063 |
| Mississippi | New Jersey | 0.00084 | Mississippi | New Mexico | 0.01053 | Mississippi | New York | 0.01581 |
| Mississippi | North Carolina | 0.01064 | Mississippi | North Dakota | 0.00084 | Mississippi | Ohio | 0.02728 |
| Mississippi | Oklahoma | 0.01450 | Mississippi | Oregon | 0.00118 | Mississippi | Pennsylvania | 0.01628 |
| Mississippi | Rhode Island | 0.00011 | Mississippi | South Carolina | 0.00532 | Mississippi | South Dakota | 0.00390 |
| Mississippi | Tennessee | 0.07695 | Mississippi | Texas | 0.08958 | Mississippi | Utah | 0.00312 |
| Mississippi | Vermont | 0.00645 | Mississippi | Virginia | 0.00653 | Mississippi | Washington | 0.00362 |
| Mississippi | West Virginia | 0.00188 | Mississippi | Wisconsin | 0.05515 | Mississippi | Wyoming | 0.00066 |
| Missouri | Alabama | 0.00188 | Missouri | Arizona | 0.00891 | Missouri | Arkansas | 0.01849 |
| Missouri | California | 0.01353 | Missouri | Colorado | 0.00753 | Missouri | Connecticut | 0.00090 |
| Missouri | Delaware | 0.00020 | Missouri | Florida | 0.02081 | Missouri | Georgia | 0.00969 |
| Missouri | Idaho | 0.00703 | Missouri | Illinois | 0.04949 | Missouri | Indiana | 0.04469 |
| Missouri | Iowa | 0.05134 | Missouri | Kansas | 0.03507 | Missouri | Kentucky | 0.02852 |
| Missouri | Louisiana | 0.00210 | Missouri | Maine | 0.00047 | Missouri | Maryland | 0.00335 |
| Missouri | Massachusetts | 0.00107 | Missouri | Michigan | 0.03349 | Missouri | Minnesota | 0.06493 |
| Missouri | Mississippi | 0.00316 | Missouri | Missouri | 0.32199 | Missouri | Montana | 0.00062 |
| Missouri | Nebraska | 0.00229 | Missouri | Nevada | 0.00037 | Missouri | New Hampshire | 0.00016 |
| Missouri | New Jersey | 0.00053 | Missouri | New Mexico | 0.01050 | Missouri | New York | 0.01283 |
| Missouri | North Carolina | 0.00269 | Missouri | North Dakota | 0.00224 | Missouri | Ohio | 0.02556 |
| Missouri | Oklahoma | 0.02240 | Missouri | Oregon | 0.00081 | Missouri | Pennsylvania | 0.01343 |
| Missouri | Rhode Island | 0.00008 | Missouri | South Carolina | 0.00155 | Missouri | South Dakota | 0.00592 |
| Missouri | Tennessee | 0.01518 | Missouri | Texas | 0.05621 | Missouri | Utah | 0.00244 |
| Missouri | Vermont | 0.00488 | Missouri | Virginia | 0.00316 | Missouri | Washington | 0.00422 |
| Missouri | West Virginia | 0.00071 | Missouri | Wisconsin | 0.08138 | Missouri | Wyoming | 0.00123 |
| Montana | Alabama | 0.00070 | Montana | Arizona | 0.02934 | Montana | Arkansas | 0.00189 |
| Montana | California | 0.10779 | Montana | Colorado | 0.03150 | Montana | Connecticut | 0.00092 |
| Montana | Delaware | 0.00017 | Montana | Florida | 0.01217 | Montana | Georgia | 0.00493 |
| Montana | Idaho | 0.18510 | Montana | Illinois | 0.00693 | Montana | Indiana | 0.01498 |
| Montana | Iowa | 0.01590 | Montana | Kansas | 0.01110 | Montana | Kentucky | 0.00835 |
| Montana | Louisiana | 0.00083 | Montana | Maine | 0.00084 | Montana | Maryland | 0.00179 |

| | | | | | | | | |
|---|---|---|---|---|---|---|---|---|
| Montana | Massachusetts | 0.00082 | Montana | Michigan | 0.02057 | Montana | Minnesota | 0.06205 |
| Montana | Mississippi | 0.00109 | Montana | Missouri | 0.01025 | Montana | Montana | 0.12074 |
| Montana | Nebraska | 0.00241 | Montana | Nevada | 0.00341 | Montana | New Hampshire | 0.00030 |
| Montana | New Jersey | 0.00037 | Montana | New Mexico | 0.01631 | Montana | New York | 0.01131 |
| Montana | North Carolina | 0.00189 | Montana | North Dakota | 0.01696 | Montana | Ohio | 0.01165 |
| Montana | Oklahoma | 0.00589 | Montana | Oregon | 0.01375 | Montana | Pennsylvania | 0.00924 |
| Montana | Rhode Island | 0.00005 | Montana | South Carolina | 0.00086 | Montana | South Dakota | 0.01579 |
| Montana | Tennessee | 0.00406 | Montana | Texas | 0.03579 | Montana | Utah | 0.03523 |
| Montana | Vermont | 0.00515 | Montana | Virginia | 0.00192 | Montana | Washington | 0.08186 |
| Montana | West Virginia | 0.00041 | Montana | Wisconsin | 0.05017 | Montana | Wyoming | 0.02444 |
| Nebraska | Alabama | 0.00064 | Nebraska | Arizona | 0.01034 | Nebraska | Arkansas | 0.00299 |
| Nebraska | California | 0.01434 | Nebraska | Colorado | 0.04486 | Nebraska | Connecticut | 0.00027 |
| Nebraska | Delaware | 0.00011 | Nebraska | Florida | 0.00932 | Nebraska | Georgia | 0.00388 |
| Nebraska | Idaho | 0.01021 | Nebraska | Illinois | 0.01411 | Nebraska | Indiana | 0.01609 |
| Nebraska | Iowa | 0.11837 | Nebraska | Kansas | 0.12590 | Nebraska | Kentucky | 0.00830 |
| Nebraska | Louisiana | 0.00099 | Nebraska | Maine | 0.00047 | Nebraska | Maryland | 0.00123 |
| Nebraska | Massachusetts | 0.00050 | Nebraska | Michigan | 0.02079 | Nebraska | Minnesota | 0.13612 |
| Nebraska | Mississippi | 0.00132 | Nebraska | Missouri | 0.04338 | Nebraska | Montana | 0.00135 |
| Nebraska | Nebraska | 0.12299 | Nebraska | Nevada | 0.00028 | Nebraska | New Hampshire | 0.00019 |
| Nebraska | New Jersey | 0.00030 | Nebraska | New Mexico | 0.01271 | Nebraska | New York | 0.00649 |
| Nebraska | North Carolina | 0.00138 | Nebraska | North Dakota | 0.00434 | Nebraska | Ohio | 0.01059 |
| Nebraska | Oklahoma | 0.01577 | Nebraska | Oregon | 0.00119 | Nebraska | Pennsylvania | 0.00534 |
| Nebraska | Rhode Island | 0.00002 | Nebraska | South Carolina | 0.00072 | Nebraska | South Dakota | 0.07890 |
| Nebraska | Tennessee | 0.00583 | Nebraska | Texas | 0.04043 | Nebraska | Utah | 0.00473 |
| Nebraska | Vermont | 0.00251 | Nebraska | Virginia | 0.00138 | Nebraska | Washington | 0.00387 |
| Nebraska | West Virginia | 0.00038 | Nebraska | Wisconsin | 0.08841 | Nebraska | Wyoming | 0.00539 |
| Nevada | Alabama | 0.00020 | Nevada | Arizona | 0.03722 | Nevada | Arkansas | 0.00095 |
| Nevada | California | 0.52169 | Nevada | Colorado | 0.00933 | Nevada | Connecticut | 0.00033 |
| Nevada | Delaware | 0.00002 | Nevada | Florida | 0.00672 | Nevada | Georgia | 0.00257 |
| Nevada | Idaho | 0.12946 | Nevada | Illinois | 0.00305 | Nevada | Indiana | 0.00553 |
| Nevada | Iowa | 0.00447 | Nevada | Kansas | 0.00422 | Nevada | Kentucky | 0.00318 |
| Nevada | Louisiana | 0.00051 | Nevada | Maine | 0.00031 | Nevada | Maryland | 0.00093 |
| Nevada | Massachusetts | 0.00029 | Nevada | Michigan | 0.00663 | Nevada | Minnesota | 0.01495 |
| Nevada | Mississippi | 0.00046 | Nevada | Missouri | 0.00456 | Nevada | Montana | 0.00263 |
| Nevada | Nebraska | 0.00051 | Nevada | Nevada | 0.08463 | Nevada | New Hampshire | 0.00013 |
| Nevada | New Jersey | 0.00009 | Nevada | New Mexico | 0.01188 | Nevada | New York | 0.00402 |
| Nevada | North Carolina | 0.00071 | Nevada | North Dakota | 0.00146 | Nevada | Ohio | 0.00513 |
| Nevada | Oklahoma | 0.00257 | Nevada | Oregon | 0.01201 | Nevada | Pennsylvania | 0.00420 |
| Nevada | Rhode Island | 0.00000 | Nevada | South Carolina | 0.00038 | Nevada | South Dakota | 0.00263 |
| Nevada | Tennessee | 0.00168 | Nevada | Texas | 0.02021 | Nevada | Utah | 0.03414 |
| Nevada | Vermont | 0.00153 | Nevada | Virginia | 0.00091 | Nevada | Washington | 0.03195 |
| Nevada | West Virginia | 0.00024 | Nevada | Wisconsin | 0.01645 | Nevada | Wyoming | 0.00232 |
| New Hampshire | Alabama | 0.00036 | New Hampshire | Arizona | 0.00259 | New Hampshire | Arkansas | 0.00076 |
| New Hampshire | California | 0.00666 | New Hampshire | Colorado | 0.00183 | New Hampshire | Connecticut | 0.03347 |
| New Hampshire | Delaware | 0.00119 | New Hampshire | Florida | 0.01408 | New Hampshire | Georgia | 0.00689 |
| New Hampshire | Idaho | 0.00231 | New Hampshire | Illinois | 0.00480 | New Hampshire | Indiana | 0.01474 |
| New Hampshire | Iowa | 0.00412 | New Hampshire | Kansas | 0.00234 | New Hampshire | Kentucky | 0.00841 |
| New Hampshire | Louisiana | 0.00038 | New Hampshire | Maine | 0.03375 | New Hampshire | Maryland | 0.00958 |
| New Hampshire | Massachusetts | 0.08113 | New Hampshire | Michigan | 0.02387 | New Hampshire | Minnesota | 0.01225 |
| New Hampshire | Mississippi | 0.00058 | New Hampshire | Missouri | 0.00412 | New Hampshire | Montana | 0.00038 |
| New Hampshire | Nebraska | 0.00013 | New Hampshire | Nevada | 0.00010 | New Hampshire | New Hampshire | 0.05866 |
| New Hampshire | New Jersey | 0.00463 | New Hampshire | New Mexico | 0.00219 | New Hampshire | New York | 0.17369 |
| New Hampshire | North Carolina | 0.00371 | New Hampshire | North Dakota | 0.00089 | New Hampshire | Ohio | 0.02298 |
| New Hampshire | Oklahoma | 0.00135 | New Hampshire | Oregon | 0.00058 | New Hampshire | Pennsylvania | 0.05663 |
| New Hampshire | Rhode Island | 0.00272 | New Hampshire | South Carolina | 0.00170 | New Hampshire | South Dakota | 0.00165 |
| New Hampshire | Tennessee | 0.00485 | New Hampshire | Texas | 0.00933 | New Hampshire | Utah | 0.00081 |
| New Hampshire | Vermont | 0.34874 | New Hampshire | Virginia | 0.00516 | New Hampshire | Washington | 0.00208 |
| New Hampshire | West Virginia | 0.00097 | New Hampshire | Wisconsin | 0.02539 | New Hampshire | Wyoming | 0.00046 |
| New Jersey | Alabama | 0.00088 | New Jersey | Arizona | 0.00400 | New Jersey | Arkansas | 0.00134 |
| New Jersey | California | 0.00992 | New Jersey | Colorado | 0.00267 | New Jersey | Connecticut | 0.01958 |
| New Jersey | Delaware | 0.01185 | New Jersey | Florida | 0.02389 | New Jersey | Georgia | 0.01164 |
| New Jersey | Idaho | 0.00302 | New Jersey | Illinois | 0.00675 | New Jersey | Indiana | 0.02389 |
| New Jersey | Iowa | 0.00653 | New Jersey | Kansas | 0.00317 | New Jersey | Kentucky | 0.01699 |
| New Jersey | Louisiana | 0.00062 | New Jersey | Maine | 0.00580 | New Jersey | Maryland | 0.06884 |
| New Jersey | Massachusetts | 0.01627 | New Jersey | Michigan | 0.03536 | New Jersey | Minnesota | 0.01524 |
| New Jersey | Mississippi | 0.00101 | New Jersey | Missouri | 0.00564 | New Jersey | Montana | 0.00040 |
| New Jersey | Nebraska | 0.00040 | New Jersey | Nevada | 0.00025 | New Jersey | New Hampshire | 0.00449 |
| New Jersey | New Jersey | 0.06119 | New Jersey | New Mexico | 0.00298 | New Jersey | New York | 0.15017 |
| New Jersey | North Carolina | 0.01115 | New Jersey | North Dakota | 0.00052 | New Jersey | Ohio | 0.04364 |
| New Jersey | Oklahoma | 0.00213 | New Jersey | Oregon | 0.00066 | New Jersey | Pennsylvania | 0.29205 |
| New Jersey | Rhode Island | 0.00096 | New Jersey | South Carolina | 0.00307 | New Jersey | South Dakota | 0.00180 |
| New Jersey | Tennessee | 0.00875 | New Jersey | Texas | 0.01290 | New Jersey | Utah | 0.00138 |
| New Jersey | Vermont | 0.04417 | New Jersey | Virginia | 0.02006 | New Jersey | Washington | 0.00253 |
| New Jersey | West Virginia | 0.00456 | New Jersey | Wisconsin | 0.03465 | New Jersey | Wyoming | 0.00022 |
| New Mexico | Alabama | 0.00046 | New Mexico | Arizona | 0.04518 | New Mexico | Arkansas | 0.00180 |
| New Mexico | California | 0.03027 | New Mexico | Colorado | 0.02284 | New Mexico | Connecticut | 0.00018 |
| New Mexico | Delaware | 0.00005 | New Mexico | Florida | 0.00794 | New Mexico | Georgia | 0.00293 |
| New Mexico | Idaho | 0.00973 | New Mexico | Illinois | 0.00311 | New Mexico | Indiana | 0.00531 |
| New Mexico | Iowa | 0.00478 | New Mexico | Kansas | 0.01382 | New Mexico | Kentucky | 0.00401 |
| New Mexico | Louisiana | 0.00095 | New Mexico | Maine | 0.00025 | New Mexico | Maryland | 0.00069 |
| New Mexico | Massachusetts | 0.00033 | New Mexico | Michigan | 0.00607 | New Mexico | Minnesota | 0.01251 |
| New Mexico | Mississippi | 0.00082 | New Mexico | Missouri | 0.00660 | New Mexico | Montana | 0.00085 |
| New Mexico | Nebraska | 0.00097 | New Mexico | Nevada | 0.00072 | New Mexico | New Hampshire | 0.00010 |
| New Mexico | New Jersey | 0.00019 | New Mexico | New Mexico | 0.44666 | New Mexico | New York | 0.00352 |
| New Mexico | North Carolina | 0.00092 | New Mexico | North Dakota | 0.00096 | New Mexico | Ohio | 0.00442 |
| New Mexico | Oklahoma | 0.01461 | New Mexico | Oregon | 0.00120 | New Mexico | Pennsylvania | 0.00297 |
| New Mexico | Rhode Island | 0.00002 | New Mexico | South Carolina | 0.00049 | New Mexico | South Dakota | 0.00279 |
| New Mexico | Tennessee | 0.00298 | New Mexico | Texas | 0.30552 | New Mexico | Utah | 0.00614 |
| New Mexico | Vermont | 0.00141 | New Mexico | Virginia | 0.00082 | New Mexico | Washington | 0.00426 |

| | | | | | | | | |
|---|---|---|---|---|---|---|---|---|
| New Mexico | West Virginia | 0.00023 | New Mexico | Wisconsin | 0.01504 | New Mexico | Wyoming | 0.00154 |
| New York | Alabama | 0.00067 | New York | Arizona | 0.00284 | New York | Arkansas | 0.00101 |
| New York | California | 0.00664 | New York | Colorado | 0.00202 | New York | Connecticut | 0.01296 |
| New York | Delaware | 0.00100 | New York | Florida | 0.01286 | New York | Georgia | 0.00726 |
| New York | Idaho | 0.00220 | New York | Illinois | 0.00565 | New York | Indiana | 0.02204 |
| New York | Iowa | 0.00601 | New York | Kansas | 0.00229 | New York | Kentucky | 0.01304 |
| New York | Louisiana | 0.00043 | New York | Maine | 0.00712 | New York | Maryland | 0.01365 |
| New York | Massachusetts | 0.01459 | New York | Michigan | 0.04122 | New York | Minnesota | 0.01306 |
| New York | Mississippi | 0.00075 | New York | Missouri | 0.00476 | New York | Montana | 0.00040 |
| New York | Nebraska | 0.00037 | New York | Nevada | 0.00014 | New York | New Hampshire | 0.00586 |
| New York | New Jersey | 0.00527 | New York | New Mexico | 0.00212 | New York | New York | 0.46696 |
| New York | North Carolina | 0.00533 | New York | North Dakota | 0.00061 | New York | Ohio | 0.04386 |
| New York | Oklahoma | 0.00172 | New York | Oregon | 0.00044 | New York | Pennsylvania | 0.12017 |
| New York | Rhode Island | 0.00062 | New York | South Carolina | 0.00165 | New York | South Dakota | 0.00166 |
| New York | Tennessee | 0.00624 | New York | Texas | 0.00901 | New York | Utah | 0.00093 |
| New York | Vermont | 0.08684 | New York | Virginia | 0.00817 | New York | Washington | 0.00185 |
| New York | West Virginia | 0.00228 | New York | Wisconsin | 0.03319 | New York | Wyoming | 0.00028 |
| North Carolina | Alabama | 0.00338 | North Carolina | Arizona | 0.00407 | North Carolina | Arkansas | 0.00187 |
| North Carolina | California | 0.00840 | North Carolina | Colorado | 0.00287 | North Carolina | Connecticut | 0.00325 |
| North Carolina | Delaware | 0.00157 | North Carolina | Florida | 0.06292 | North Carolina | Georgia | 0.06649 |
| North Carolina | Idaho | 0.00335 | North Carolina | Illinois | 0.00977 | North Carolina | Indiana | 0.03870 |
| North Carolina | Iowa | 0.00755 | North Carolina | Kansas | 0.00357 | North Carolina | Kentucky | 0.05610 |
| North Carolina | Louisiana | 0.00128 | North Carolina | Maine | 0.00158 | North Carolina | Maryland | 0.02146 |
| North Carolina | Massachusetts | 0.00316 | North Carolina | Michigan | 0.03453 | North Carolina | Minnesota | 0.01772 |
| North Carolina | Mississippi | 0.00234 | North Carolina | Missouri | 0.00906 | North Carolina | Montana | 0.00033 |
| North Carolina | Nebraska | 0.00045 | North Carolina | Nevada | 0.00016 | North Carolina | New Hampshire | 0.00078 |
| North Carolina | New Jersey | 0.00270 | North Carolina | New Mexico | 0.00374 | North Carolina | New York | 0.03658 |
| North Carolina | North Carolina | 0.23235 | North Carolina | North Dakota | 0.00083 | North Carolina | Ohio | 0.05713 |
| North Carolina | Oklahoma | 0.00285 | North Carolina | Oregon | 0.00053 | North Carolina | Pennsylvania | 0.06184 |
| North Carolina | Rhode Island | 0.00017 | North Carolina | South Carolina | 0.03842 | North Carolina | South Dakota | 0.00189 |
| North Carolina | Tennessee | 0.05327 | North Carolina | Texas | 0.01845 | North Carolina | Utah | 0.00112 |
| North Carolina | Vermont | 0.01158 | North Carolina | Virginia | 0.07069 | North Carolina | Washington | 0.00229 |
| North Carolina | West Virginia | 0.00590 | North Carolina | Wisconsin | 0.03052 | North Carolina | Wyoming | 0.00046 |
| North Dakota | Alabama | 0.00082 | North Dakota | Arizona | 0.01825 | North Dakota | Arkansas | 0.00329 |
| North Dakota | California | 0.05667 | North Dakota | Colorado | 0.03040 | North Dakota | Connecticut | 0.00127 |
| North Dakota | Delaware | 0.00015 | North Dakota | Florida | 0.01405 | North Dakota | Georgia | 0.00770 |
| North Dakota | Idaho | 0.02628 | North Dakota | Illinois | 0.01184 | North Dakota | Indiana | 0.02324 |
| North Dakota | Iowa | 0.04437 | North Dakota | Kansas | 0.01723 | North Dakota | Kentucky | 0.01547 |
| North Dakota | Louisiana | 0.00097 | North Dakota | Maine | 0.00175 | North Dakota | Maryland | 0.00229 |
| North Dakota | Massachusetts | 0.00108 | North Dakota | Michigan | 0.03616 | North Dakota | Minnesota | 0.18448 |
| North Dakota | Mississippi | 0.00145 | North Dakota | Missouri | 0.01405 | North Dakota | Montana | 0.01421 |
| North Dakota | Nebraska | 0.00617 | North Dakota | Nevada | 0.00093 | North Dakota | New Hampshire | 0.00075 |
| North Dakota | New Jersey | 0.00050 | North Dakota | New Mexico | 0.01227 | North Dakota | New York | 0.01520 |
| North Dakota | North Carolina | 0.00293 | North Dakota | North Dakota | 0.09926 | North Dakota | Ohio | 0.01790 |
| North Dakota | Oklahoma | 0.00783 | North Dakota | Oregon | 0.00361 | North Dakota | Pennsylvania | 0.01211 |
| North Dakota | Rhode Island | 0.00004 | North Dakota | South Carolina | 0.00112 | North Dakota | South Dakota | 0.09206 |
| North Dakota | Tennessee | 0.00690 | North Dakota | Texas | 0.03675 | North Dakota | Utah | 0.01089 |
| North Dakota | Vermont | 0.00629 | North Dakota | Virginia | 0.00297 | North Dakota | Washington | 0.01504 |
| North Dakota | West Virginia | 0.00076 | North Dakota | Wisconsin | 0.11576 | North Dakota | Wyoming | 0.00450 |
| Ohio | Alabama | 0.00104 | Ohio | Arizona | 0.00320 | Ohio | Arkansas | 0.00152 |
| Ohio | California | 0.00721 | Ohio | Colorado | 0.00247 | Ohio | Connecticut | 0.00161 |
| Ohio | Delaware | 0.00043 | Ohio | Florida | 0.01525 | Ohio | Georgia | 0.01009 |
| Ohio | Idaho | 0.00254 | Ohio | Illinois | 0.01206 | Ohio | Indiana | 0.11935 |
| Ohio | Iowa | 0.00897 | Ohio | Kansas | 0.00313 | Ohio | Kentucky | 0.04079 |
| Ohio | Louisiana | 0.00060 | Ohio | Maine | 0.00141 | Ohio | Maryland | 0.00915 |
| Ohio | Massachusetts | 0.00176 | Ohio | Michigan | 0.08826 | Ohio | Minnesota | 0.01734 |
| Ohio | Mississippi | 0.00106 | Ohio | Missouri | 0.00758 | Ohio | Montana | 0.00041 |
| Ohio | Nebraska | 0.00047 | Ohio | Nevada | 0.00013 | Ohio | New Hampshire | 0.00064 |
| Ohio | New Jersey | 0.00137 | Ohio | New Mexico | 0.00254 | Ohio | New York | 0.03745 |
| Ohio | North Carolina | 0.00669 | Ohio | North Dakota | 0.00071 | Ohio | Ohio | 0.42866 |
| Ohio | Oklahoma | 0.00230 | Ohio | Oregon | 0.00048 | Ohio | Pennsylvania | 0.05739 |
| Ohio | Rhode Island | 0.00010 | Ohio | South Carolina | 0.00217 | Ohio | South Dakota | 0.00208 |
| Ohio | Tennessee | 0.01217 | Ohio | Texas | 0.01167 | Ohio | Utah | 0.00100 |
| Ohio | Vermont | 0.00870 | Ohio | Virginia | 0.01023 | Ohio | Washington | 0.00182 |
| Ohio | West Virginia | 0.00421 | Ohio | Wisconsin | 0.04940 | Ohio | Wyoming | 0.00038 |
| Oklahoma | Alabama | 0.00114 | Oklahoma | Arizona | 0.01525 | Oklahoma | Arkansas | 0.02092 |
| Oklahoma | California | 0.02208 | Oklahoma | Colorado | 0.02122 | Oklahoma | Connecticut | 0.00046 |
| Oklahoma | Delaware | 0.00011 | Oklahoma | Florida | 0.01406 | Oklahoma | Georgia | 0.00732 |
| Oklahoma | Idaho | 0.00781 | Oklahoma | Illinois | 0.01063 | Oklahoma | Indiana | 0.01596 |
| Oklahoma | Iowa | 0.01597 | Oklahoma | Kansas | 0.07604 | Oklahoma | Kentucky | 0.01253 |
| Oklahoma | Louisiana | 0.00284 | Oklahoma | Maine | 0.00043 | Oklahoma | Maryland | 0.00123 |
| Oklahoma | Massachusetts | 0.00049 | Oklahoma | Michigan | 0.01431 | Oklahoma | Minnesota | 0.02675 |
| Oklahoma | Mississippi | 0.00256 | Oklahoma | Missouri | 0.05296 | Oklahoma | Montana | 0.00085 |
| Oklahoma | Nebraska | 0.00235 | Oklahoma | Nevada | 0.00034 | Oklahoma | New Hampshire | 0.00014 |
| Oklahoma | New Jersey | 0.00022 | Oklahoma | New Mexico | 0.04122 | Oklahoma | New York | 0.00608 |
| Oklahoma | North Carolina | 0.00181 | Oklahoma | North Dakota | 0.00155 | Oklahoma | Ohio | 0.01075 |
| Oklahoma | Oklahoma | 0.23030 | Oklahoma | Oregon | 0.00092 | Oklahoma | Pennsylvania | 0.00649 |
| Oklahoma | Rhode Island | 0.00002 | Oklahoma | South Carolina | 0.00096 | Oklahoma | South Dakota | 0.00531 |
| Oklahoma | Tennessee | 0.00695 | Oklahoma | Texas | 0.29429 | Oklahoma | Utah | 0.00427 |
| Oklahoma | Vermont | 0.00235 | Oklahoma | Virginia | 0.00171 | Oklahoma | Washington | 0.00386 |
| Oklahoma | West Virginia | 0.00031 | Oklahoma | Wisconsin | 0.03275 | Oklahoma | Wyoming | 0.00114 |
| Oregon | Alabama | 0.00032 | Oregon | Arizona | 0.01804 | Oregon | Arkansas | 0.00079 |
| Oregon | California | 0.16506 | Oregon | Colorado | 0.00810 | Oregon | Connecticut | 0.00030 |
| Oregon | Delaware | 0.00005 | Oregon | Florida | 0.00546 | Oregon | Georgia | 0.00243 |
| Oregon | Idaho | 0.09050 | Oregon | Illinois | 0.00223 | Oregon | Indiana | 0.00467 |
| Oregon | Iowa | 0.00414 | Oregon | Kansas | 0.00362 | Oregon | Kentucky | 0.00319 |
| Oregon | Louisiana | 0.00032 | Oregon | Maine | 0.00044 | Oregon | Maryland | 0.00059 |
| Oregon | Massachusetts | 0.00033 | Oregon | Michigan | 0.00603 | Oregon | Minnesota | 0.01138 |
| Oregon | Mississippi | 0.00038 | Oregon | Missouri | 0.00352 | Oregon | Montana | 0.00464 |

| | | | | | | | | |
|---|---|---|---|---|---|---|---|---|
| Oregon | Nebraska | 0.00064 | Oregon | Nevada | 0.00479 | Oregon | New Hampshire | 0.00013 |
| Oregon | New Jersey | 0.00017 | Oregon | New Mexico | 0.00672 | Oregon | New York | 0.00372 |
| Oregon | North Carolina | 0.00095 | Oregon | North Dakota | 0.00131 | Oregon | Ohio | 0.00414 |
| Oregon | Oklahoma | 0.00231 | Oregon | Oregon | 0.32254 | Oregon | Pennsylvania | 0.00309 |
| Oregon | Rhode Island | 0.00002 | Oregon | South Carolina | 0.00039 | Oregon | South Dakota | 0.00275 |
| Oregon | Tennessee | 0.00179 | Oregon | Texas | 0.01445 | Oregon | Utah | 0.01335 |
| Oregon | Vermont | 0.00185 | Oregon | Virginia | 0.00079 | Oregon | Washington | 0.26108 |
| Oregon | West Virginia | 0.00018 | Oregon | Wisconsin | 0.01451 | Oregon | Wyoming | 0.00178 |
| Pennsylvania | Alabama | 0.00086 | Pennsylvania | Arizona | 0.00313 | Pennsylvania | Arkansas | 0.00103 |
| Pennsylvania | California | 0.00693 | Pennsylvania | Colorado | 0.00209 | Pennsylvania | Connecticut | 0.00583 |
| Pennsylvania | Delaware | 0.00345 | Pennsylvania | Florida | 0.01746 | Pennsylvania | Georgia | 0.00936 |
| Pennsylvania | Idaho | 0.00243 | Pennsylvania | Illinois | 0.00614 | Pennsylvania | Indiana | 0.02447 |
| Pennsylvania | Iowa | 0.00559 | Pennsylvania | Kansas | 0.00241 | Pennsylvania | Kentucky | 0.01564 |
| Pennsylvania | Louisiana | 0.00057 | Pennsylvania | Maine | 0.00287 | Pennsylvania | Maryland | 0.06982 |
| Pennsylvania | Massachusetts | 0.00685 | Pennsylvania | Michigan | 0.03615 | Pennsylvania | Minnesota | 0.01349 |
| Pennsylvania | Mississippi | 0.00087 | Pennsylvania | Missouri | 0.00543 | Pennsylvania | Montana | 0.00039 |
| Pennsylvania | Nebraska | 0.00036 | Pennsylvania | Nevada | 0.00016 | Pennsylvania | New Hampshire | 0.00167 |
| Pennsylvania | New Jersey | 0.01352 | Pennsylvania | New Mexico | 0.00239 | Pennsylvania | New York | 0.11661 |
| Pennsylvania | North Carolina | 0.00828 | Pennsylvania | North Dakota | 0.00064 | Pennsylvania | Ohio | 0.06572 |
| Pennsylvania | Oklahoma | 0.00192 | Pennsylvania | Oregon | 0.00046 | Pennsylvania | Pennsylvania | 0.43927 |
| Pennsylvania | Rhode Island | 0.00038 | Pennsylvania | South Carolina | 0.00248 | Pennsylvania | South Dakota | 0.00163 |
| Pennsylvania | Tennessee | 0.00827 | Pennsylvania | Texas | 0.01030 | Pennsylvania | Utah | 0.00101 |
| Pennsylvania | Vermont | 0.02278 | Pennsylvania | Virginia | 0.01878 | Pennsylvania | Washington | 0.00200 |
| Pennsylvania | West Virginia | 0.00610 | Pennsylvania | Wisconsin | 0.03169 | Pennsylvania | Wyoming | 0.00032 |
| Rhode Island | Alabama | 0.00064 | Rhode Island | Arizona | 0.00355 | Rhode Island | Arkansas | 0.00177 |
| Rhode Island | California | 0.01015 | Rhode Island | Colorado | 0.00419 | Rhode Island | Connecticut | 0.10171 |
| Rhode Island | Delaware | 0.00193 | Rhode Island | Florida | 0.02837 | Rhode Island | Georgia | 0.01273 |
| Rhode Island | Idaho | 0.00306 | Rhode Island | Illinois | 0.00790 | Rhode Island | Indiana | 0.01918 |
| Rhode Island | Iowa | 0.00548 | Rhode Island | Kansas | 0.00371 | Rhode Island | Kentucky | 0.01515 |
| Rhode Island | Louisiana | 0.00081 | Rhode Island | Maine | 0.02031 | Rhode Island | Maryland | 0.01676 |
| Rhode Island | Massachusetts | 0.13459 | Rhode Island | Michigan | 0.03498 | Rhode Island | Minnesota | 0.01934 |
| Rhode Island | Mississippi | 0.00097 | Rhode Island | Missouri | 0.00725 | Rhode Island | Montana | 0.00048 |
| Rhode Island | Nebraska | 0.00048 | Rhode Island | Nevada | 0.00081 | Rhode Island | New Hampshire | 0.01564 |
| Rhode Island | New Jersey | 0.01225 | Rhode Island | New Mexico | 0.00403 | Rhode Island | New York | 0.15861 |
| Rhode Island | North Carolina | 0.00774 | Rhode Island | North Dakota | 0.00097 | Rhode Island | Ohio | 0.03320 |
| Rhode Island | Oklahoma | 0.00419 | Rhode Island | Oregon | 0.00048 | Rhode Island | Pennsylvania | 0.08511 |
| Rhode Island | Rhode Island | 0.01451 | Rhode Island | South Carolina | 0.00274 | Rhode Island | South Dakota | 0.00419 |
| Rhode Island | Tennessee | 0.01048 | Rhode Island | Texas | 0.01354 | Rhode Island | Utah | 0.00145 |
| Rhode Island | Vermont | 0.11928 | Rhode Island | Virginia | 0.00774 | Rhode Island | Washington | 0.00387 |
| Rhode Island | West Virginia | 0.00242 | Rhode Island | Wisconsin | 0.04062 | Rhode Island | Wyoming | 0.00064 |
| South Carolina | Alabama | 0.00522 | South Carolina | Arizona | 0.00665 | South Carolina | Arkansas | 0.00288 |
| South Carolina | California | 0.01455 | South Carolina | Colorado | 0.00375 | South Carolina | Connecticut | 0.00234 |
| South Carolina | Delaware | 0.00083 | South Carolina | Florida | 0.12352 | South Carolina | Georgia | 0.17180 |
| South Carolina | Idaho | 0.00468 | South Carolina | Illinois | 0.01241 | South Carolina | Indiana | 0.03828 |
| South Carolina | Iowa | 0.00899 | South Carolina | Kansas | 0.00494 | South Carolina | Kentucky | 0.05233 |
| South Carolina | Louisiana | 0.00186 | South Carolina | Maine | 0.00207 | South Carolina | Maryland | 0.01071 |
| South Carolina | Massachusetts | 0.00258 | South Carolina | Michigan | 0.03272 | South Carolina | Minnesota | 0.01918 |
| South Carolina | Mississippi | 0.00366 | South Carolina | Missouri | 0.01180 | South Carolina | Montana | 0.00054 |
| South Carolina | Nebraska | 0.00060 | South Carolina | Nevada | 0.00025 | South Carolina | New Hampshire | 0.00094 |
| South Carolina | New Jersey | 0.00174 | South Carolina | New Mexico | 0.00538 | South Carolina | New York | 0.02956 |
| South Carolina | North Carolina | 0.07642 | South Carolina | North Dakota | 0.00088 | South Carolina | Ohio | 0.04454 |
| South Carolina | Oklahoma | 0.00405 | South Carolina | Oregon | 0.00069 | South Carolina | Pennsylvania | 0.03874 |
| South Carolina | Rhode Island | 0.00018 | South Carolina | South Carolina | 0.08500 | South Carolina | South Dakota | 0.00243 |
| South Carolina | Tennessee | 0.05608 | South Carolina | Texas | 0.02578 | South Carolina | Utah | 0.00181 |
| South Carolina | Vermont | 0.01103 | South Carolina | Virginia | 0.02537 | South Carolina | Washington | 0.00337 |
| South Carolina | West Virginia | 0.00413 | South Carolina | Wisconsin | 0.04233 | South Carolina | Wyoming | 0.00041 |
| South Dakota | Alabama | 0.00093 | South Dakota | Arizona | 0.01390 | South Dakota | Arkansas | 0.00251 |
| South Dakota | California | 0.03045 | South Dakota | Colorado | 0.02073 | South Dakota | Connecticut | 0.00080 |
| South Dakota | Delaware | 0.00021 | South Dakota | Florida | 0.01424 | South Dakota | Georgia | 0.00628 |
| South Dakota | Idaho | 0.01692 | South Dakota | Illinois | 0.01468 | South Dakota | Indiana | 0.02381 |
| South Dakota | Iowa | 0.08119 | South Dakota | Kansas | 0.01912 | South Dakota | Kentucky | 0.01073 |
| South Dakota | Louisiana | 0.00088 | South Dakota | Maine | 0.00085 | South Dakota | Maryland | 0.00239 |
| South Dakota | Massachusetts | 0.00138 | South Dakota | Michigan | 0.02991 | South Dakota | Minnesota | 0.25211 |
| South Dakota | Mississippi | 0.00119 | South Dakota | Missouri | 0.02110 | South Dakota | Montana | 0.00296 |
| South Dakota | Nebraska | 0.00981 | South Dakota | Nevada | 0.00093 | South Dakota | New Hampshire | 0.00035 |
| South Dakota | New Jersey | 0.00049 | South Dakota | New Mexico | 0.01242 | South Dakota | New York | 0.01292 |
| South Dakota | North Carolina | 0.00219 | South Dakota | North Dakota | 0.01648 | South Dakota | Ohio | 0.01806 |
| South Dakota | Oklahoma | 0.00775 | South Dakota | Oregon | 0.00211 | South Dakota | Pennsylvania | 0.01183 |
| South Dakota | Rhode Island | 0.00010 | South Dakota | South Carolina | 0.00095 | South Dakota | South Dakota | 0.14989 |
| South Dakota | Tennessee | 0.00607 | South Dakota | Texas | 0.03515 | South Dakota | Utah | 0.00648 |
| South Dakota | Vermont | 0.00539 | South Dakota | Virginia | 0.00237 | South Dakota | Washington | 0.01063 |
| South Dakota | West Virginia | 0.00057 | South Dakota | Wisconsin | 0.11430 | South Dakota | Wyoming | 0.00350 |
| Tennessee | Alabama | 0.00914 | Tennessee | Arizona | 0.00770 | Tennessee | Arkansas | 0.00663 |
| Tennessee | California | 0.01639 | Tennessee | Colorado | 0.00492 | Tennessee | Connecticut | 0.00171 |
| Tennessee | Delaware | 0.00047 | Tennessee | Florida | 0.05649 | Tennessee | Georgia | 0.06230 |
| Tennessee | Idaho | 0.00518 | Tennessee | Illinois | 0.02397 | Tennessee | Indiana | 0.05989 |
| Tennessee | Iowa | 0.01395 | Tennessee | Kansas | 0.00728 | Tennessee | Kentucky | 0.15326 |
| Tennessee | Louisiana | 0.00236 | Tennessee | Maine | 0.00149 | Tennessee | Maryland | 0.00670 |
| Tennessee | Massachusetts | 0.00179 | Tennessee | Michigan | 0.03854 | Tennessee | Minnesota | 0.02539 |
| Tennessee | Mississippi | 0.00744 | Tennessee | Missouri | 0.02343 | Tennessee | Montana | 0.00067 |
| Tennessee | Nebraska | 0.00079 | Tennessee | Nevada | 0.00032 | Tennessee | New Hampshire | 0.00061 |
| Tennessee | New Jersey | 0.00110 | Tennessee | New Mexico | 0.00666 | Tennessee | New York | 0.02317 |
| Tennessee | North Carolina | 0.02289 | Tennessee | North Dakota | 0.00104 | Tennessee | Ohio | 0.04987 |
| Tennessee | Oklahoma | 0.00633 | Tennessee | Oregon | 0.00082 | Tennessee | Pennsylvania | 0.02867 |
| Tennessee | Rhode Island | 0.00012 | Tennessee | South Carolina | 0.01008 | Tennessee | South Dakota | 0.00324 |
| Tennessee | Tennessee | 0.18518 | Tennessee | Texas | 0.03609 | Tennessee | Utah | 0.00211 |
| Tennessee | Vermont | 0.00792 | Tennessee | Virginia | 0.01681 | Tennessee | Washington | 0.00360 |
| Tennessee | West Virginia | 0.00286 | Tennessee | Wisconsin | 0.05198 | Tennessee | Wyoming | 0.00064 |
| Texas | Alabama | 0.00121 | Texas | Arizona | 0.02424 | Texas | Arkansas | 0.00627 |

| | | | | | | | | |
|---|---|---|---|---|---|---|---|---|
| Texas | California | 0.03248 | Texas | Colorado | 0.01781 | Texas | Connecticut | 0.00055 |
| Texas | Delaware | 0.00011 | Texas | Florida | 0.01905 | Texas | Georgia | 0.00782 |
| Texas | Idaho | 0.00952 | Texas | Illinois | 0.00665 | Texas | Indiana | 0.01215 |
| Texas | Iowa | 0.00943 | Texas | Kansas | 0.01878 | Texas | Kentucky | 0.01049 |
| Texas | Louisiana | 0.00386 | Texas | Maine | 0.00062 | Texas | Maryland | 0.00156 |
| Texas | Massachusetts | 0.00060 | Texas | Michigan | 0.01237 | Texas | Minnesota | 0.01936 |
| Texas | Mississippi | 0.00262 | Texas | Missouri | 0.01460 | Texas | Montana | 0.00112 |
| Texas | Nebraska | 0.00143 | Texas | Nevada | 0.00059 | Texas | New Hampshire | 0.00023 |
| Texas | New Jersey | 0.00032 | Texas | New Mexico | 0.10178 | Texas | New York | 0.00715 |
| Texas | North Carolina | 0.00214 | Texas | North Dakota | 0.00130 | Texas | Ohio | 0.00977 |
| Texas | Oklahoma | 0.02982 | Texas | Oregon | 0.00131 | Texas | Pennsylvania | 0.00679 |
| Texas | Rhode Island | 0.00004 | Texas | South Carolina | 0.00113 | Texas | South Dakota | 0.00407 |
| Texas | Tennessee | 0.00662 | Texas | Texas | 0.54949 | Texas | Utah | 0.00512 |
| Texas | Vermont | 0.00295 | Texas | Virginia | 0.00196 | Texas | Washington | 0.00508 |
| Texas | West Virginia | 0.00042 | Texas | Wisconsin | 0.02585 | Texas | Wyoming | 0.00133 |
| Utah | Alabama | 0.00054 | Utah | Arizona | 0.05857 | Utah | Arkansas | 0.00153 |
| Utah | California | 0.15770 | Utah | Colorado | 0.03357 | Utah | Connecticut | 0.00054 |
| Utah | Delaware | 0.00008 | Utah | Florida | 0.00985 | Utah | Georgia | 0.00403 |
| Utah | Idaho | 0.22273 | Utah | Illinois | 0.00446 | Utah | Indiana | 0.00904 |
| Utah | Iowa | 0.00858 | Utah | Kansas | 0.00946 | Utah | Kentucky | 0.00556 |
| Utah | Louisiana | 0.00085 | Utah | Maine | 0.00049 | Utah | Maryland | 0.00117 |
| Utah | Massachusetts | 0.00059 | Utah | Michigan | 0.01039 | Utah | Minnesota | 0.02311 |
| Utah | Mississippi | 0.00089 | Utah | Missouri | 0.00786 | Utah | Montana | 0.00664 |
| Utah | Nebraska | 0.00152 | Utah | Nevada | 0.00650 | Utah | New Hampshire | 0.00014 |
| Utah | New Jersey | 0.00024 | Utah | New Mexico | 0.02259 | Utah | New York | 0.00635 |
| Utah | North Carolina | 0.00134 | Utah | North Dakota | 0.00317 | Utah | Ohio | 0.00757 |
| Utah | Oklahoma | 0.00565 | Utah | Oregon | 0.00739 | Utah | Pennsylvania | 0.00572 |
| Utah | Rhode Island | 0.00003 | Utah | South Carolina | 0.00070 | Utah | South Dakota | 0.00597 |
| Utah | Tennessee | 0.00307 | Utah | Texas | 0.03743 | Utah | Utah | 0.24007 |
| Utah | Vermont | 0.00279 | Utah | Virginia | 0.00135 | Utah | Washington | 0.02651 |
| Utah | West Virginia | 0.00032 | Utah | Wisconsin | 0.02417 | Utah | Wyoming | 0.01116 |
| Vermont | Alabama | 0.00032 | Vermont | Arizona | 0.00137 | Vermont | Arkansas | 0.00049 |
| Vermont | California | 0.00349 | Vermont | Colorado | 0.00080 | Vermont | Connecticut | 0.01393 |
| Vermont | Delaware | 0.00040 | Vermont | Florida | 0.00583 | Vermont | Georgia | 0.00368 |
| Vermont | Idaho | 0.00106 | Vermont | Illinois | 0.00286 | Vermont | Indiana | 0.01044 |
| Vermont | Iowa | 0.00322 | Vermont | Kansas | 0.00100 | Vermont | Kentucky | 0.00525 |
| Vermont | Louisiana | 0.00026 | Vermont | Maine | 0.01420 | Vermont | Maryland | 0.00544 |
| Vermont | Massachusetts | 0.02240 | Vermont | Michigan | 0.01516 | Vermont | Minnesota | 0.00745 |
| Vermont | Mississippi | 0.00035 | Vermont | Missouri | 0.00273 | Vermont | Montana | 0.00023 |
| Vermont | Nebraska | 0.00020 | Vermont | Nevada | 0.00004 | Vermont | New Hampshire | 0.01624 |
| Vermont | New Jersey | 0.00193 | Vermont | New Mexico | 0.00120 | Vermont | New York | 0.15541 |
| Vermont | North Carolina | 0.00195 | Vermont | North Dakota | 0.00047 | Vermont | Ohio | 0.01497 |
| Vermont | Oklahoma | 0.00070 | Vermont | Oregon | 0.00022 | Vermont | Pennsylvania | 0.03687 |
| Vermont | Rhode Island | 0.00070 | Vermont | South Carolina | 0.00073 | Vermont | South Dakota | 0.00067 |
| Vermont | Tennessee | 0.00222 | Vermont | Texas | 0.00489 | Vermont | Utah | 0.00054 |
| Vermont | Vermont | 0.61956 | Vermont | Virginia | 0.00297 | Vermont | Washington | 0.00103 |
| Vermont | West Virginia | 0.00062 | Vermont | Wisconsin | 0.01338 | Vermont | Wyoming | 0.00013 |
| Virginia | Alabama | 0.00212 | Virginia | Arizona | 0.00421 | Virginia | Arkansas | 0.00173 |
| Virginia | California | 0.00989 | Virginia | Colorado | 0.00271 | Virginia | Connecticut | 0.00451 |
| Virginia | Delaware | 0.00240 | Virginia | Florida | 0.03698 | Virginia | Georgia | 0.03099 |
| Virginia | Idaho | 0.00339 | Virginia | Illinois | 0.01030 | Virginia | Indiana | 0.04496 |
| Virginia | Iowa | 0.00842 | Virginia | Kansas | 0.00344 | Virginia | Kentucky | 0.05517 |
| Virginia | Louisiana | 0.00102 | Virginia | Maine | 0.00209 | Virginia | Maryland | 0.05590 |
| Virginia | Massachusetts | 0.00389 | Virginia | Michigan | 0.04274 | Virginia | Minnesota | 0.01873 |
| Virginia | Mississippi | 0.00178 | Virginia | Missouri | 0.00933 | Virginia | Montana | 0.00051 |
| Virginia | Nebraska | 0.00047 | Virginia | Nevada | 0.00017 | Virginia | New Hampshire | 0.00108 |
| Virginia | New Jersey | 0.00411 | Virginia | New Mexico | 0.00378 | Virginia | New York | 0.05460 |
| Virginia | North Carolina | 0.05107 | Virginia | North Dakota | 0.00097 | Virginia | Ohio | 0.08349 |
| Virginia | Oklahoma | 0.00273 | Virginia | Oregon | 0.00053 | Virginia | Pennsylvania | 0.13734 |
| Virginia | Rhode Island | 0.00025 | Virginia | South Carolina | 0.00964 | Virginia | South Dakota | 0.00215 |
| Virginia | Tennessee | 0.03356 | Virginia | Texas | 0.01742 | Virginia | Utah | 0.00129 |
| Virginia | Vermont | 0.01429 | Virginia | Virginia | 0.16797 | Virginia | Washington | 0.00254 |
| Virginia | West Virginia | 0.01739 | Virginia | Wisconsin | 0.03545 | Virginia | Wyoming | 0.00047 |
| Washington | Alabama | 0.00028 | Washington | Arizona | 0.01241 | Washington | Arkansas | 0.00066 |
| Washington | California | 0.07346 | Washington | Colorado | 0.00647 | Washington | Connecticut | 0.00030 |
| Washington | Delaware | 0.00006 | Washington | Florida | 0.00490 | Washington | Georgia | 0.00206 |
| Washington | Idaho | 0.04815 | Washington | Illinois | 0.00214 | Washington | Indiana | 0.00441 |
| Washington | Iowa | 0.00389 | Washington | Kansas | 0.00312 | Washington | Kentucky | 0.00298 |
| Washington | Louisiana | 0.00032 | Washington | Maine | 0.00036 | Washington | Maryland | 0.00064 |
| Washington | Massachusetts | 0.00034 | Washington | Michigan | 0.00565 | Washington | Minnesota | 0.01112 |
| Washington | Mississippi | 0.00034 | Washington | Missouri | 0.00310 | Washington | Montana | 0.00489 |
| Washington | Nebraska | 0.00050 | Washington | Nevada | 0.00217 | Washington | New Hampshire | 0.00013 |
| Washington | New Jersey | 0.00016 | Washington | New Mexico | 0.00525 | Washington | New York | 0.00351 |
| Washington | North Carolina | 0.00078 | Washington | North Dakota | 0.00147 | Washington | Ohio | 0.00389 |
| Washington | Oklahoma | 0.00204 | Washington | Oregon | 0.04473 | Washington | Pennsylvania | 0.00309 |
| Washington | Rhode Island | 0.00003 | Washington | South Carolina | 0.00034 | Washington | South Dakota | 0.00262 |
| Washington | Tennessee | 0.00174 | Washington | Texas | 0.01224 | Washington | Utah | 0.00839 |
| Washington | Vermont | 0.00181 | Washington | Virginia | 0.00073 | Washington | Washington | 0.69685 |
| Washington | West Virginia | 0.00017 | Washington | Wisconsin | 0.01348 | Washington | Wyoming | 0.00183 |
| West Virginia | Alabama | 0.00169 | West Virginia | Arizona | 0.00472 | West Virginia | Arkansas | 0.00208 |
| West Virginia | California | 0.01133 | West Virginia | Colorado | 0.00317 | West Virginia | Connecticut | 0.00377 |
| West Virginia | Delaware | 0.00187 | West Virginia | Florida | 0.03180 | West Virginia | Georgia | 0.02239 |
| West Virginia | Idaho | 0.00389 | West Virginia | Illinois | 0.01185 | West Virginia | Indiana | 0.05654 |
| West Virginia | Iowa | 0.00955 | West Virginia | Kansas | 0.00370 | West Virginia | Kentucky | 0.05347 |
| West Virginia | Louisiana | 0.00106 | West Virginia | Maine | 0.00274 | West Virginia | Maryland | 0.05669 |
| West Virginia | Massachusetts | 0.00360 | West Virginia | Michigan | 0.05753 | West Virginia | Minnesota | 0.01964 |
| West Virginia | Mississippi | 0.00168 | West Virginia | Missouri | 0.00892 | West Virginia | Montana | 0.00047 |
| West Virginia | Nebraska | 0.00051 | West Virginia | Nevada | 0.00020 | West Virginia | New Hampshire | 0.00116 |
| West Virginia | New Jersey | 0.00407 | West Virginia | New Mexico | 0.00344 | West Virginia | New York | 0.06104 |

| | | | | | | | | |
|---|---|---|---|---|---|---|---|---|
| West Virginia | North Carolina | 0.02636 | West Virginia | North Dakota | 0.00087 | West Virginia | Ohio | 0.14715 |
| West Virginia | Oklahoma | 0.00287 | West Virginia | Oregon | 0.00054 | West Virginia | Pennsylvania | 0.15425 |
| West Virginia | Rhode Island | 0.00023 | West Virginia | South Carolina | 0.00596 | West Virginia | South Dakota | 0.00215 |
| West Virginia | Tennessee | 0.02535 | West Virginia | Texas | 0.01658 | West Virginia | Utah | 0.00130 |
| West Virginia | Vermont | 0.01577 | West Virginia | Virginia | 0.07397 | West Virginia | Washington | 0.00262 |
| West Virginia | West Virginia | 0.02881 | West Virginia | Wisconsin | 0.05023 | West Virginia | Wyoming | 0.00044 |
| Wisconsin | Alabama | 0.00038 | Wisconsin | Arizona | 0.00223 | Wisconsin | Arkansas | 0.00100 |
| Wisconsin | California | 0.00498 | Wisconsin | Colorado | 0.00228 | Wisconsin | Connecticut | 0.00048 |
| Wisconsin | Delaware | 0.00008 | Wisconsin | Florida | 0.00502 | Wisconsin | Georgia | 0.00297 |
| Wisconsin | Idaho | 0.00203 | Wisconsin | Illinois | 0.02475 | Wisconsin | Indiana | 0.02609 |
| Wisconsin | Iowa | 0.04106 | Wisconsin | Kansas | 0.00330 | Wisconsin | Kentucky | 0.00783 |
| Wisconsin | Louisiana | 0.00031 | Wisconsin | Maine | 0.00043 | Wisconsin | Maryland | 0.00128 |
| Wisconsin | Massachusetts | 0.00053 | Wisconsin | Michigan | 0.04911 | Wisconsin | Minnesota | 0.09218 |
| Wisconsin | Mississippi | 0.00050 | Wisconsin | Missouri | 0.00788 | Wisconsin | Montana | 0.00044 |
| Wisconsin | Nebraska | 0.00067 | Wisconsin | Nevada | 0.00012 | Wisconsin | New Hampshire | 0.00019 |
| Wisconsin | New Jersey | 0.00026 | Wisconsin | New Mexico | 0.00185 | Wisconsin | New York | 0.00756 |
| Wisconsin | North Carolina | 0.00127 | Wisconsin | North Dakota | 0.00111 | Wisconsin | Ohio | 0.01430 |
| Wisconsin | Oklahoma | 0.00184 | Wisconsin | Oregon | 0.00032 | Wisconsin | Pennsylvania | 0.00666 |
| Wisconsin | Rhode Island | 0.00003 | Wisconsin | South Carolina | 0.00055 | Wisconsin | South Dakota | 0.00379 |
| Wisconsin | Tennessee | 0.00335 | Wisconsin | Texas | 0.00777 | Wisconsin | Utah | 0.00083 |
| Wisconsin | Vermont | 0.00266 | Wisconsin | Virginia | 0.00143 | Wisconsin | Washington | 0.00141 |
| Wisconsin | West Virginia | 0.00040 | Wisconsin | Wisconsin | 0.66412 | Wisconsin | Wyoming | 0.00035 |
| Wyoming | Alabama | 0.00069 | Wyoming | Arizona | 0.03712 | Wyoming | Arkansas | 0.00268 |
| Wyoming | California | 0.10129 | Wyoming | Colorado | 0.12777 | Wyoming | Connecticut | 0.00088 |
| Wyoming | Delaware | 0.00009 | Wyoming | Florida | 0.01482 | Wyoming | Georgia | 0.00498 |
| Wyoming | Idaho | 0.13957 | Wyoming | Illinois | 0.00861 | Wyoming | Indiana | 0.01548 |
| Wyoming | Iowa | 0.02006 | Wyoming | Kansas | 0.02116 | Wyoming | Kentucky | 0.00873 |
| Wyoming | Louisiana | 0.00088 | Wyoming | Maine | 0.00088 | Wyoming | Maryland | 0.00177 |
| Wyoming | Massachusetts | 0.00082 | Wyoming | Michigan | 0.02173 | Wyoming | Minnesota | 0.05701 |
| Wyoming | Mississippi | 0.00101 | Wyoming | Missouri | 0.01242 | Wyoming | Montana | 0.02132 |
| Wyoming | Nebraska | 0.00555 | Wyoming | Nevada | 0.00382 | Wyoming | New Hampshire | 0.00057 |
| Wyoming | New Jersey | 0.00038 | Wyoming | New Mexico | 0.02280 | Wyoming | New York | 0.01025 |
| Wyoming | North Carolina | 0.00240 | Wyoming | North Dakota | 0.00892 | Wyoming | Ohio | 0.01299 |
| Wyoming | Oklahoma | 0.00889 | Wyoming | Oregon | 0.00662 | Wyoming | Pennsylvania | 0.00959 |
| Wyoming | Rhode Island | 0.00006 | Wyoming | South Carolina | 0.00107 | Wyoming | South Dakota | 0.02069 |
| Wyoming | Tennessee | 0.00605 | Wyoming | Texas | 0.04803 | Wyoming | Utah | 0.06026 |
| Wyoming | Vermont | 0.00454 | Wyoming | Virginia | 0.00170 | Wyoming | Washington | 0.03046 |
| Wyoming | West Virginia | 0.00038 | Wyoming | Wisconsin | 0.05143 | Wyoming | Wyoming | 0.06077 |

After stochastically allocating a destination state in accordance with the above movement matrix, we then also stochastically assign which herd in the destination state the exported cattle will be sent to. If a herd was picked at random with equal weighting, this would cause all herd populations to eventually converge to the size herd size, since the number of cattle exported is a proportion of the origin herd's total population. Therefore, this probability is weighted by the respective candidate destination herd's total population. For herd $i$ in the destination state, the probability of the cattle being sent to that respective herd is $\frac{N_i}{\sum_{k=1}^{N_{\text{herds}}} N_k}$.

At this stage, we know i) if the herd in question is exporting cattle, ii) how many cattle are being exported, and iii) which herd and state it is exporting to. However, before moving the cattle from the epidemiological compartments of the origin herd to the destination herd, we have to decide if the exported cattle pass the interstate testing mandates. As detailed in the main manuscript, as of April 29th 2024, cattle exported interstate have up to 30 cows in the cohort tested for H5N1 influenza [3]. Should the herd test positive, the export cannot proceed, and the origin herd must be quarantined for 30 days before being tested again.

If the destination state is the same as the origin state, i.e. the exported cattle are not crossing state boundaries, then the export proceeds - movements within-state are not tested. If the model time step is before April 29th 2024, then the export proceeds. If the destination herd is in a different state, and if the model time step is at a point after April 29th 2024, then we simulate the testing of the exported cattle. If there are less then 30 cattle in the exported cohort, then all the cattle will be tested. Therefore, if there are any infected cattle in the exported cohort, i.e. $I_{i,j}^{\text{exported}} > 0$, then the export does not go ahead, and the cattle remain in their origin herd. Note that cattle in the exposed category, $E_{i,j}^{\text{exported}}$, will still be exported, allowing for a scenario where newly infected cattle, who have not yet incubated the virus long enough to test positive, will be exported.

If there are more than 30 cattle being exported, we simulate the process of testing 30 cows for infection via a hypergeometric distribution:

$$X \sim \text{Hypergeometric}\left(I_{i,j}^{\text{exported}}, S_{i,j}^{\text{exported}} + E_{i,j}^{\text{exported}} + R_{i,j}^{\text{exported}}, 30\right). \quad (17)$$

i.e. we make 30 draws, without replacement, from a population made up of $I_{i,j}^{\text{exported}}$ "successes", and $S_{i,j}^{\text{exported}} + E_{i,j}^{\text{exported}} + R_{i,j}^{\text{exported}}$ "failures". Thus, if $X > 0$, then an infected cow has been detected in the cohort, and the export does not proceed. If, instead, $X = 0$, then the export of cattle proceeds.

#### 2.5 Epidemiological Parameter Fitting

We fit the five epidemiological parameters detailed in Table S2 via Bayesian evidence synthesis. We build a likelihood function comparing the fit of the step function of "Time of first outbreak detection" for each state to the model simulation of said function. The data extracted from the USDA outbreak portal provides $Y_{j,t}$ - the number of new reported outbreaks in week $t$ in state $j$. We convert this to the step function, $Y_{j,t}^{\text{first outbreak}}$ like so:

$$Y_{j,\tau}^{\text{first outbreak}} = \begin{cases} 0 & \text{if } Y_{j,t} = 0 \quad \forall\, t < \tau \\ 1 & \text{if } Y_{j,t} \neq 0 \quad \forall\, t < \tau. \end{cases} \tag{18}$$

We use the following likelihood function within our Bayesian evidence synthesis approach;

$$Y_{j,\tau}^{\text{first outbreak}} \sim \text{Bernoulli}(\hat{Y}_{j,\tau}^{\text{first outbreak}}). \tag{19}$$

Where $\hat{Y}_{j,\tau}^{\text{first outbreak}}$, is the model simulation of the "time of first outbreak" function. Therefore,

$$\text{Log-likelihood} = \sum_{j=1}^{48}\sum_{t=1}^{T} Y_{j,t}^{\text{first outbreak}} \log\left(\varepsilon + \hat{Y}_{j,t}^{\text{first outbreak}}\right) + \left(1 - Y_{j,t}^{\text{first outbreak}}\right)\log\left(1 - \varepsilon + \hat{Y}_{j,t}^{\text{first outbreak}}\right), \tag{20}$$

where $\varepsilon = 1\text{e}^{-6}$ is a very small value to avoid the evaluation of log(0).

We also produce all model results using a different likelihood choice - fitting to the number of new weekly reported outbreaks by state, instead of "time of first outbreak detection". This format and results are presented in Section 3.2.1 below.

The fitting is performed via particle Markov Chain Monte Carlo simulation (pMCMC) [4, 5] via the `dust2` and `monty` packages. 16 chains are run for 40,000 iterations. To avoid stuck chains and improve parameter space exploration, particle weighting is reset every 200 iterations. Convergence is assessed via the potential scale reduction factor (psrf), with all parameters displaying a psrf < 1.1. Prior distribution, posterior distribution, and psrf for each parameter is given in Table S6. Note that we fit $\frac{\beta}{\gamma}$ instead of just $\beta$ to improve chain mixing due to observed correlation between $\beta$ and $\gamma$.

| Parameter | Prior Distribution | Posterior median (95% CrI) | psrf |
|---|---|---|---|
| $\frac{\beta}{\gamma}$ | Uniform(0.05, 3) | 1.864 (0.929-2.932) | 1.06 |
| $\alpha$ | Uniform(0, 0.1) | 0.063 (0.009-0.098) | 1.01 |
| $\sigma$ | Uniform(0.05, 2) | 1.050 (0.199-1.956) | 1.06 |
| $\gamma$ | Uniform(0.05, 2) | 1.084 (0.384-1.942) | 1.08 |
| $A^{\text{asc}}$ | Beta(1, 1) | 0.648 (0.091-0.986) | 1.01 |

**Table S6:** Prior distributions, posterior distributions, and potential scale reduction factors (psrf) for each model parameter fit.

Figures S2 to S7 plot the chain trajectories for the first 4 of the 16 chains for each parameter, and the associated log-likelihoods. Figures S8 to S12 show the density plots of the prior and posterior distributions for each parameter.

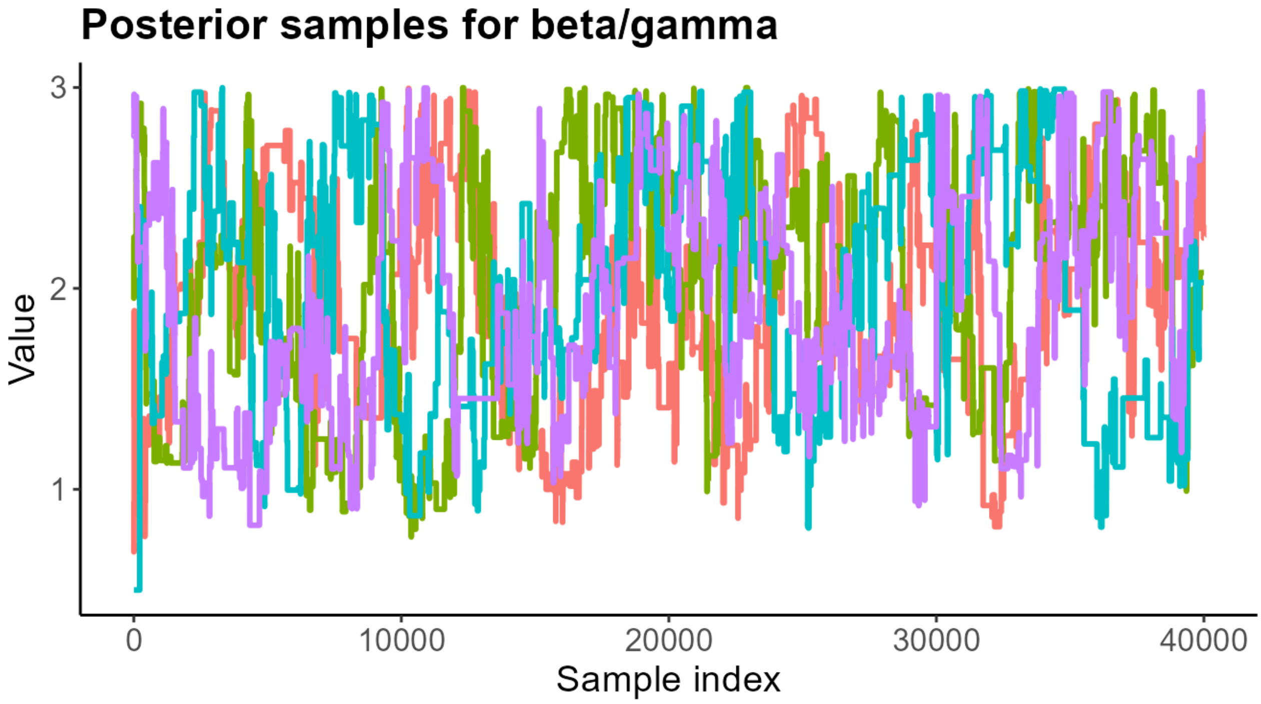

**Figure S2:** The posterior chain trajectories for all 40000 steps of the pMCMC for the first four of the 16 chains for $\frac{\beta}{\gamma}$.

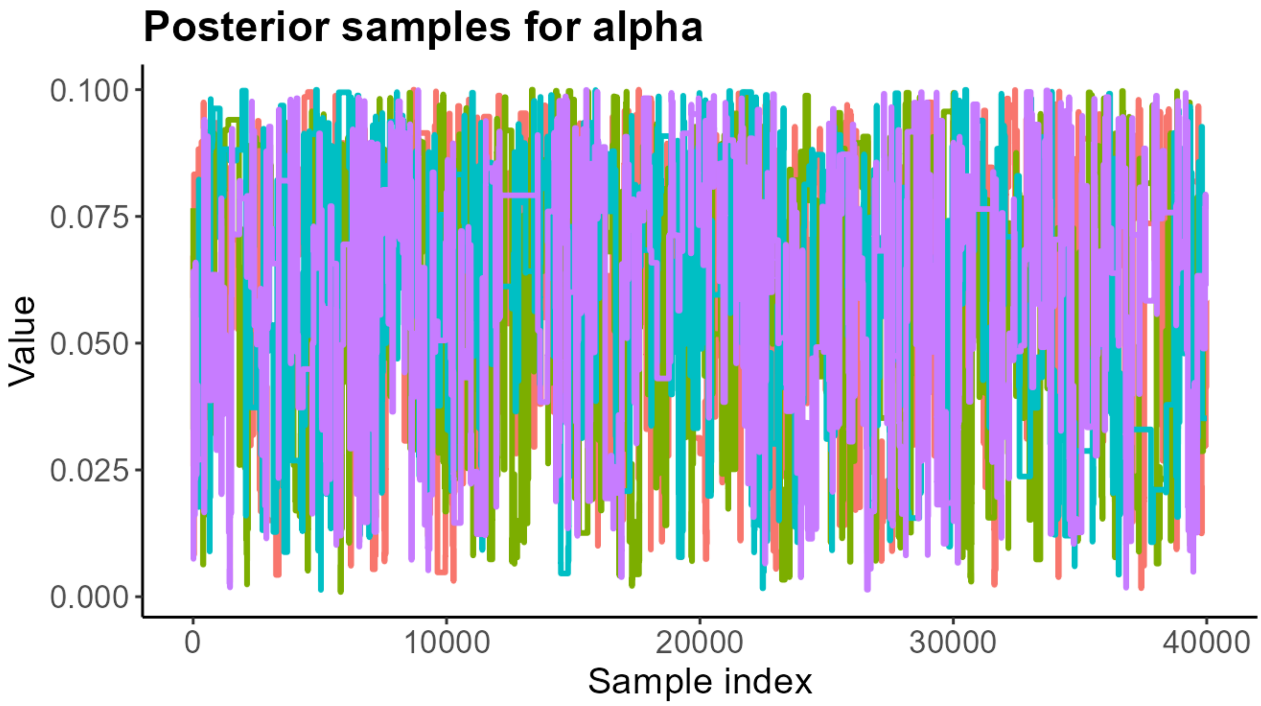

**Figure S3:** The posterior chain trajectories for all 40000 steps of the pMCMC for the first four of the 16 chains for $\alpha$.

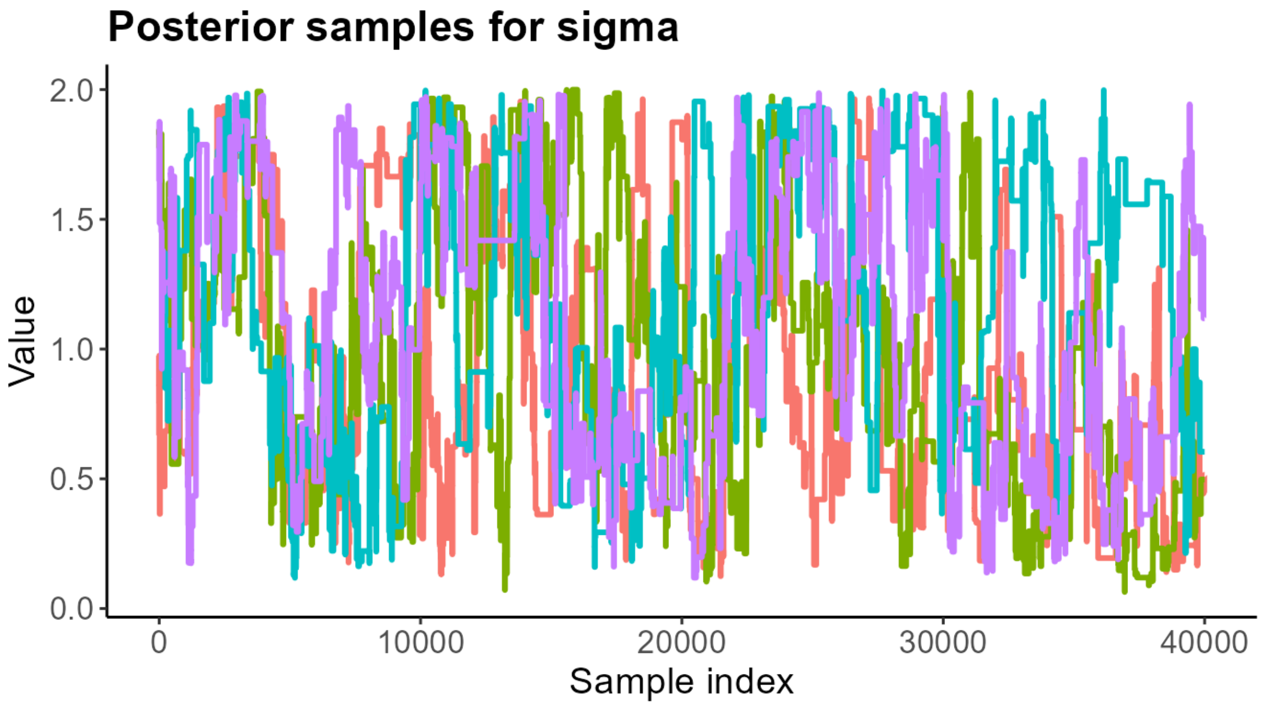

**Figure S4:** The posterior chain trajectories for all 40000 steps of the pMCMC for the first four of the 16 chains for $\sigma$.

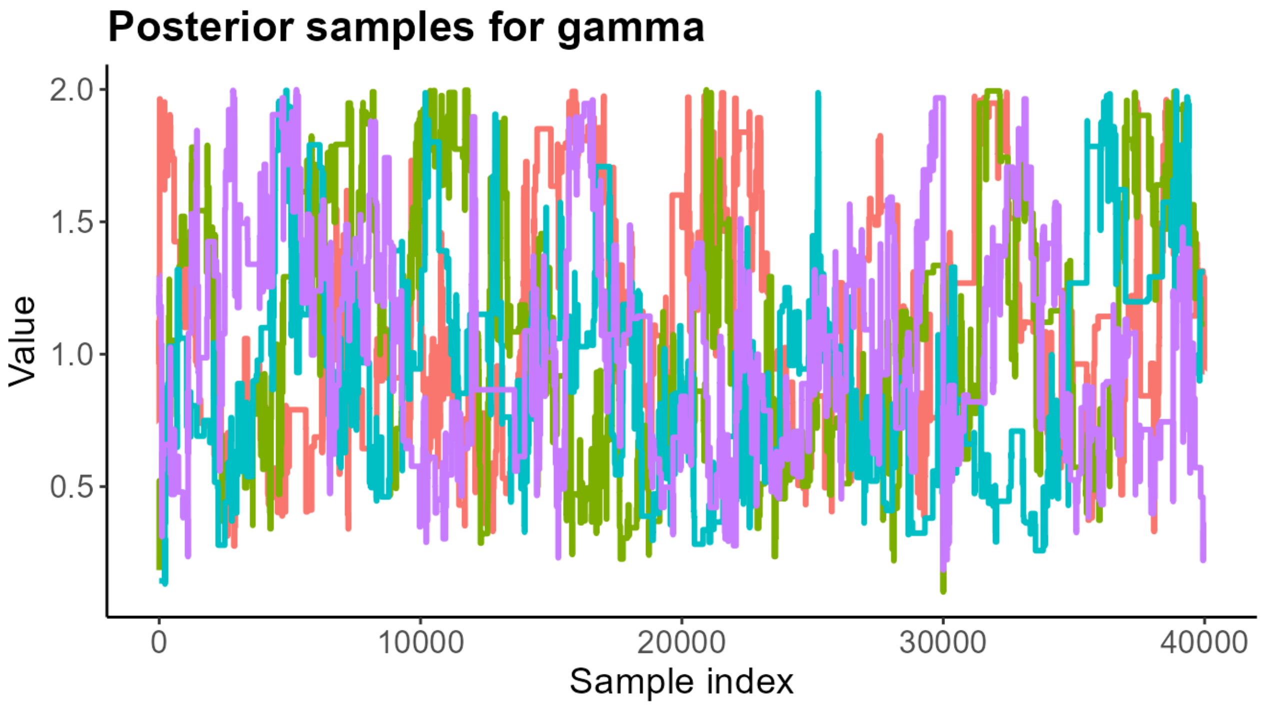

**Figure S5:** The posterior chain trajectories for all 40000 steps of the pMCMC for the first four of the 16 chains for $\gamma$.

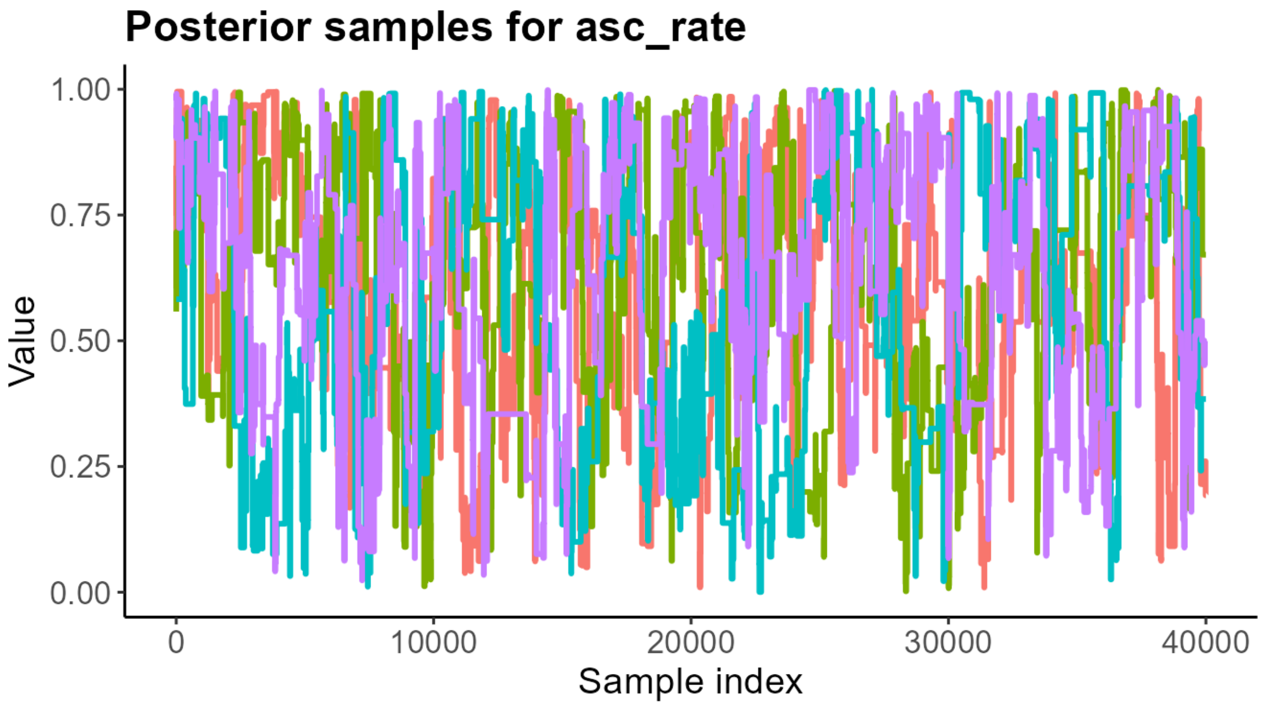

**Figure S6:** The posterior chain trajectories for all 40000 steps of the pMCMC for the first four of the 16 chains for $A^{\text{asc}}$.

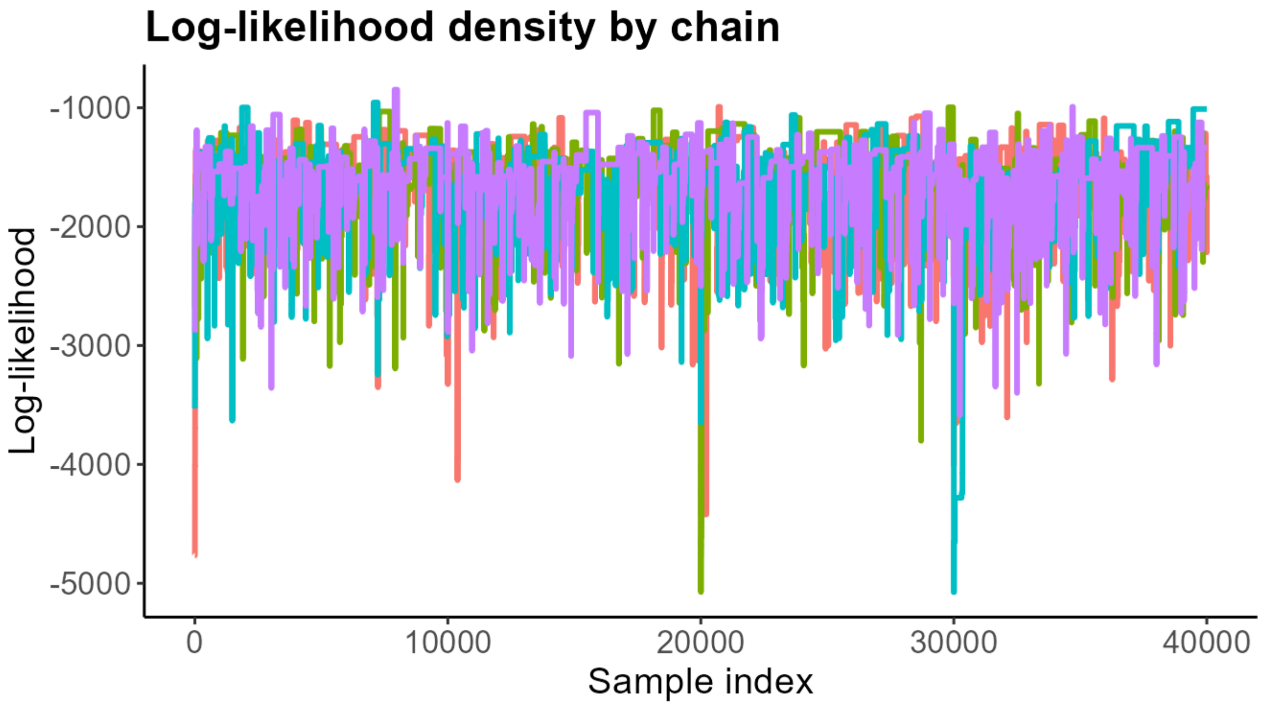

**Figure S7:** The log-likelihoods for all 40000 steps of the pMCMC for the first four of the 16 chains.

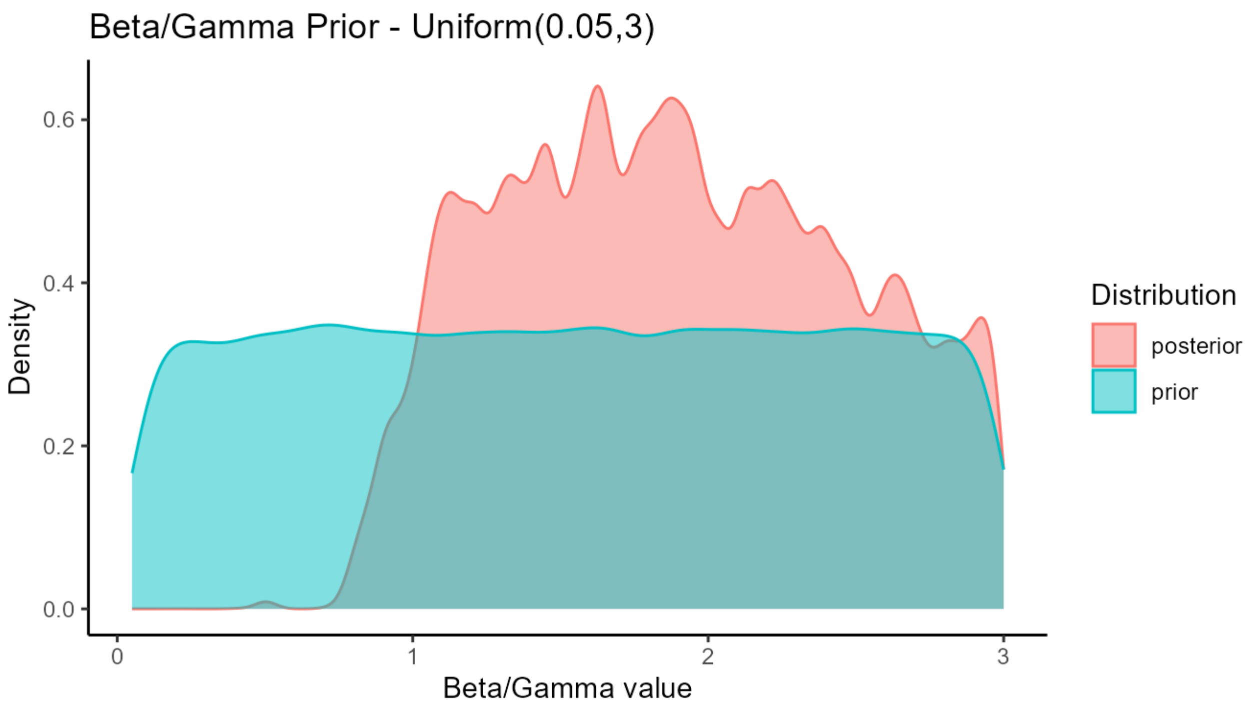

**Figure S8:** The density plots of the prior and posterior distributions for $\frac{\beta}{\gamma}$.

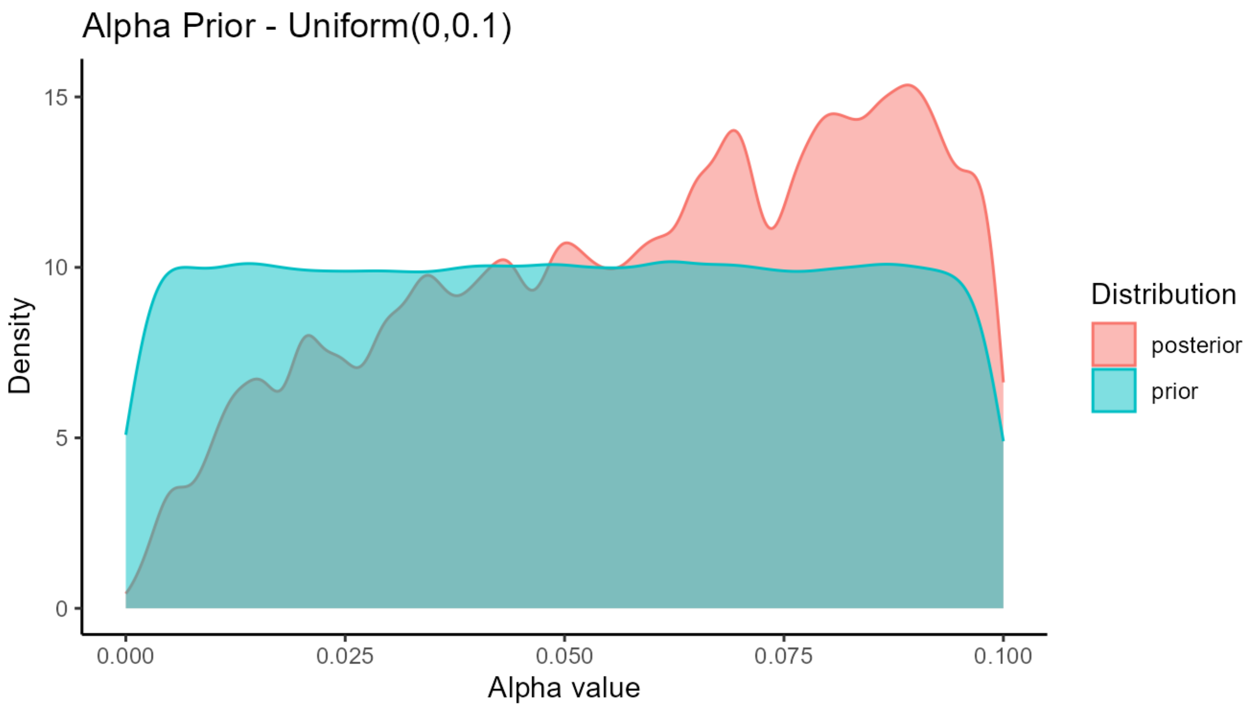

**Figure S9:** The density plots of the prior and posterior distributions for $\alpha$.

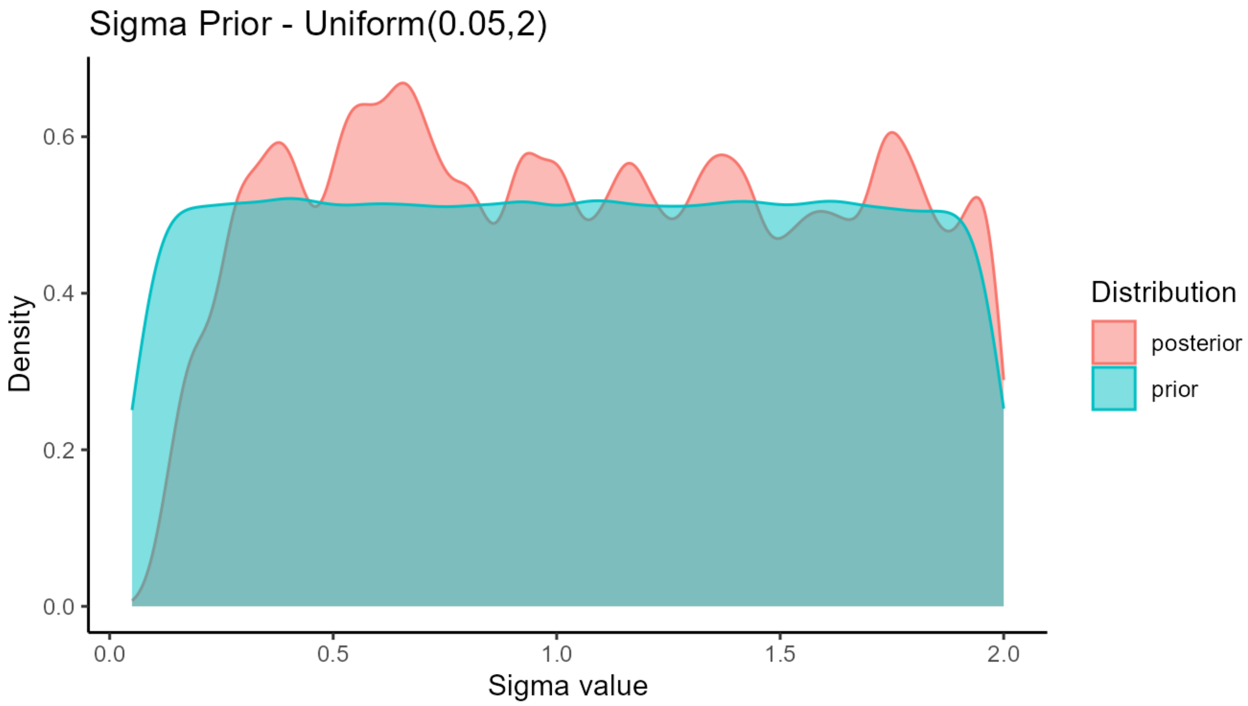

**Figure S10:** The density plots of the prior and posterior distributions for $\sigma$.

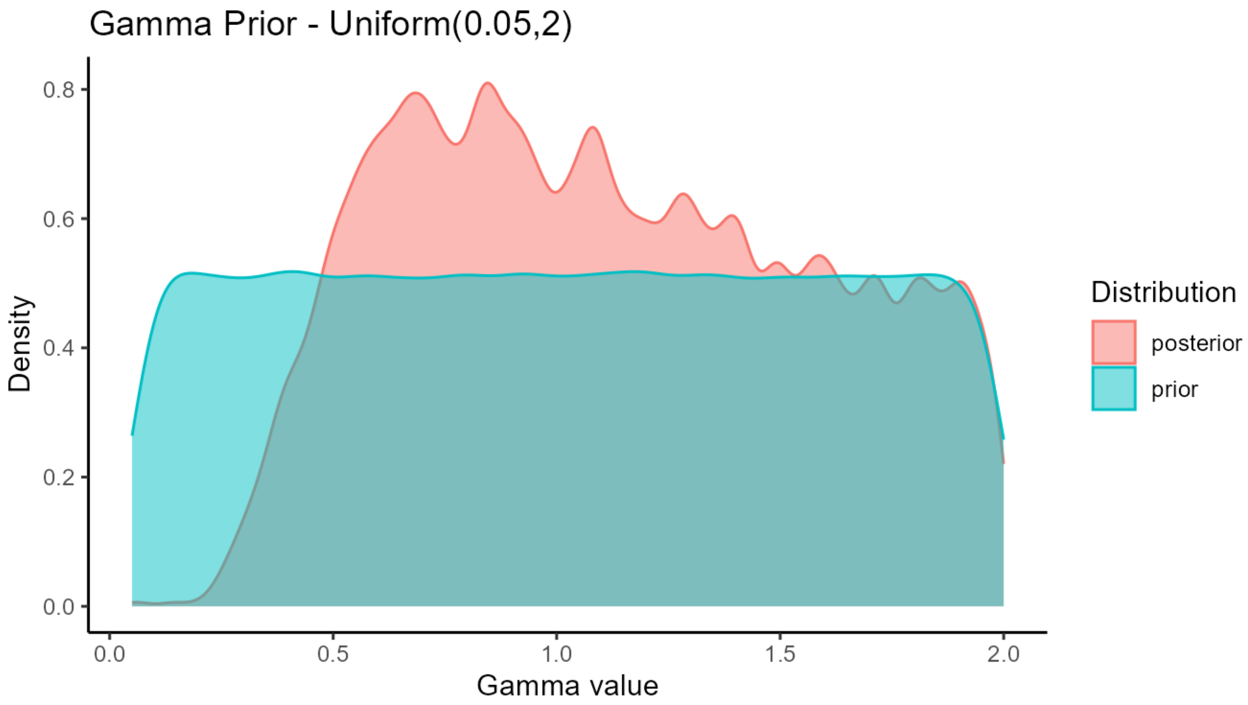

**Figure S11:** The density plots of the prior and posterior distributions for $\gamma$.

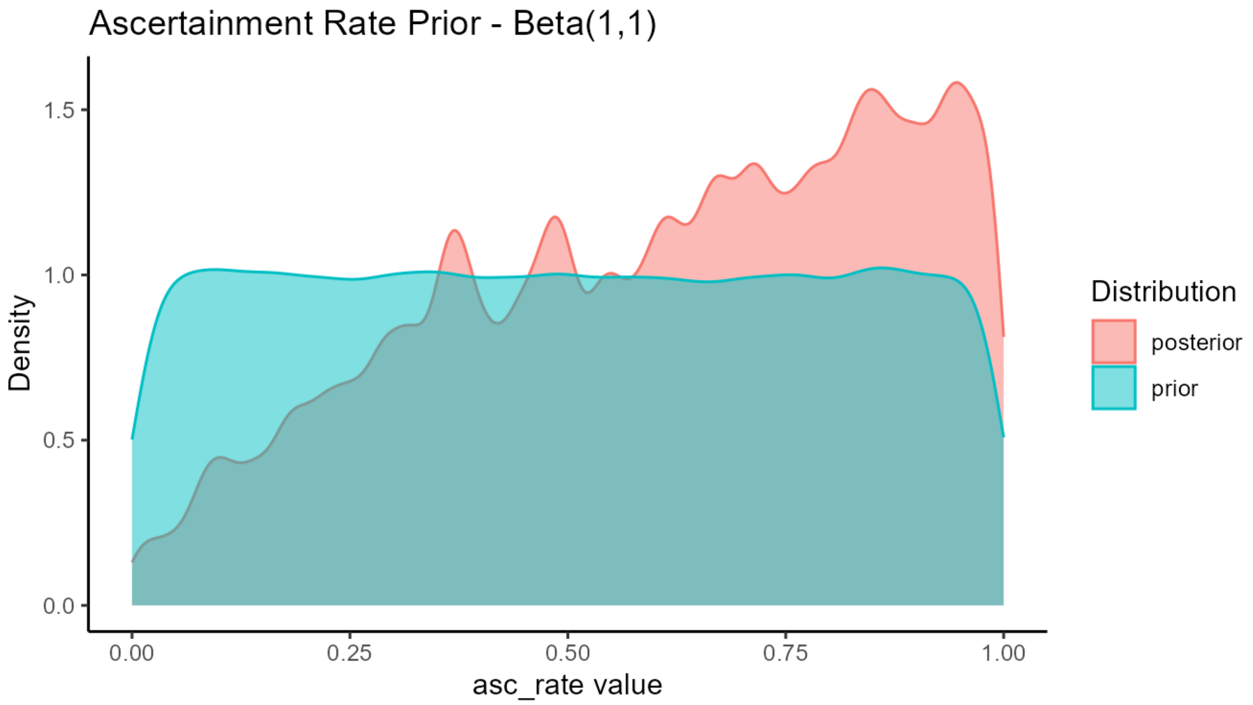

**Figure S12:** The density plots of the prior and posterior distributions for $A^{\text{asc}}$.

### 3 Supplementary Results

#### 3.1 Additional Results

Figure 2A in the main manuscript presents the model simulation results for the time of first outbreak reported for each state. Figure 2B presents the model simulation results for the number of new weekly reported outbreaks by state. Figure S13 below plots an additional model output to accompany these results - the "true" proportion of infected herds. i.e. the proportion of herds in each state over time that contain any infected cattle. Figure 2B incorporates the impact of outbreak ascertainment, while Figure S13 does not.

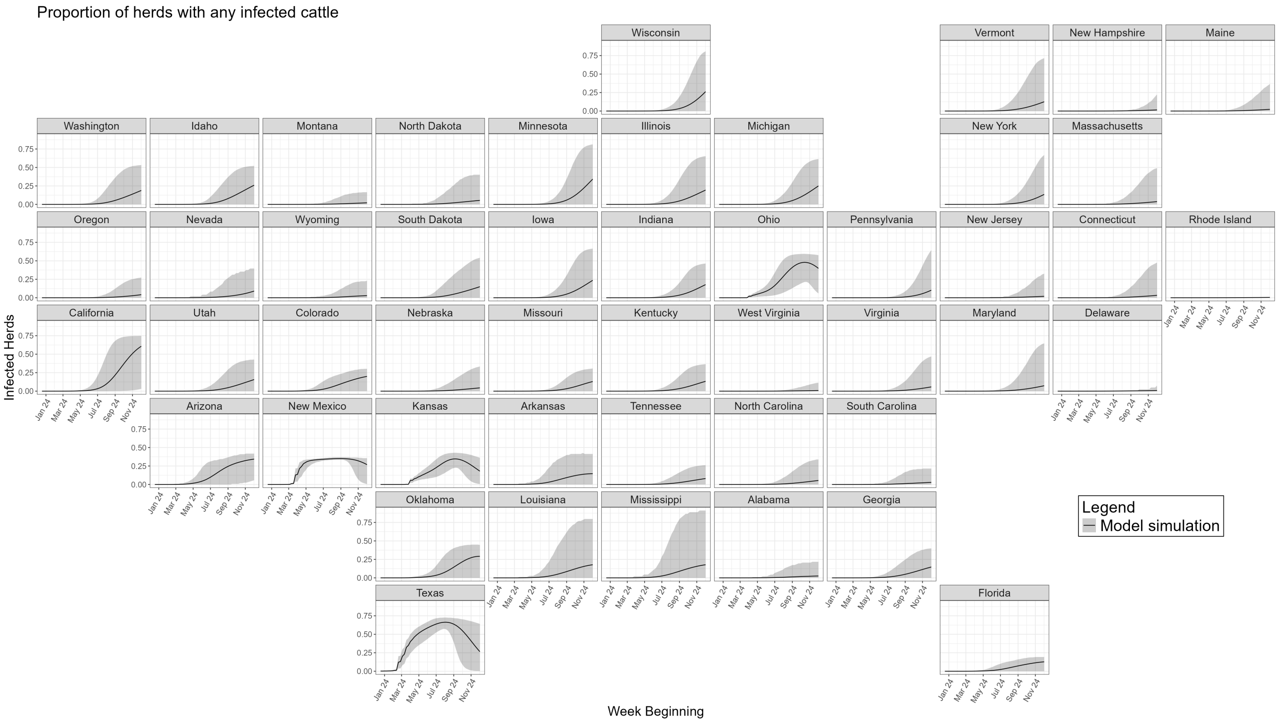

**Figure S13:** An additional model output accompanying Figure 2 of the main manuscript. The model simulation results of the proportion of herds in each state over time containing any infected cattle. 20,000 model simulations are run, the black line represents the mean result, and the shaded region the 95% CrI.

Figure 4 in the main manuscript displays the mean probability that an exported herd would test positive, for all US states during 3 different weeks over the course of the epidemic. Figure S14 below displays forest plots of those same means but with the associated 95% CrIs.

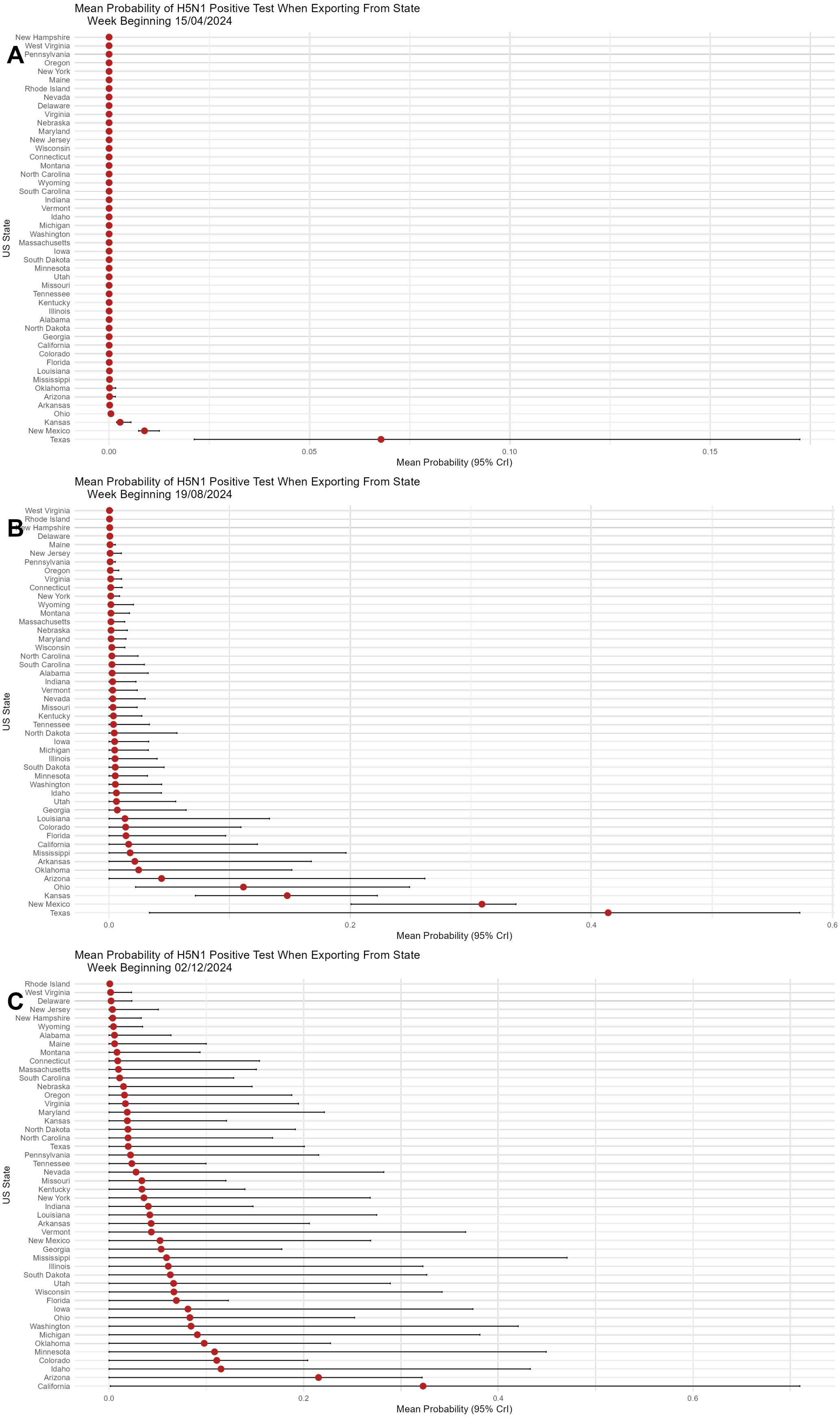

**Figure S14:** When moving cattle inter-state, up to 30 cattle will be tested for H5N1. Above panels show the state average probability of such an export testing positive over time: **(A)** week beginning April 15th 2024, **(B)** week beginning August 19th 2024, and **(C)** week beginning December 2nd 2024. Red dots indicate mean probability, and black bars the 95% CrIs.

#### 3.2 Sensitivity Analyses

To explore the implications of modelling decisions, we present alternate versions of all main manuscript figures, but with parameters fit and simulations run under different modeling assumptions. We see that the main findings and presented results are unchanged under these assumptions.

##### 3.2.1 Alternate likelihood

As detailed in Section 2.5 above, our main results are fit using a likelihood function set to the "time of first outbreak detection" instead of weekly outbreak incidence. This is because the number of outbreaks are rarely true independent data samples. When some sites reported outbreaks, some neighbouring farms would be checked or seek testing, meaning the number of reported outbreaks is likely to be extremely auto-correlated. Nonetheless, we also present our results when a likelihood function fitting to the weekly new number of reported outbreaks is used:

$$Y_{j,\tau}^{\text{first outbreak}} \sim \text{Negative Binomial}(\text{mean} = \hat{Y}_{j,\tau}^{\text{first outbreak}}, \text{dispersion} = 1). \tag{21}$$

Figures S15 to S17 below present new versions of Figures 2, 4, and 5 from the main manuscript reproduced under this modeling choice.

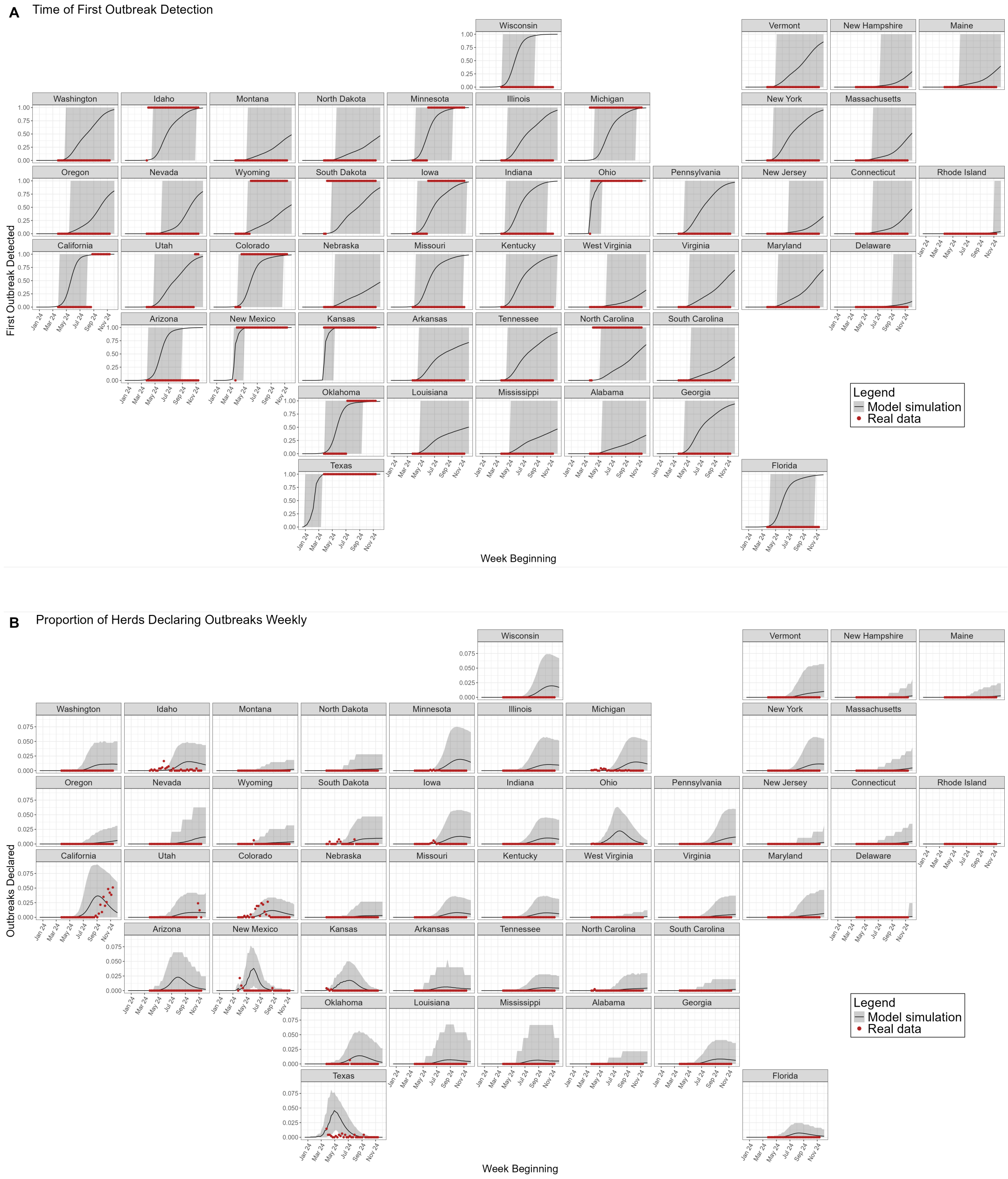

**Figure S15:** This is a recreation of Figure 2 from the main manuscript when using a likelihood function fitting to new weekly reported outbreaks. **(A)** shows the time at which the first outbreak is detected in a state. **(B)** shows the proportion of herds in each state which report new outbreaks per state each week, accounting for under-reporting. Red points depict real world data. The black line depicts the model mean, the shaded grey region depicts the 95% credible interval (95% CrI).

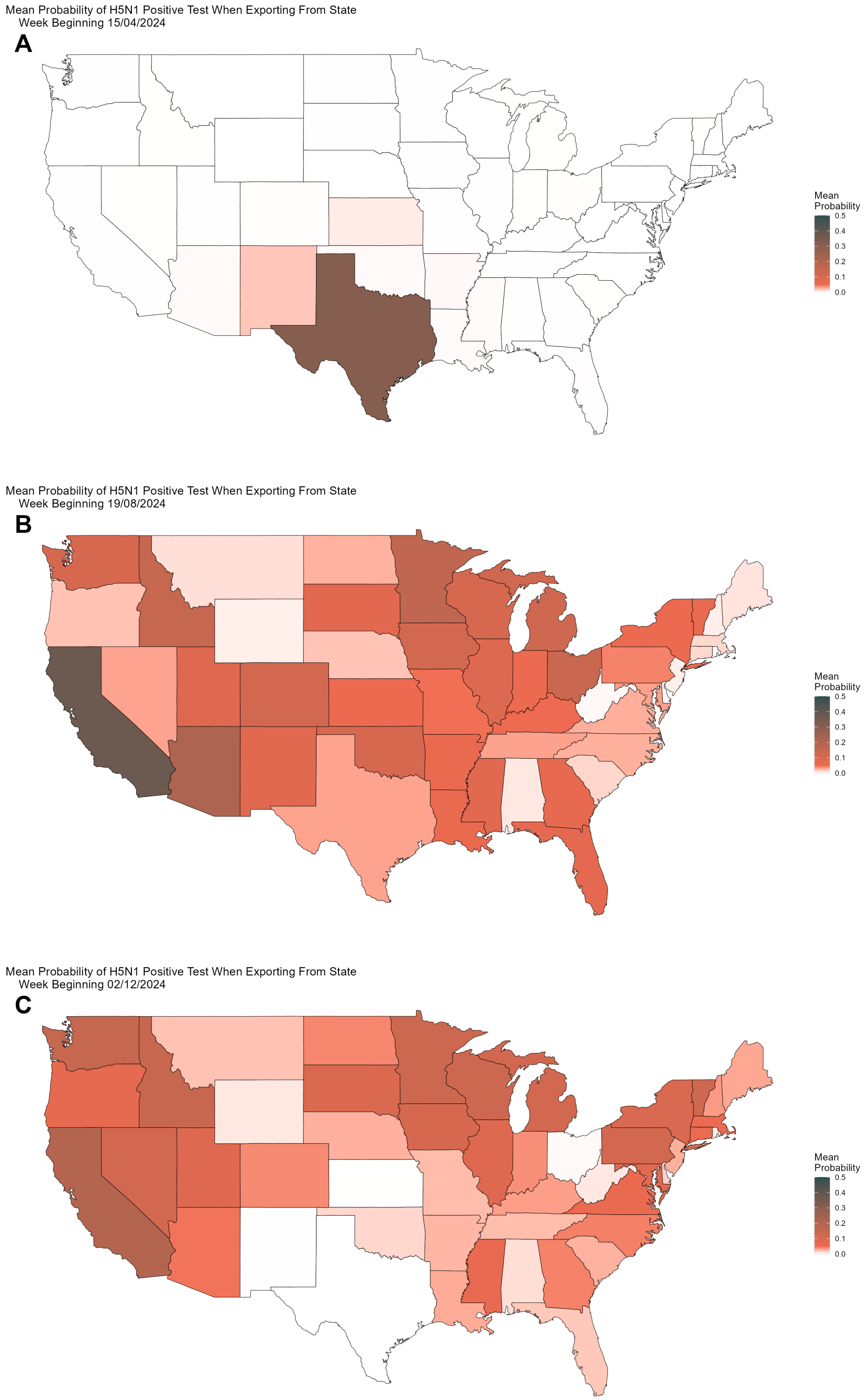

**Figure S16:** This is a recreation of Figure 4 from the main manuscript when using a likelihood function fitting to new weekly reported outbreaks. When moving cattle inter-state, up to 30 cattle will be tested for H5N1. Above panels show the state average probability of such an export testing positive over time: **(A)** week beginning April 15th 2024, **(B)** week beginning August 19th 2024, and **(C)** week beginning December 2nd 2024.

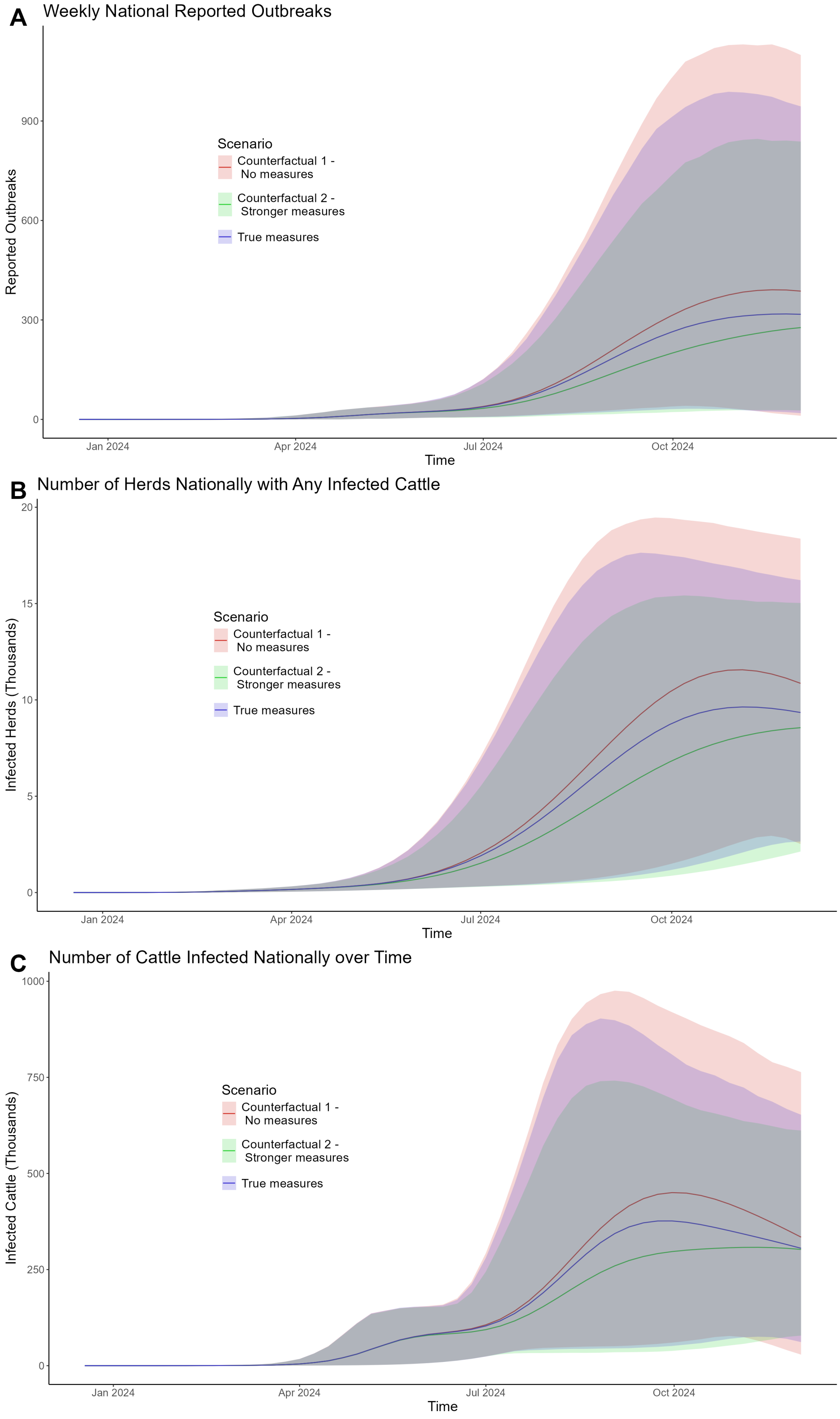

**Figure S17:** This is a recreation of Figure 5 from the main manuscript when using a likelihood function fitting to new weekly reported outbreaks. **(A)** The number of new reported outbreaks weekly. **(B)** The number of herds nationally with any infected cattle. **(C)** The total number of infected cows nationally over time. Solid lines show simulation mean. Shaded regions show 95% CrI. Blue depicts baseline model assumptions. Red depicts the scenario with no border testing. Green depicts border testing of up to 100 cows, implemented 28 days earlier, on April 1st 2024.

#### 3.2.2 Real ICVI data

All the model assumptions on probabilities of export and movement matrices are based upon USAMM model outputs, as detailed in section 2.4 above. While this is the most comprehensive source of information regarding the inherent uncertainty regarding cattle movements, alternate data streams are available. We were provided the real world ICVI data gathered by Cabezas et al. (2021) [6] detailing a series of exact cattle shipments for the year 2016. Using the same methods as detailed above using the USAMM results, we reproduce our probability of export (Table S3) and movement matrix (Table S5) using this data instead.

Figures S18 to S20 below present new versions of Figures 2, 4, and 5 from the main manuscript reproduced with model fits and simulations using these movement probabilities instead.

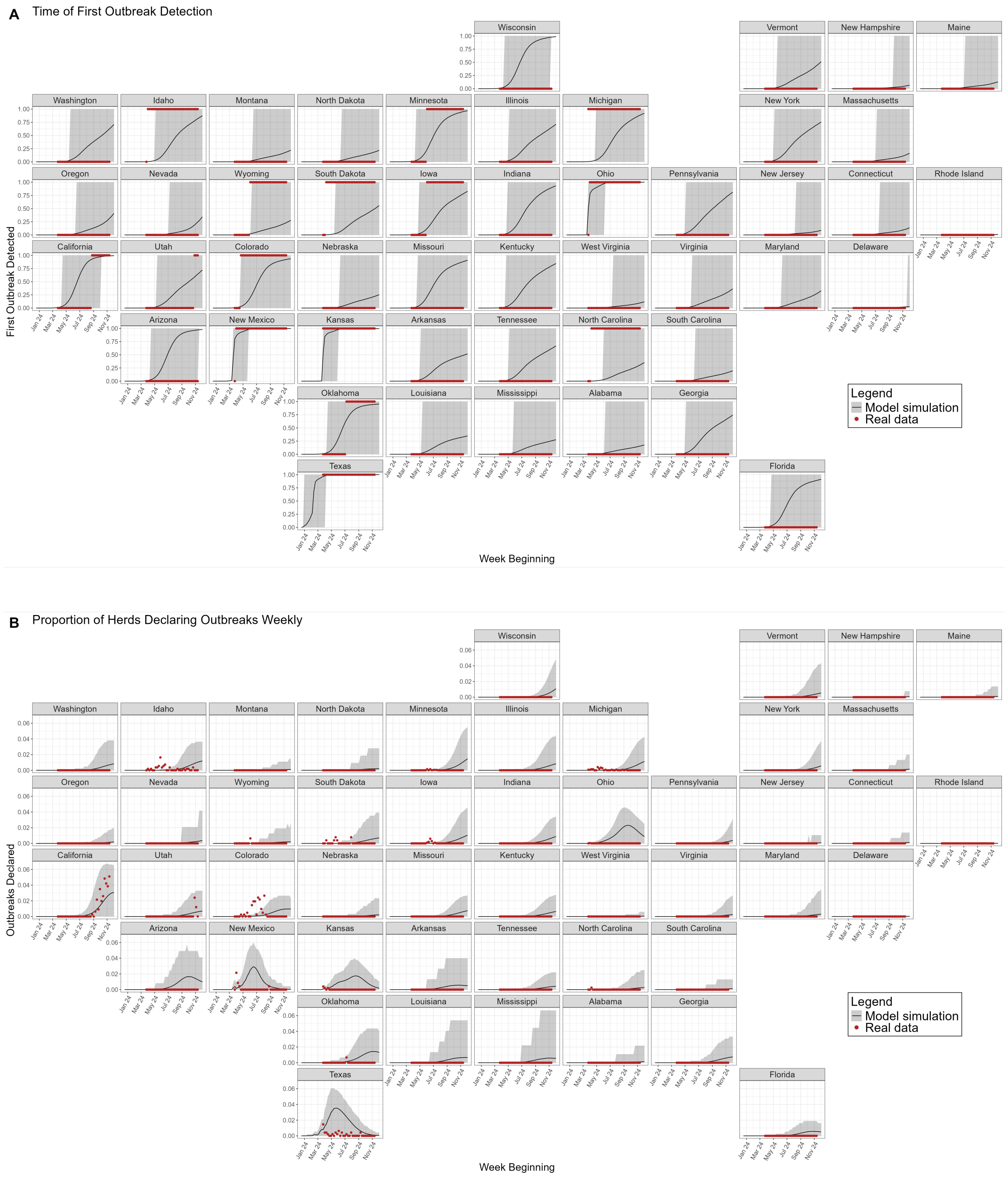

**Figure S18:** This is a recreation of Figure 2 from the main manuscript when using movement probabilities directly calculated from 2016 ICVI data. **(A)** shows the time at which the first outbreak is detected in a state. **(B)** shows the proportion of herds in each state which report new outbreaks per state each week, accounting for under-reporting. Red points depict real world data. The black line depicts the model mean, the shaded grey region depicts the 95% credible interval (95% CrI).

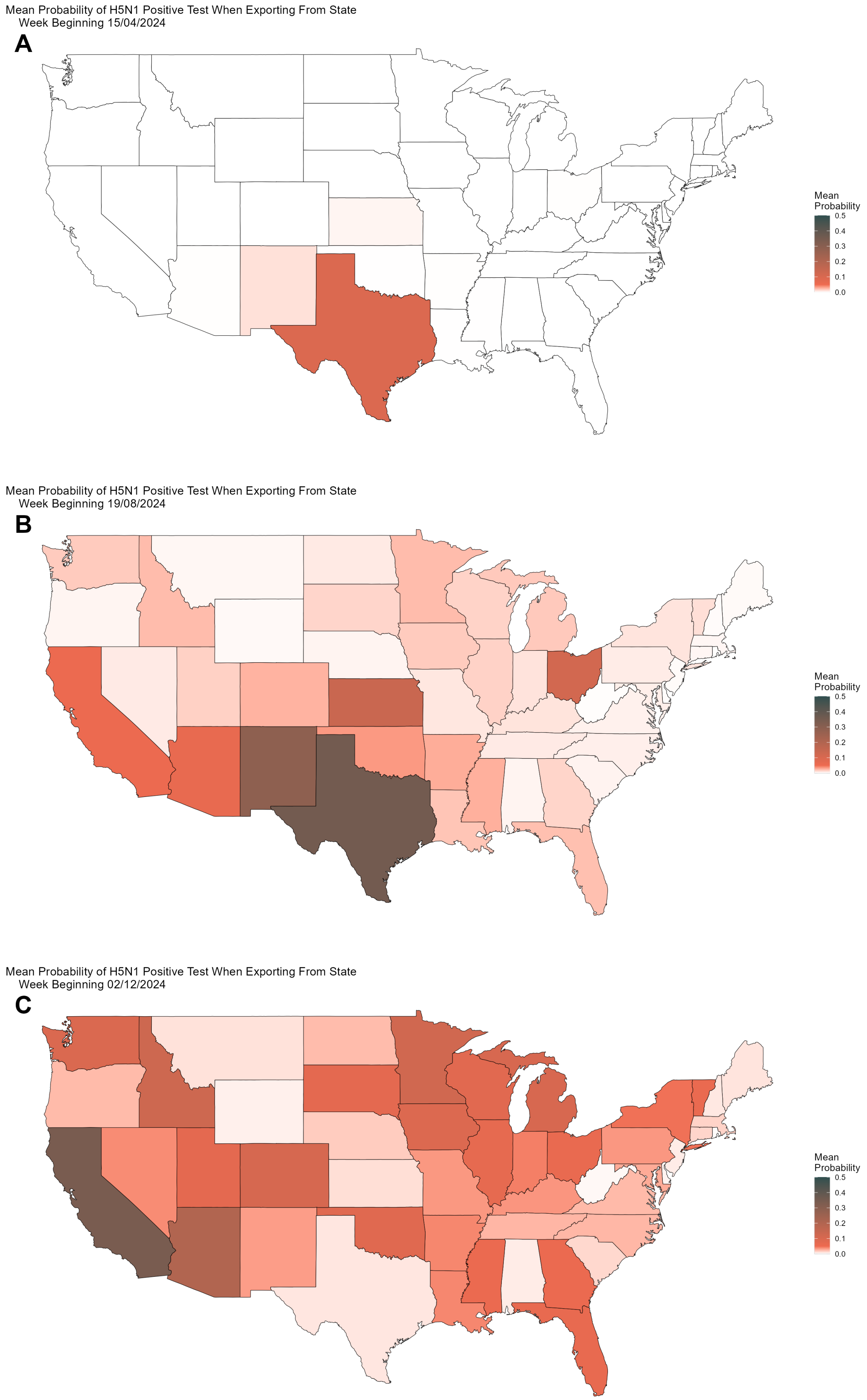

**Figure S19:** This is a recreation of Figure 4 from the main manuscript when using movement probabilities directly calculated from 2016 ICVI data. When moving cattle inter-state, up to 30 cattle will be tested for H5N1. Above panels show the state average probability of such an export testing positive over time: **(A)** week beginning April 15th 2024, **(B)** week beginning August 19th 2024, and **(C)** week beginning December 2nd 2024.

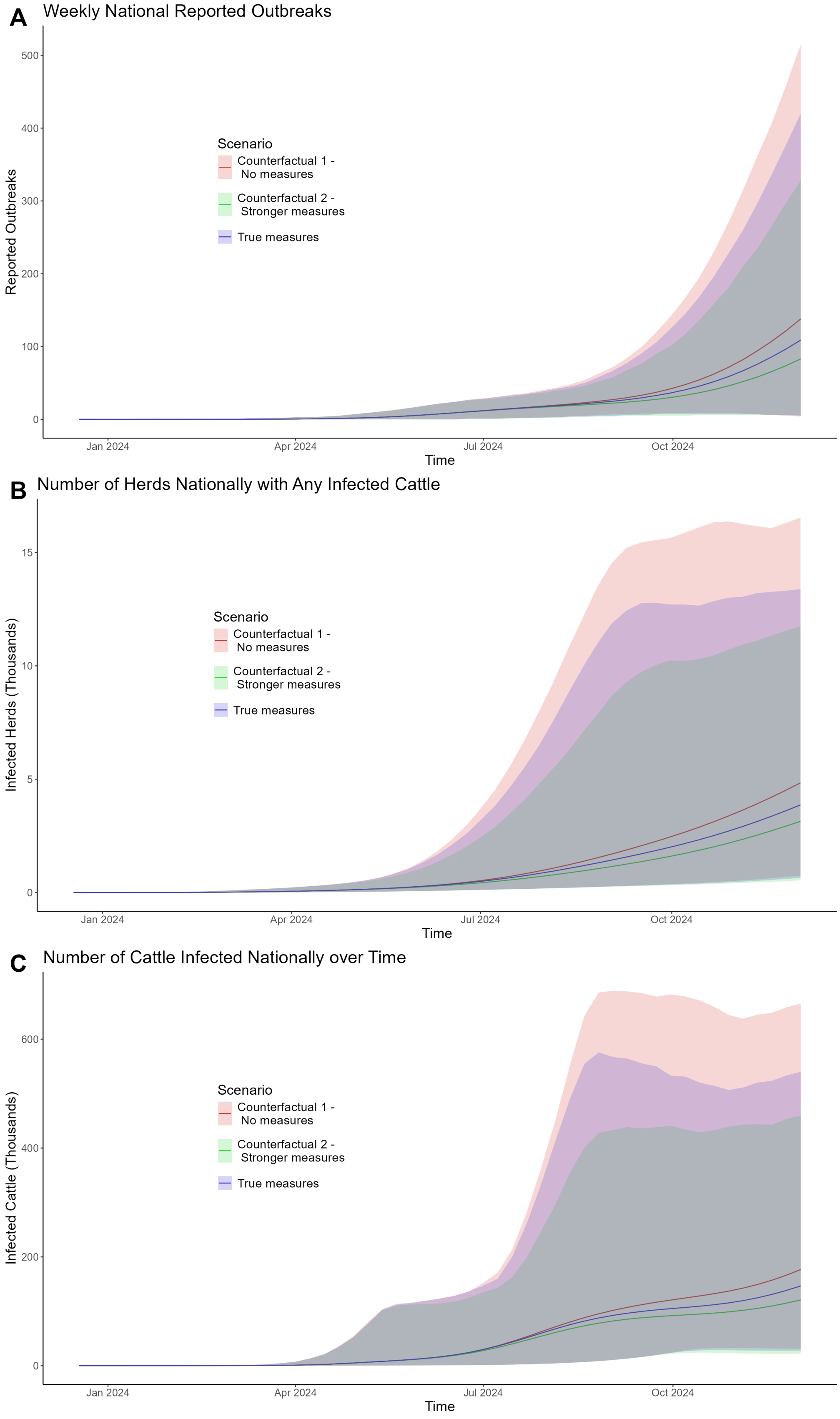

**Figure S20:** This is a recreation of Figure 5 from the main manuscript when using movement probabilities directly calculated from 2016 ICVI data. **(A)** The number of new reported outbreaks weekly. **(B)** The number of herds nationally with any infected cattle. **(C)** The total number of infected cows nationally over time. Solid lines show simulation mean. Shaded regions show 95% CrI. Blue depicts baseline model assumptions. Red depicts the scenario with no border testing. Green depicts border testing of up to 100 cows, implemented 28 days earlier, on April 1st 2024.

#### 3.2.3 Alternate Ascertainment

The base model assumes that the probability of a herd reporting an outbreak is dependent on the absolute number of infected cattle in the herd, and the proportion of the herd is infected. These assumptions are outlaid in Equation (12) above.

In this sensitivity analysis, we re-fit the model and produce all model results using a more relaxed model assumption, where ascertainment is scaled just by the proportion of the herd that is infected. Specifically, we replace Equation (12) with:

$$\phi_i = \left(\frac{I_i}{N_i}\right) A^{\text{asc}} dt. \qquad (22)$$

Figures S21 to S23 below present new versions of Figures 2, 4, and 5 from the main manuscript reproduced with model fits and simulations using these movement probabilities instead.

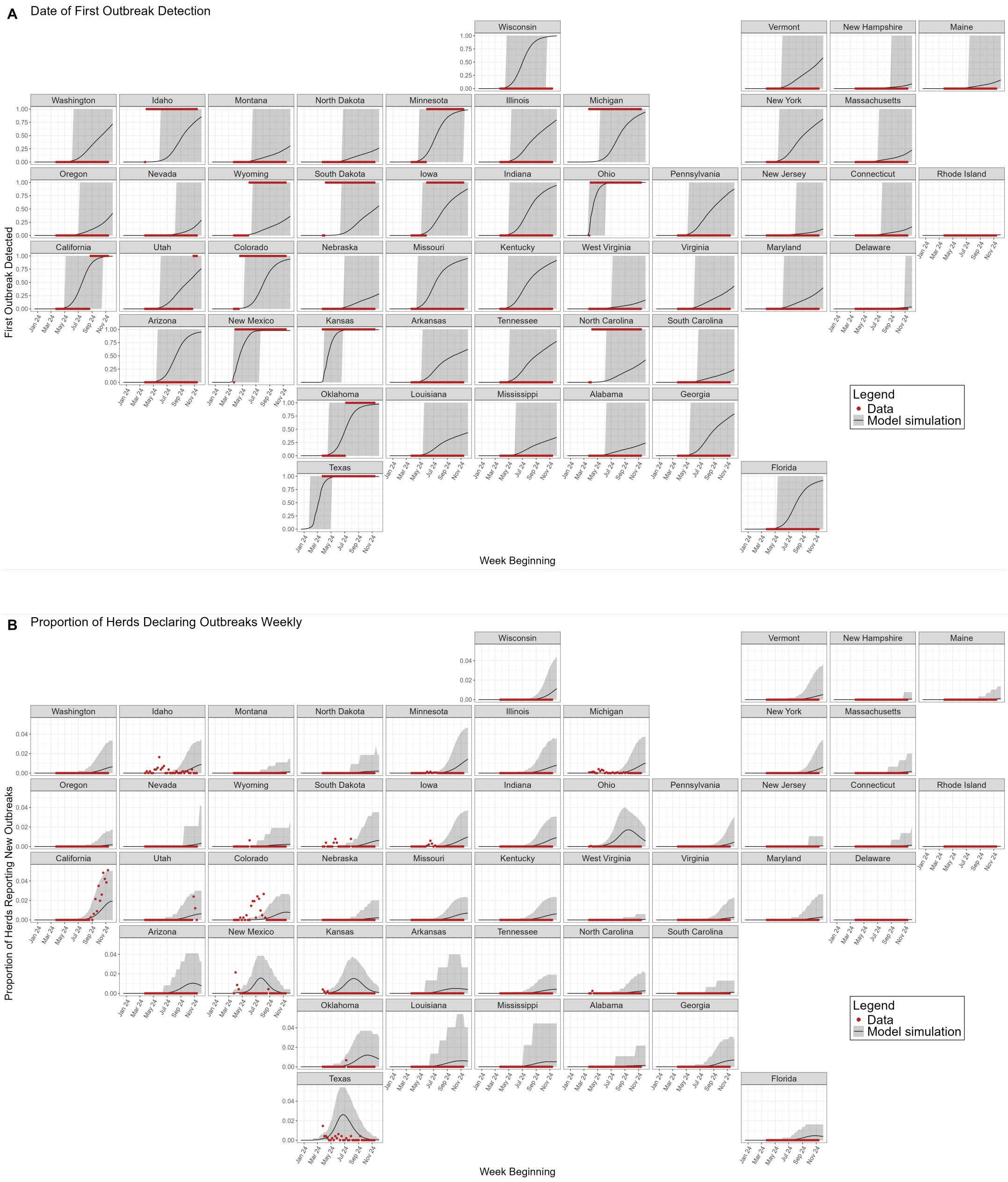

**Figure S21:** This is a recreation of Figure 2 from the main manuscript when using alternate ascertainment rate assumptions as outlined in Equation (22). **(A)** shows the time at which the first outbreak is detected in a state. **(B)** shows the proportion of herds in each state which report new outbreaks per state each week, accounting for under-reporting. Red points depict real world data. The black line depicts the model mean, the shaded grey region depicts the 95% credible interval (95% CrI).

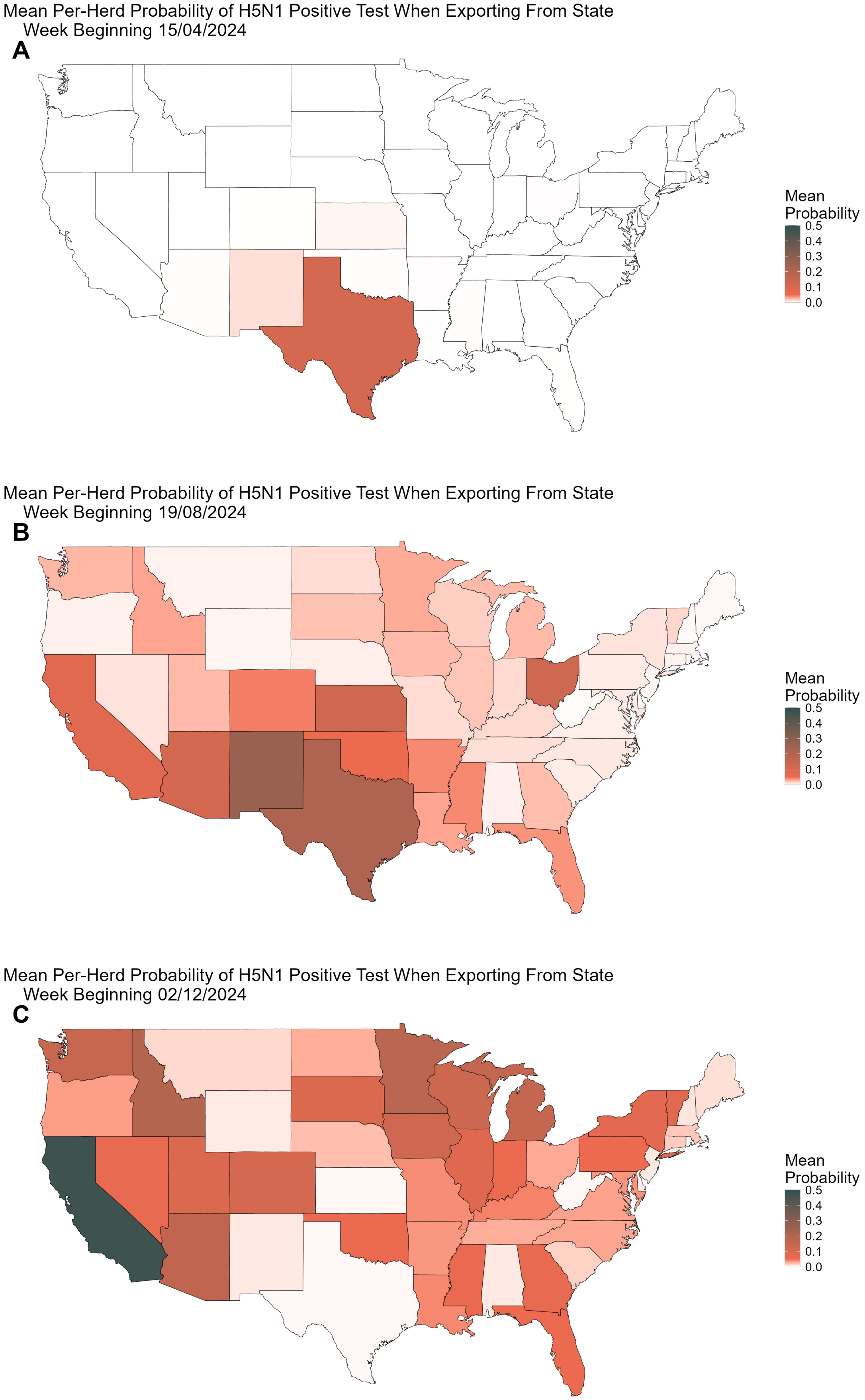

**Figure S22:** This is a recreation of Figure 4 from the main manuscript when using alternate ascertainment rate assumptions as outlined in Equation (22). When moving cattle inter-state, up to 30 cattle will be tested for H5N1. Above panels show the state average probability of such an export testing positive over time: **(A)** week beginning April 15th 2024, **(B)** week beginning August 19th 2024, and **(C)** week beginning December 2nd 2024.

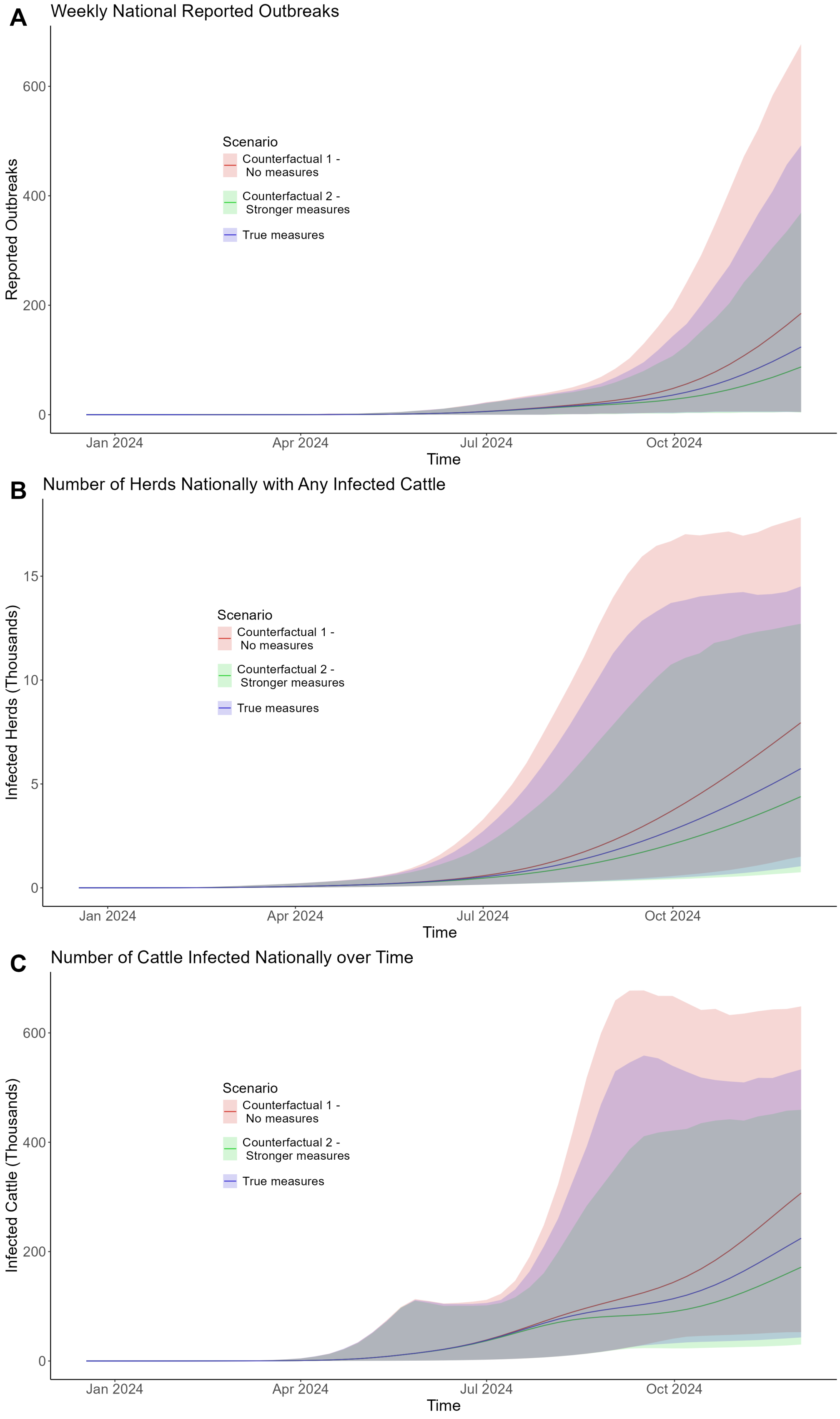

**Figure S23:** This is a recreation of Figure 5 from the main manuscript when using alternate ascertainment rate assumptions as outlined in Equation (22). **(A)** The number of new reported outbreaks weekly. **(B)** The number of herds nationally with any infected cattle. **(C)** The total number of infected cows nationally over time. Solid lines show simulation mean. Shaded regions show 95% CrI. Blue depicts baseline model assumptions. Red depicts the scenario with no border testing. Green depicts border testing of up to 100 cows, implemented 28 days earlier, on April 1st 2024.
